# Adjunctive Accelerated Theta-Burst Stimulation for Inpatient Treatment-Resistant Depression: A Randomized Clinical Trial

**DOI:** 10.64898/2026.06.25.26356553

**Authors:** Christina Müller, Marc Onken, Andrea Hildebrandt, Maximilian Kiebs, Andrew Zalesky, Robin F.H. Cash, Dirk Scheele, René Hurlemann

## Abstract

**Objective:** Hospitalized patients with treatment-resistant depression need rapid treatments, yet conventional brain stimulation protocols are prolonged. Given limited sham-controlled inpatient evidence, the authors examined whether accelerated intermittent theta-burst stimulation (iTBS) improves depressive symptoms when added to routine inpatient treatment.

**Methods:** In this double-blind, sham-controlled trial, inpatients aged 18–65 years with unipolar treatment-resistant depression were randomized to active or sham accelerated iTBS (3 sessions/day, 10 weekdays; 54,000 pulses) targeting an individualized left dorsolateral prefrontal site. Primary and secondary outcomes were change over the 2-week stimulation phase in Montgomery-Åsberg Depression Rating Scale (MADRS) and daily Beck Depression Inventory-II (BDI-II) scores. Follow-up was 6 weeks after treatment.

**Results:** Of 57 randomized patients, 51 initiated and completed stimulation (active, n=27; sham, n=24); 42 (82%) had psychiatric comorbidity. MADRS scores decreased more with active than sham stimulation (−3.54 points/week; 95% CI, −5.53 to −1.55; Cohen’s d=−0.94; model-estimated reductions, 12.06 vs 4.98 points). The effect remained significant after adjustment for the active group’s higher baseline score (d=−0.77; 95% CI, −1.32 to −0.22) and in sensitivity analyses. BDI-II scores changed in the same direction (−0.23 points/day), but not significantly when individual rates of change were modeled (95% CI, −0.57 to 0.10). During follow-up, improvements were largely maintained in both groups; the baseline-adjusted week-8 difference was not significant. No serious adverse events occurred.

**Conclusions:** Accelerated iTBS provided added benefit beyond routine inpatient care, robustly reducing clinician-rated depressive symptoms over 2 weeks; self-rated changes were directionally consistent but not robust. Multicenter trials should determine durability and optimal implementation.

**Trial Registration:** ClinicalTrials.gov Identifier: NCT04832750.

## Introduction

Major depressive disorder (MDD) is a leading cause of disability worldwide and major contributor to premature mortality and economic burden (1). A substantial proportion of patients fail to achieve adequate symptom relief with first-line pharmacotherapy and psychotherapy, resulting in treatment-resistant depression (TRD), which is associated with prolonged suffering, greater functional impairment, and repeated health-care utilization including repeated hospitalizations compared to treatment-responsive patients (2). In inpatient psychiatric settings, depressive episodes are often more severe and clinically urgent, making rapid and effective interventions especially important. Repetitive transcranial magnetic stimulation (rTMS) has emerged as a safe and well-tolerated treatment option, and has shown antidepressant efficacy in TRD (3).

Yet, two central issues limit the translation of rTMS into routine inpatient care. First, standard 4-to-6-week rTMS courses can span most or all of an inpatient stay, whereas acute settings call for rapid stabilization. Second, most sham-controlled rTMS evidence has been generated in outpatient populations (4, 5). A comprehensive meta-analysis identified only a small subset of exclusively hospitalized samples and found no superior antidepressant effect (6). In contrast, naturalistic inpatient studies suggest that rTMS may be clinically useful under routine-care inpatient conditions (7–9). This divergence between controlled trial outcomes and observational clinical data has been widely discussed (10, 11) and underscores the need for prospective, sham-controlled studies conducted in naturalistic hospital environments to determine the clinical utility of rTMS as an augmentation strategy for TRD.

To address the challenge of prolonged treatment duration and optimize efficacy for severe TRD, accelerated rTMS protocols delivering multiple sessions per day over a condensed timeframe have gained momentum, especially following the Stanford Neuromodulation Therapy (12, 13) trials. These trials achieved high rapid remission rates with 10 daily sessions over five days. However, translating such intensive regimens into widely accessible standard public healthcare systems presents major logistical and economic challenges. Protocols requiring approximately 10 hours of daily clinical management are difficult to integrate into routine clinical workflows and, in inpatient settings, limit participation in the multimodal therapeutic program, which combines pharmacotherapy, psychotherapy, and group, occupational, creative, and exercise therapies. Because active rTMS has outperformed sham when added to psychological or other nonpharmacological treatments (14), stimulation may complement this program, provided that accelerated protocols are compatible with the length of an inpatient stay and the daily therapeutic schedule.

The present randomized, double-blind, sham-controlled trial evaluated an accelerated intermittent theta-burst stimulation (iTBS) protocol delivering three daily sessions over 10 weekdays as an augmentation strategy for hospitalized patients with TRD.

Crucially, the trial was conducted under naturalistic inpatient conditions, permitting typical psychiatric comorbidities and concurrent medication adjustments within broader multimodal care, while the sham-controlled design allowed rTMS-specific effects to be distinguished from symptom improvement related to routine inpatient care. We hypothesized that a clinically implementable accelerated iTBS protocol would provide added antidepressant benefit beyond ongoing inpatient treatment.

## Methods

### Study design

We conducted a double-blind, randomized, sham-controlled trial. The broader trial protocol, which included further outcomes and patient groups, was prospectively registered at ClinicalTrials.gov (NCT04832750), and the analytical plan for the current investigation on the Open Science Framework (doi:10.17605/OSF.IO/6EMSU). The analysis plan was registered on November 7, 2024, after recruitment and before unblinding (December 2024); protocol, plan, and deviations are provided in Supplement 1. Procedures complied with the Declaration of Helsinki and were approved by the Medical Ethics Committee of the University of Oldenburg [2020-144].

### Participants

Based on an a priori power analysis (Supplement 2), we targeted 50 participants and planned to recruit 60. Participants were recruited via clinical referrals and community outreach at the Department of Psychiatry, University of Oldenburg located at the Karl-Jaspers-Klinik, Bad Zwischenahn, Germany, from May 2021 to May 2024 (last follow-up, August 2024). Inclusion criteria were age 18 to 65 years, a current moderate-to-severe major depressive episode (Structured Clinical Interview for DSM-5), and a TRD (≥1 failed adequate antidepressant trial during the current episode). To maintain a clinically representative inpatient sample, psychiatric comorbidities were permitted excluding acute suicidality and, as a precaution for the high-dose accelerated protocol, current substance dependence and psychotic or bipolar disorder. Suicidality led to exclusion only when it precluded safe participation, as judged clinically without a rating-scale cutoff; no screened patient was excluded for this reason. Further exclusion criteria comprised standard neurological, MRI or rTMS (15) safety contraindications. All participants provided written informed consent after receiving a complete description of the study.

### Randomization and Blinding

Participants were randomly assigned 1:1 to active or sham stimulation using a computer-generated sequence of permuted blocks, applied unchanged throughout recruitment. To conceal allocation, the sequence was held by a researcher with no other role in the study, and stimulation was delivered with identical, code-labelled active and sham coils. Participants, raters, stimulation staff, ward clinicians, and investigators were blind to allocation. Blinding was assessed with treatment guesses, summarized with the Bang blinding index, guess confidence, and session tolerability ratings (Supplement 2); guesses were collected after the 6-week follow-up and before unblinding.

### Intervention

Neuronavigated accelerated iTBS was delivered with the Apollo TMS system (MAG & More) at 90% resting motor threshold: 1,800 pulses/session, 3 afternoon sessions/day separated by 50-minute intervals for 10 weekdays (30 sessions; 54,000 pulses). Participants underwent structural and resting-state MRI for individualized connectivity-guided target selection of the left dorsolateral prefrontal cortex (DLPFC) cluster most anticorrelated with the subgenual anterior cingulate cortex independent of allocation, following Cash et al. (16) (Supplement 2). Post hoc quality control identified a template-orientation mismatch in the targeting pipeline, resulting in mean differences of 8.6 mm (active) and 10.4 mm (sham) between original and corrected target coordinates. These differences were unrelated to MADRS improvement (Supplement 2). Stimulation was added to routine multimodal inpatient treatment, which included pharmacotherapy, individual and group psychotherapy, and occupational, creative, and exercise therapies. All participants had access to the same range of inpatient treatment options provided by clinicians blind to allocation throughout hospitalization. Benzodiazepines and bupropion were prohibited during the stimulation phase (15, 17).

### Clinical outcomes

The primary outcome was change in clinician-rated depression severity assessed with the Montgomery-Åsberg Depression Rating Scale (MADRS) (18). The secondary outcome was change in self-reported Beck Depression Inventory-II (BDI-II) (19). The assessment obtained immediately before the first iTBS session served as treatment baseline. MADRS interviews were conducted at 7-day intervals: baseline, mid-treatment, and post-treatment. To capture high-resolution symptom trajectories, BDI-II was administered daily before the first iTBS session and at post-treatment. Both outcomes were reassessed by telephone at 6-week follow-up.

Baseline variables encompassed demographic factors (age, self-reported sex and gender identity, employment), illness chronicity (total illness duration, duration of the current episode, chronic MDD), treatment-resistance indices (Maudsley Staging Method (20), MSM; number of antidepressant and augmentation trials, and hospitalizations), and clinical variables, including comorbidities (SCID-5 (21, 22) and medical records).

### Safety and side effects

Adverse events and common side effects, including headache, nausea, discomfort, and fatigue, were recorded on each treatment day irrespective of attributed cause.

### Statistical analysis

Efficacy analyses included all participants who initiated stimulation (modified intention-to-treat population, n=51); because none discontinued, this equaled the preregistered per-protocol set, and the preregistered intention-to-treat sensitivity analysis for ≥5 discontinuations was not triggered. A post hoc sensitivity analysis included all 57 randomized participants; because the 6 non-initiators had no MADRS ratings, including at baseline, their values were multiply imputed (Supplement 2). All *P values* were 2-sided; confirmatory *P values* were adjusted using the Benjamini-Hochberg false discovery rate procedure and reported as *P*_FDR_.

#### Confirmatory analyses

MADRS scores were analyzed using latent growth models (LGMs) across three time points. For BDI-II, preregistered LGMs were estimated but showed inadequate fit for the daily data; BDI-II trajectories were therefore analyzed using linear mixed models (LMMs), which are less strict and more parsimonious, offering benefits when working with small samples (23). Models followed the preregistered sequence: linear and non-parametric time functions were compared first, treatment group was added next; prespecified covariates (age, sex, chronic MDD) and current episode duration as moderator were examined separately. Because random-intercept models assume a common rate of change for all patients in a group, all BDI-II models were fitted both with random intercepts and with additional random slopes for time; both versions are reported. Final models were selected based on fit, parsimony, and information criteria. Non-preregistered sensitivity models adjusted the MADRS slope for baseline severity, medication changes during stimulation, or rater (Supplement 2). Missing data were handled under missing-at-random assumptions using robust maximum likelihood for LGMs. LMMs used maximum likelihood for fit indices and model comparisons and restricted maximum likelihood for parameter estimates. Effect-size and model-fit details are in Supplement 2.

#### Exploratory analyses

Deviating from the preregistration due to poor LGM fit and baseline severity differences in MADRS, follow-up trajectories were analyzed using a piecewise LMM with a knot at Week 2, separating acute treatment and follow-up phases. For the baseline-adjusted week-8 difference, the group difference at baseline was constrained to zero; further sensitivity analyses are described in Supplement 2.

Clinical response (≥50% symptom reduction) and remission (MADRS<10 (24); BDI-II<13 (25)) rates were analyzed using Chi-square or Fisher’s exact tests, with odds ratios (OR) and respective CIs. Additional non-preregistered baseline-adjusted Firth logistic regression sensitivity analyses were conducted for MADRS outcomes to account for baseline differences. Additional moderation analyses of predefined demographic, clinical/psychological, and social baseline variables are reported in Supplement 2.

All analyses were performed in R (version 4.3.1) using lavaan for LGMs and lme4 for LMMs. Additional methodological details are provided in Supplement 2.

## Results

### Demographics

A total of 128 patients were assessed for eligibility, 57 were randomized; 51 patients received and completed assigned intervention (sham: *n*=24; active: *n*=27). Six randomized patients (3 per group) did not start stimulation (reasons in Figure 1); none discontinued thereafter. Baseline demographic and clinical characteristics are summarized in Table 1. The final sample represented a clinically complex inpatient TRD sample, with long illness and current-episode durations, multiple prior treatment failures, moderate-to-severe treatment resistance, and frequent psychiatric comorbidity. All participants had at least two failed adequate trials in the current episode, meeting common TRD criteria (2). Overall, 42 participants (82%) had at least one comorbid psychiatric diagnosis. Baseline characteristics, comorbidities, concomitant medication classes, and medication changes were comparable between groups (Table 1; Tables S1 and S2); none received electroconvulsive therapy or (es)ketamine. Treatment-baseline MADRS scores were higher in the active than in the sham group (mean±SD, 28.4±7.1 vs 23.1±8.1); thus baseline-adjusted sensitivity analyses are reported below.

**Figure 1.**
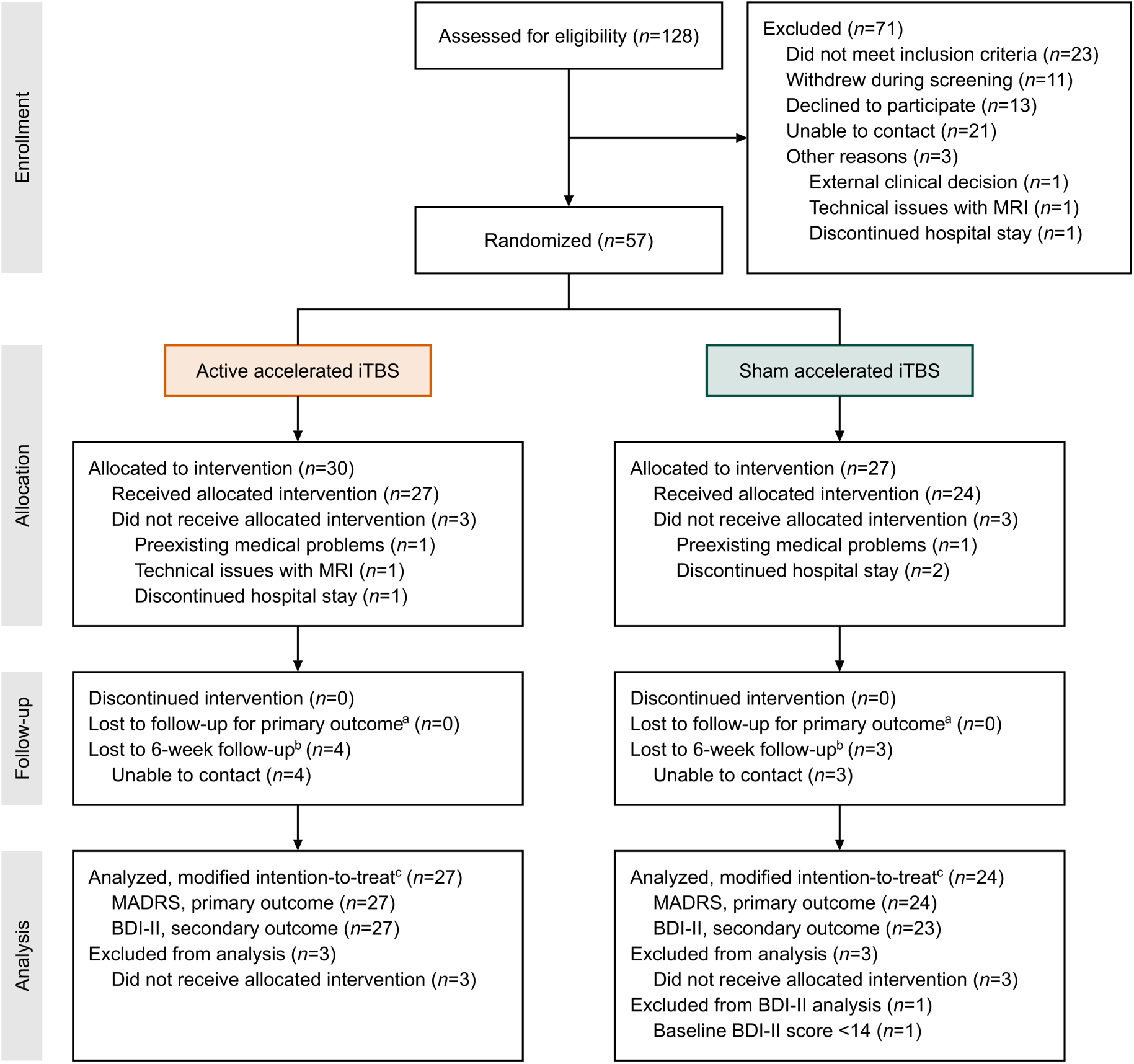
CONSORT flow diagram of participant enrollment, allocation, follow-up, and analysis. ^a^ MADRS ratings were missing for 1 participant per group (sham, week 0; active, weeks 1 and 2); both participants remained in the analyses (full-information maximum likelihood). ^b^ Lost to follow-up for the MADRS. Week-8 BDI-II scores were nevertheless obtained from 1 of these active and 2 of these sham participants, and 2 other sham participants had a week-8 MADRS but no BDI-II rating (BDI-II by online questionnaire, MADRS by telephone interview). Week-8 data were therefore available for the MADRS from 23 active and 21 sham participants and for the BDI-II from 24 active and 20 sham participants; the follow-up models included all participants. ^c^ Modified intention-to-treat population: all randomized participants who initiated stimulation. The 6 non-initiators had no MADRS ratings and no treatment-baseline BDI-II score (both first obtained immediately before the first stimulation session). A post hoc sensitivity analysis of the primary outcome included all 57 randomized participants, with the MADRS values of the non-initiators multiply imputed (Table S5 in Supplement 2). Abbreviations: BDI-II, Beck Depression Inventory-II; iTBS, intermittent theta-burst stimulation; MADRS, Montgomery-Åsberg Depression Rating Scale; MRI, magnetic resonance imaging.

**Table 1.** Baseline demographic and clinical characteristics of participants in the trial.

| Variable | Sham (N=24) |  | Active (N=27) |  |
|---|---|---|---|---|
|  | N | % | N | % |
|---|---|---|---|---|
| **Sex<sup>a</sup>** |  |  |  |  |
| Female | 13 | 54 | 16 | 59 |
| Male | 11 | 46 | 11 | 41 |
| **Gender identity<sup>a</sup>** |  |  |  |  |
| Woman | 13 | 54 | 16 | 59 |
| Man | 11 | 46 | 10 | 37 |
| Diverse | 0 | 0 | 1 | 4 |
| **Employment status** |  |  |  |  |
| Not employed | 13 | 54 | 15 | 56 |
| Employed | 11 | 46 | 12 | 44 |
|  | Mean | SD | Mean | SD |
| **Age (years)** | 40 | 12 | 40 | 14 |
| **Education (years)** | 16 | 3 | 15 | 3 |
| **MADRS score (baseline)<sup>b</sup>** | 23.1 | 8.1 | 28.4 | 7.1 |
| **BDI-II score (baseline)<sup>b</sup>** | 26.9 | 12.1 | 28.2 | 10.5 |
| **Total duration of illness (years)** | 12.4 | 8.8 | 12.7 | 8.1 |
| **Duration of current episode (months)** | 42.5 | 61.6 | 57.4 | 39.2 |
| **Maudsley Staging Method (MSM)<sup>c</sup>** | 10.5 | 2.3 | 11.3 | 1.9 |
| **Number of antidepressant trialsd** | 6.4 | 3.3 | 7.4 | 4.2 |
| **Number of augmentation trials** | 3.5 | 1.5 | 4.8 | 3.9 |
| **Number of previous hospitalizations** | 2.5 | 2.5 | 2.7 | 2.5 |
| **Motor threshold (% of machine output)** | 51.3 | 8.9 | 50.8 | 7.6 |
| **Treatment intensity (% of machine output)** | 45.4 | 8.0 | 45.5 | 5.8 |
| **Treatment intensity (% of motor threshold)<sup>e</sup>** | 88.3 | 3.5 | 89.1 | 2.8 |

Abbreviations: BDI-II, Beck Depression Inventory-II; MADRS, Montgomery-Åsberg Depression Rating Scale; rTMS, repetitive transcranial magnetic stimulation; SD, standard deviation.
Race and ethnicity were not assessed because these characteristics are not routinely collected in clinical research in Germany (racial or ethnic origin is a special category of personal data under the EU General Data Protection Regulation).
<sup>a</sup> Sex and gender identity were self-reported (gender identity: woman, man, or diverse, the third gender option in German civil law). Analyses used sex; the participant with diverse gender identity reported male sex.
<sup>b</sup> Baseline assessed after admission
<sup>c</sup> Prior electroconvulsive therapy: sham, 1 (4%); active, 2 (7%). Prior rTMS: sham, 0 (0%); active, 2 (7%).
<sup>d</sup> All participants had ≥2 failed antidepressant trials in the current episode.
<sup>e</sup> Mean intensities slightly below 90% reflect tolerability-based titration (Supplement 2).

### Primary outcome

Observed MADRS trajectories and percentage changes across the acute treatment and follow-up phases are illustrated in Figure 2A–B. Overall, MADRS scores decreased by −4.40 points/week (95% CI, −5.53 to −3.28; z=−7.68, *P*_FDR_<.001), corresponding to a model-estimated reduction of −8.81 points over the acute treatment phase (Table 2).

**Figure 2.**
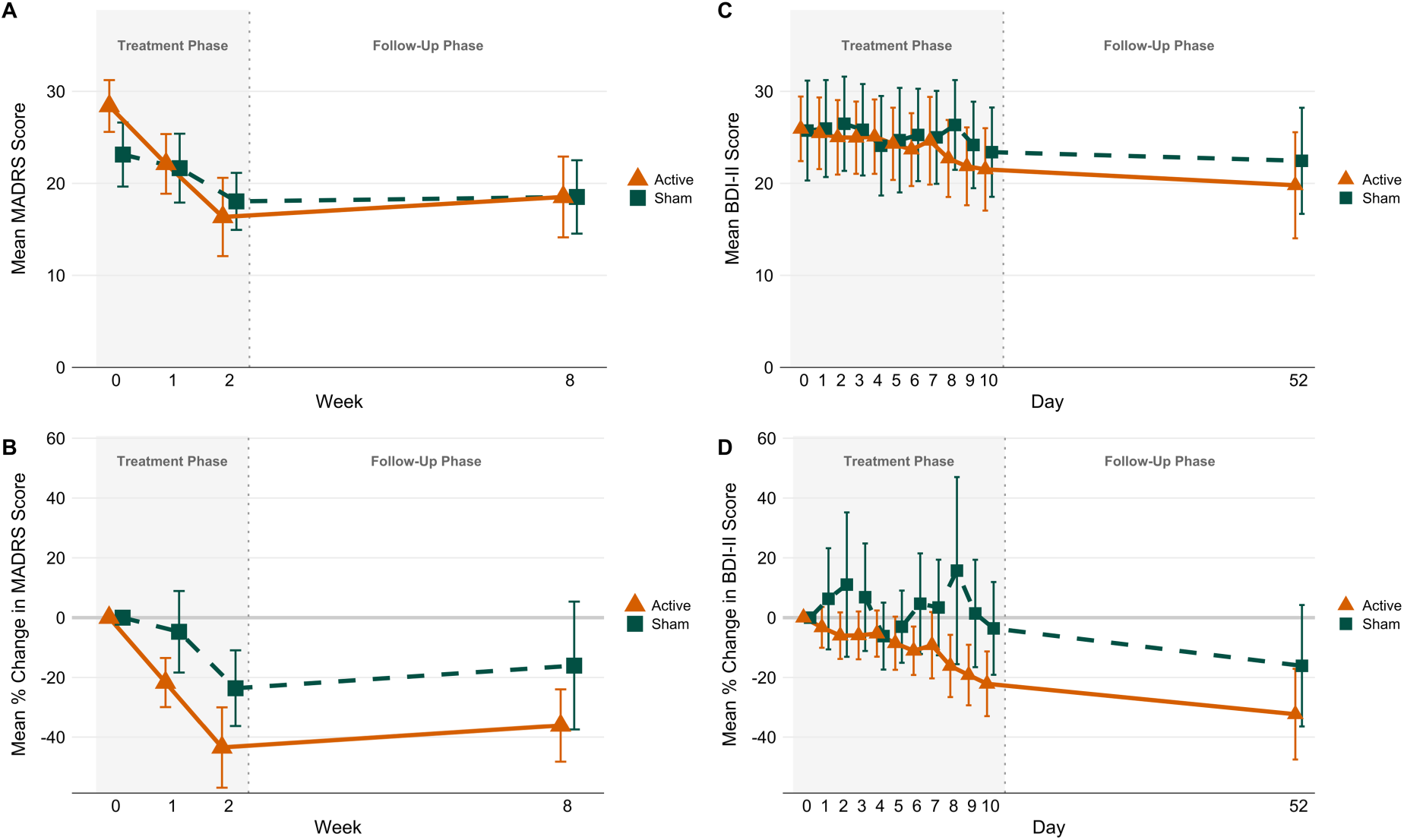
Observed MADRS and BDI-II scores during treatment and follow-up in participants receiving active (*n*=27) or sham (*n*=24) accelerated iTBS^a^. ^a^ For the BDI-II, sham *n*=23 (one participant with a baseline BDI-II score <14 excluded). Panels A and C show observed mean scores; panels B and D show the mean of individual percentage changes from baseline (MADRS, week 0; BDI-II, day 0). MADRS was rated at weeks 0, 1, 2, and 8; BDI-II daily during treatment (days 0–10) and at follow-up (day 52). The shaded area marks the 2-week treatment phase; the follow-up interval is shown compressed. Error bars indicate 95% confidence intervals of the mean. Abbreviations: BDI-II, Beck Depression Inventory-II; iTBS, intermittent theta-burst stimulation; MADRS, Montgomery-Åsberg Depression Rating Scale.

**Table 2.** Model-estimated longitudinal trajectories and treatment effects for clinician-rated and self-reported depression severity.

| Outcome and comparison<sup>a</sup> | Hypothesis<sup>b</sup> | Estimate<sup>c</sup> | 95% CI | Cohen’s *d*<sup>d</sup> | z or t<sup>e</sup> | df<sup>e</sup> | P<sup>f</sup> |
|---|---|---|---|---|---|---|---|
| **MADRS (primary outcome)** |  |  |  |  |  |  |  |
| Treatment phase: change from baseline to week 2 |  |  |  |  |  |  |  |
| All participants (N=51) | H1 | −8.81 | −11.05 to −6.56 | −1.17 | −7.68 |  | <.001 |
| Sham group (N=24) |  | −4.98 | −7.06 to −2.91 |  |  |  |  |
| Active group (N=27) |  | −12.06 | −15.46 to −8.67 |  |  |  |  |
| Difference, active vs. sham | H2 | −7.08 | −11.05 to −3.10 | −0.94 | −3.49 |  | <.001 |
| Difference, adjusted for baseline severity |  | −5.84 | −9.99 to −1.69 | −0.77 | −2.76 |  | .006 |
| Episode duration and change from baseline to week 2<sup>g</sup> |  |  |  |  |  |  |  |
| Association with change | H3a | 2.66 | −2.14 to 7.47 |  | 1.09 |  | .28 |
| Difference in association, active vs. sham | H3b | 5.89 | −3.75 to 15.52 |  | 1.20 |  | .28 |
| Follow-up: change from week 2 to week 8<sup>h</sup> |  |  |  |  |  |  |  |
| Sham group |  | −0.66 | −3.60 to 2.27 |  |  |  |  |
| Active group |  | 1.68 | −1.15 to 4.51 |  |  |  |  |
| Difference, active vs. sham |  | 2.34 | −1.73 to 6.42 | 0.31 | 1.14 | 139.6 | .26 |
| **BDI-II (secondary outcome)**<sup>i</sup> |  |  |  |  |  |  |  |
| Treatment phase: change from baseline to week 2 |  |  |  |  |  |  |  |
| All participants (N=50) | H1 | −3.05 | −4.74 to −1.36 | −0.28 | −3.63 | 49.0 | .003 |
| Sham group (N=23) |  | −1.79 | −4.26 to 0.67 |  |  |  |  |
| Active group (N=27) |  | −4.12 | −6.40 to −1.85 |  |  |  |  |
| Difference, active vs. sham | H2 | −2.33 | −5.69 to 1.03 | −0.22 | −1.40 | 48.0 | .34 |
| Episode duration and change from baseline to week 2<sup>g</sup> |  |  |  |  |  |  |  |
| Association with change | H3a | 1.97 | −2.71 to 6.66 |  | 0.85 | 46.0 | .40 |
| Difference in association, active vs. sham | H3b | 4.16 | −5.21 to 13.53 |  | 0.89 | 46.0 | .40 |
| Follow-up: change from week 2 to week 8<sup>h</sup> |  |  |  |  |  |  |  |
| Sham group |  | −3.47 | −5.21 to −1.73 |  |  |  |  |
| Active group |  | −2.96 | −4.55 to −1.37 |  |  |  |  |
| Difference, active vs. sham |  | 0.51 | −1.84 to 2.86 | 0.05 | 0.43 | 497.5 | .67 |

Abbreviations: BDI-II, Beck Depression Inventory-II; CI, confidence interval; FDR, false discovery rate; MADRS, Montgomery-Åsberg Depression Rating Scale; SD, standard deviation.
<sup>a</sup> Model terms: all participants, mean change (slope) over time; sham and active groups, model-derived change within each group; difference, effect of group on change; difference adjusted for baseline severity, effect of group on change adjusted for the latent baseline (MADRS only); association with change, effect of current episode duration on change; difference in association, group × episode duration interaction; follow-up, effect of group on change from week 2 to week 8 (piecewise model).
<sup>b</sup> H1–H3 denote the preregistered confirmatory hypotheses (doi:10.17605/OSF.IO/6EMSU), each tested for MADRS and BDI-II: H1, depressive symptoms decrease over the 2-week stimulation phase; H2, this decrease differs between active and sham stimulation; H3, current episode duration is associated with symptom change (H3a) and moderates the group difference (H3b). Rows without a label are exploratory or sensitivity analyses.
<sup>c</sup> Model-estimated total change over the period in scale points (MADRS, 0–60; BDI-II, 0–63); negative values indicate improvement. Differences are active minus sham.
<sup>d</sup> Estimate divided by the pooled baseline SD (MADRS, 7.56; BDI-II, 10.72). Not reported for episode duration and its interaction with group: for a continuous predictor, a standardized effect depends on the chosen contrast (here, a 10-fold longer episode), would not be comparable with the group differences, and could easily be misinterpreted.
<sup>e</sup> z, Wald z of the latent growth model (MADRS, treatment phase; no df); t, t test of the linear mixed model with Satterthwaite df (MADRS follow-up and BDI-II).
<sup>f</sup> P values for the confirmatory hypotheses (H1–H3) are FDR-adjusted; all other P values are unadjusted (exploratory analyses).
<sup>g</sup> Difference in change (scale points) per 10-fold longer current episode (episode duration log<sub>10</sub>-transformed and mean-centered, not standardized). Both rows come from the moderation model with effect-coded group (sham, −0.5; active, +0.5): association with change, mean association across both groups; difference in association, active minus sham.
<sup>h</sup> Estimated with all participants (likelihood-based estimation, assuming data missing at random). Week-8 ratings were available for MADRS, sham n=21, active n=23; BDI-II, sham n=20, active n=24.
<sup>i</sup> Linear mixed model with random intercepts and random slopes for time (restricted maximum likelihood); results of the random-intercept model are reported in the text and in Supplement 3. BDI-II was rated daily during treatment (days 0–10).

Adding treatment allocation yielded a significant time-by-group interaction. MADRS scores decreased more steeply with active iTBS than sham (β=−3.54 points/week; 95% CI, −5.53 to −1.55; z=−3.49, *P*_FDR_<.001), corresponding to a model-estimated between-group difference of −7.08 points over the acute treatment phase and a large effect (Cohen’s *d*=−0.94). Adding baseline MADRS severity as a sensitivity covariate did not significantly influence symptom change (β=−0.12, z=−1.18, *P*=.24), and the treatment effect remained significant (β=−2.92 points/week; 95% CI, −4.99 to −0.85; z=−2.76, *P*=.006; Cohen’s *d*=−0.77). Moreover, the effect remained significant in all further sensitivity analyses: week-2 analyses of covariance and mixed models for repeated measures, inclusion of all 57 randomized participants by multiple imputation, and adjustment for medication changes during stimulation or for rater (Table S5).

### Secondary outcome

Observed daily BDI-II trajectories and percentage changes are shown in Figure 2C–D. Across the full cohort, BDI-II scores decreased by −0.31 points/day (95% CI, −0.40 to −0.21; t(499.0)=−6.62, *P*_FDR_<.001), corresponding to a model-estimated reduction of −3.05 points over the acute treatment phase. This decrease remained significant with random slopes (Table 2).

Adding treatment allocation, BDI-II scores decreased more steeply with active iTBS than with sham (β=−0.23 points/day; Cohen’s *d*=−0.22), corresponding to a model-estimated between-group difference of −2.33 points over the 10-day acute treatment phase. The difference was significant in the random-intercept model (95% CI, −0.41 to −0.05; t(498.0)=−2.53, *P*_FDR_=.02) but not in the random-slope model (95% CI, −0.57 to 0.10; t(48.0)=−1.40, *P*_FDR_=.34; Table 2), which accounts for the considerable variation of individual rates of change (SD, 0.52 points/day; χ²(1:2)=66.66, *P*<.001).

### Covariates and moderation

For both outcomes, male participants had lower baseline scores (MADRS, β=−4.66; z=−2.65, *P*=.008; BDI-II, β=−7.70; t(47.0)=−2.62, P=.01), whereas age and sex were not associated with change. Chronic MDD was associated with higher baseline MADRS scores (β=5.44; z=1.97, *P*=.048) and, in the random-intercept model only, with a steeper BDI-II decrease (β=−0.24 points/day; t(497.0)=−2.64, P=.009; random slope, t(47.0)=−1.46, P=.15). Current episode duration, a preregistered moderator, was not associated with symptom change and did not moderate the treatment effect on either outcome (all *P*_FDR_≥.12). In the final MADRS model, active treatment allocation remained the only significant predictor of change (β=−3.60 points/week; 95% CI, −5.60 to −1.60; z=−3.52, *P*<.001; Figure S2; Supplement 3, Tables S6 and S7).

### Exploratory outcomes

#### Follow-up trajectories

Improvements achieved during stimulation were largely maintained in both groups: from week 2 to the 6-week follow-up, MADRS scores remained stable and BDI-II scores decreased slightly, with no group difference in the rate of change (MADRS, t(139.6)=1.14, *P*=.26; BDI-II, t(497.5)=0.43, *P*=.67; Table 2). Despite higher baseline MADRS scores in the active group, both groups had the same observed mean score at week 8 (18.5). After baseline adjustment, the week-8 difference favored active stimulation in all analyses, including those addressing missing follow-up data, but none was significant (piecewise model, −3.0 points; 95% CI, −6.8 to 0.8; t(169.7)=−1.56, *P*=.12; Table S5).

#### Response and remission rates

Clinical response (≥50% symptom reduction) and remission (MADRS<10; BDI-II<13) were evaluated as exploratory acute-phase endpoints (Figure 3; Table S3). The active iTBS group demonstrated a significantly higher MADRS response rate compared to sham (11/26 [42.3%] vs 3/23 [13.0%]; OR=4.73; 95% CI, 1.01 to 31.09; Fisher exact test, *P*=.03), while numerically but not statistically higher MADRS remission rate (7/26 [26.9%] vs 3/24 [12.5%]; OR=2.53; 95% CI, 0.49 to 17.36; *P*=.29), BDI-II response (4/27 [14.8%] vs 1/22 [4.5%]; OR=3.57; 95% CI, 0.32 to 188.07; *P*=.36), or BDI-II remission (7/27 [25.9%] vs 5/22 [22.7%]; OR=1.19; 95% CI, 0.27 to 5.68; χ²(1)=0.07, *P*=.80).

**Figure 3.**
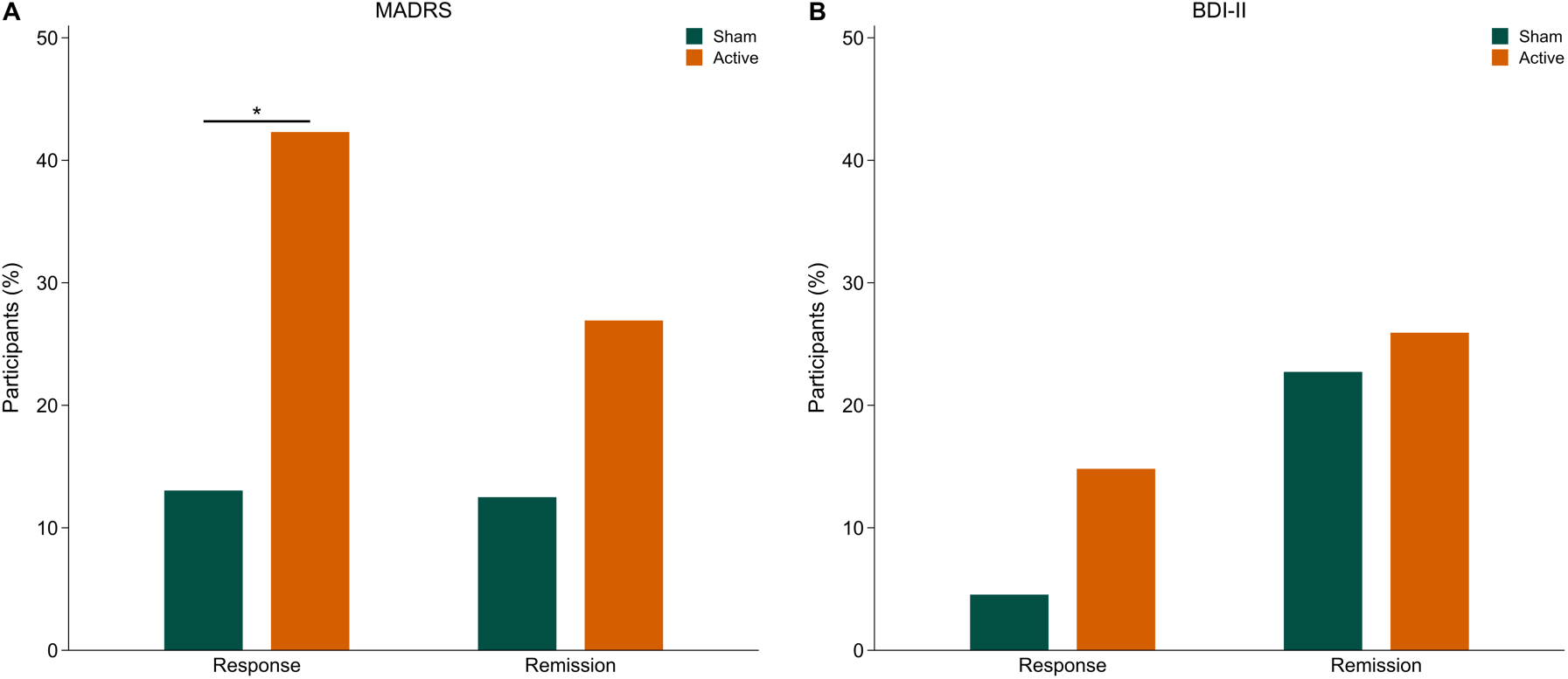
Response and remission at week 2 in participants receiving active (*n*=27) or sham (*n*=24) accelerated iTBS^a^. ^a^ Response, ≥50% reduction from treatment baseline; remission, MADRS score <10 or BDI-II score <13. Participants meeting the criterion/participants with the required ratings (response: treatment baseline and week 2; remission: week 2): MADRS response, sham 3/23 (13.0%), active 11/26 (42.3%); MADRS remission, sham 3/24 (12.5%), active 7/26 (26.9%); BDI-II response, sham 1/22 (4.5%), active 4/27 (14.8%); BDI-II remission, sham 5/22 (22.7%), active 7/27 (25.9%). Risk differences (active minus sham, percentage points, with Newcombe hybrid score 95% CIs): MADRS response, 29.3 (3.9 to 49.8); MADRS remission, 14.4 (−8.3 to 35.2); BDI-II response, 10.3 (−9.1 to 28.3); BDI-II remission, 3.2 (−21.1 to 25.8). For 1 sham participant, the last BDI-II rating was obtained before the final stimulation session; it was included in the longitudinal models but not used as post-treatment end point. *Preregistered unadjusted analysis (MADRS response, Fisher exact test, *P*=.03). Baseline-adjusted analyses: see Results. Abbreviations: BDI-II, Beck Depression Inventory-II; iTBS, intermittent theta-burst stimulation; MADRS, Montgomery-Åsberg Depression Rating Scale.

In baseline-adjusted logistic sensitivity models for MADRS outcomes, active iTBS was associated with higher odds of response (OR=8.53; 95% CI, 1.88 to 57.40; χ²(1)=8.22, *P*=.004) and remission (OR=5.73; 95% CI, 1.11 to 44.73; χ²(1)=4.41, *P*=.04; Table S3).

### Safety and Blinding

No serious adverse events occurred, and no seizures, syncope, (hypo)manic symptoms, treatment-emergent suicidal ideation, or cognitive complaints were observed. Headache (sham, 10 of 24; active, 15 of 27), fatigue (10 vs 8), discomfort (6 vs 7), and nausea (5 vs 4) did not differ significantly between groups (Table S4).

Of 38 participants who provided guesses, 24 (63%) guessed correctly (Fisher exact test, P=.12); Bang blinding indices were 0.20 (95% CI, −0.23 to 0.63) for active and 0.33 (95% CI, −0.10 to 0.77) for sham stimulation. Confidence did not differ between correct and incorrect guesses, and tolerability neither differed between groups nor predicted guesses (Supplement 2).

## Discussion

In this randomized, double-blind, sham-controlled trial, accelerated, high-dose iTBS delivered as three daily sessions over 10 weekdays produced steeper acute symptom reduction than sham stimulation when added to ongoing multimodal inpatient care in hospitalized patients with TRD. During this acute stimulation phase, active iTBS resulted in a model-estimated 12.06-point reduction on the primary clinician-rated MADRS, compared with a 4.98-point reduction in the sham group, corresponding to a large between-group treatment effect. Self-reported BDI-II scores changed in the same direction, but this smaller effect was not robust to between-patient variation in rates of change. Improvements were largely maintained through follow-up in both groups; the baseline-adjusted advantage of active stimulation at week 8 was smaller and not significant, so a lasting between-group benefit remains to be established.

The higher baseline MADRS score in the active group raises the possibility of regression to the mean. However, inclusion did not depend on a MADRS threshold, baseline severity was not significantly associated with subsequent change, and baseline adjustment reduced the 2-week difference by 1.23 points (26). Adjustment also did not weaken the response and remission results, and BDI-II trajectories, without baseline imbalance, pointed in the same direction.

These findings help address a major limitation of the rTMS evidence base in TRD: sham-controlled data from hospitalized patients remain sparse, and prior meta-analytic work did not identify significant antidepressant effects within the small subset of exclusively inpatient samples (6). Naturalistic inpatient studies have nevertheless reported clinically meaningful improvement under routine-care conditions (7, 9) and one large inpatient cohort exhibited shorter hospitalization with twice-daily compared to standard rTMS (8). The present trial therefore adds randomized sham-controlled evidence to the limited and predominantly naturalistic inpatient literature.

The clinical relevance should be interpreted considering the cohort’s clinical complexity, reflected in protracted depressive episodes, high treatment resistance, and frequent psychiatric comorbidity. This profile is clinically important as depressive and anxiety symptom severity, treatment resistance, prior treatment failures, chronicity, psychiatric admissions, and functional status predicted poorer antidepressant outcomes following rTMS-based treatments (27, 28). Current episode duration, examined as a preregistered moderator, however, did not modify MADRS or BDI-II treatment trajectories in the present trial. Evidence for the impact of psychiatric comorbidity beyond anxiety remains mixed: broader antidepressant-treatment data indicate poorer outcomes with psychiatric comorbidities (29), whereas TMS-specific studies suggest patients with comorbid anxiety, trauma- and stressor-related, personality, or somatic symptom disorders can still experience meaningful improvement, with inconsistent evidence on whether these comorbidities attenuate antidepressant response (30, 31). Recent accelerated rTMS studies identified largely overlapping predictor profiles (32). Thus, demonstrating a meaningful treatment effect despite the cohort’s substantial clinical complexity and frequent psychiatric comorbidities reinforces the utility of accelerated rTMS in inpatient TRD samples.

Accelerated rTMS protocols address a practical limitation of conventional several-week courses by condensing treatment schedules through multiple daily sessions. Meta-analytic evidence supports rapid symptom reduction across accelerated rTMS approaches, while optimal implementation remains unresolved (4, 33). Stanford Neuromodulation Therapy studies exemplify the most intensive end of the accelerated-treatment spectrum, delivering 50 sessions over five days, and have highlighted emergency and inpatient settings as potential applications (12, 13). For routine multimodal inpatient care, however, feasibility depends not only on shortening treatment courses but also on integrating stimulation alongside psychotherapy, other ward-based interventions, medication management, and daily clinical care. The present protocol thus represents a pragmatic middle-ground schedule with three daily iTBS sessions over 10 weekdays, broadly consistent with sham-controlled outpatient evidence supporting a similar 3-session/day accelerated approach in TRD (34).

Several factors may explain the smaller, less robust self-reported effect. Clinician ratings yield on average larger effects than self-reports, also with masked raters (35, 36) and in a large sham-controlled rTMS trial (37). The MADRS was designed to be sensitive to change (18), whereas the BDI-II emphasizes self-evaluative symptoms that correlate only moderately with it (38). Moreover, in this largely naturalistic sample with frequent comorbidity, self-ratings may diverge from clinician ratings, as shown for personality disorders (39).

The exploratory response and remission analyses provide categorical translation of the continuous symptom trajectories. Active stimulation yielded higher MADRS response and numerically but not statistically higher MADRS remission than sham. Baseline-adjusted sensitivity analyses supported higher odds of both response and remission, with uncertainty reflected in wide confidence intervals. The active-arm MADRS response rate falls within the wide range reported in a recent meta-regression of accelerated protocols, which found response rates of 19.4% to 90.5% and average response and remission rates of 52.2% and 32.5%, respectively, amid heterogeneity (40). Direct comparisons, however, are limited by differences in study designs, stimulation protocols, outcome definitions, and inpatient status. The accelerated schedule was well tolerated, with no serious adverse events observed and comparable side-effect rates across groups.

The findings are strengthened by the double-blind, sham-controlled design, a representative comorbid inpatient sample, the integration of stimulation into routine multimodal inpatient care, and complete adherence after treatment initiation. Several limitations, however, should be considered. First, the single-site design and modest sample size limit precision, particularly for categorical endpoints, moderator analyses, and follow-up comparisons. Second, this integration, including medication adjustments and ward-based co-interventions, means that the findings reflect additive clinical value during hospitalization rather than standalone efficacy under stable-medication conditions; although medication patterns were comparable between groups, contributions of concurrent treatments cannot be separated. Third, the bundled protocol cannot isolate the contributions of accelerated scheduling, pulse dose, intersession interval, or individualized connectivity-guided targeting (33, 40).

In conclusion, this randomized sham-controlled trial provides evidence that an accelerated iTBS protocol can produce added antidepressant benefit when integrated into routine multimodal inpatient care for hospitalized patients with TRD. The effect was clearest for clinician-rated depressive symptoms and occurred in a clinically complex cohort characterized by chronicity, high treatment resistance, and substantial psychiatric comorbidity. Larger multicenter trials are needed to establish durability, clarify optimal stimulation parameters, and guide implementation in routine inpatient psychiatric care.

## Data Availability

The data that support the findings of this study are not publicly available due to privacy reasons but are available from the corresponding author upon reasonable request and with approval from the Institutional Review Board approval.

## Acknowledgments

The authors thank all study participants, present and former members of the Neuromodulation of Emotion Lab and the Neuroimaging Unit at the Carl von Ossietzky University of Oldenburg, and all clinical staff who contributed to the study. The study was supported by institutional funds of the Department of Psychiatry and Psychotherapy, School of Medicine and Health Sciences, Carl von Ossietzky University of Oldenburg; no project-specific external funding was received. MRI data were acquired at the Neuroimaging Unit of the Carl von Ossietzky University of Oldenburg on a 3-T scanner funded by the German Research Foundation (Deutsche Forschungsgemeinschaft [DFG], INST 184/152-1 FUGG). The multiband imaging sequence was provided by the Center for Magnetic Resonance Research, University of Minnesota. fMRI preprocessing was performed on the high-performance computing clusters CARL and ROSA at the Carl von Ossietzky University of Oldenburg, funded by the DFG (INST 184/157-1 FUGG and INST 184/225-1 FUGG) and the Lower Saxony Ministry of Science and Culture. The funders had no role in the design, conduct, analysis, or reporting of the study. The authors used Claude (Anthropic) to assist with language editing of the revised manuscript, checking analysis code and the consistency of reported values across documents; all content was reviewed and verified by the authors, who take full responsibility for it.

## Data Sharing Statement

Deidentified individual participant data underlying the reported results will be available upon reasonable request to the corresponding author, subject to a data use agreement and to the restrictions of the ethics approval and applicable data protection law, beginning with publication and ending 10 years after publication. The study protocol and statistical analysis plan are available in Supplement 1 and on the Open Science Framework.

