## Supplement 1 for "Adjunctive Accelerated Theta-Burst Stimulation for Inpatient Treatment-Resistant Depression: A Randomized Clinical Trial"

### Supplement 1: Protocol Summary, Statistical Analysis Plan, and Deviations

*Clinical antidepressant outcome analysis of the DREAMS randomized active-vs-sham iTBS inpatient trial*

#### 1. Protocol and SAP provenance

This document summarizes the trial protocol and statistical analysis plan relevant to the clinical antidepressant outcome analysis reported in the present manuscript. The original DREAMS ethics protocol and ClinicalTrials.gov registration described a broader mechanistic study of dorsolateral prefrontal cortex (DLPFC) modulation in major depressive disorder, including sleep, interoception, cognitive control, neuroimaging, biomarker, and symptom-burden outcomes. The present manuscript reports the randomized active-vs-sham intermittent theta-burst stimulation (iTBS) clinical antidepressant analysis in the inpatient major depressive disorder (MDD) sample.

The clinical antidepressant analysis was specified before analysis in the Open Science Framework preregistration (doi:10.17605/OSF.IO/6EMSU), which defined the clinical hypotheses, primary and secondary depression outcomes, model sequence, exclusion criteria, missing-data handling, and planned exploratory analyses. Older protocol and registry documents used the terms placebo and verum; in the present protocol summary and manuscript, these conditions are referred to as sham and active stimulation, consistent with common terminology in rTMS trials.

The ClinicalTrials.gov record reflected the broader DREAMS project structure, including mechanistic outcomes and healthy-control assessments. The present Supplement 1 refers to the randomized intervention component in the inpatient MDD sample that is analyzed in the manuscript.

#### 2. Administrative information

Manuscript title: Adjunctive Accelerated Theta-Burst Stimulation for Inpatient Treatment-Resistant Depression: A Randomized Clinical Trial.

Broader ethics-protocol title: Investigation of the causal relationship between prefrontal cortex hypoactivity and depression-associated changes in impulse control and interoception.

ClinicalTrials.gov registration: NCT04832750 (2021-04-06); registered title: Depression-Reduction by Accelerated Personalized NeuroModulation and Its Effects on Sleep (DREAMS).

OSF statistical analysis plan/preregistration: doi:10.17605/OSF.IO/6EMSU (2024-11-07).

Ethics approval: Medical Ethics Committee of the Carl von Ossietzky University Oldenburg, approval number 2020-144.

Protocol versions available: Version 1, September 16, 2020; Version 2, November 17, 2020; Version 3, March 18, 2021; Version 4, July 27, 2021.

Sponsor: University of Oldenburg.

Trial site: Department of Psychiatry and Psychotherapy, University of Oldenburg / Karl-Jaspers-Klinik, Bad Zwischenahn, Germany.

Principal investigators: René Hurlemann and Dirk Scheele. Study directors/coordinating investigators: Christina Müller and Marc Onken.

Funding: Budget of the University Clinic for Psychiatry and Psychotherapy.

Patient and Public Involvement: Patients and public contributors were not formally involved in developing the research question, trial design, conduct, outcome selection, or manuscript preparation. Patient interests were represented through the ethics review process, which included patient/public representatives of the Medical Ethics Committee of Carl von Ossietzky University of Oldenburg. The committee reviewed and approved the study protocol and participant-facing documents.

#### 3. Background and rationale

Major depressive disorder is a prevalent and disabling psychiatric disorder. A clinically important subgroup of patients does not respond adequately to antidepressant pharmacotherapy and psychotherapy and is classified as having treatment-resistant depression (TRD). Inpatient treatment for depression typically involves multimodal care, including pharmacotherapy, psychotherapy, occupational therapy, physical therapy, and other non-pharmacological interventions. However, patients with treatment-resistant or chronic depression often continue to experience substantial residual symptoms despite inpatient treatment.

Repetitive transcranial magnetic stimulation (rTMS) is an established noninvasive brain stimulation treatment for depression. Intermittent theta-burst stimulation (iTBS) allows shorter stimulation sessions than conventional rTMS protocols and can therefore be integrated into accelerated treatment schedules. Accelerated iTBS may be particularly relevant for inpatient settings, where treatment windows are shorter and rapid symptom stabilization is clinically important.

The original DREAMS protocol was designed as a broader mechanistic study of DLPFC modulation in major depressive disorder, with outcomes spanning sleep, interoception, cognitive control, neuroimaging, biomarkers, and symptom burden. The present manuscript focuses on the clinical antidepressant analysis of the randomized inpatient MDD sample and tests whether accelerated connectivity-guided iTBS provides an antidepressant add-on benefit beyond routine multimodal inpatient care.

#### 4. Objectives and hypotheses

Primary objective: To determine whether active connectivity-guided accelerated iTBS improves clinician-rated depressive symptoms beyond sham stimulation when administered as an add-on to routine multimodal inpatient treatment in patients with treatment-resistant MDD.

Secondary objective: To determine whether active iTBS improves self-reported depressive symptoms during the acute stimulation phase.

Preregistered confirmatory clinical hypotheses:

(H1) MADRS and BDI-II scores would decrease during the acute stimulation phase;

(H2) decreases in MADRS and BDI-II scores would be greater with active than sham iTBS; and

(H3) longer current episode duration would be associated with weaker symptom reduction (H3a) and would attenuate the active-vs-sham treatment difference (H3b).

Exploratory objectives included evaluation of 6-week follow-up trajectories, categorical response and remission, loss of response or remission at follow-up, safety and side effects, blinding integrity, exploratory multigroup latent growth parameters, predefined psychological/social-integration/employment moderators, and post hoc target-coordinate quality control.

#### 5. Trial design

This was a single-site, randomized, double-blind, sham-controlled inpatient clinical trial comparing active and sham accelerated iTBS as augmentation to routine multimodal inpatient care in patients with treatment-resistant unipolar MDD.

Participants were allocated to active or sham stimulation. All patients continued routine inpatient care. Healthy-control data and broader mechanistic DREAMS outcomes were not included in the present clinical antidepressant outcome analysis. The ethics protocol also planned subgroups of patients with depression and comorbid borderline personality disorder (BPD) or attention-deficit/hyperactivity disorder (ADHD), while the trial registry specified a subgroup with comorbid BPD. These subgroups were not recruited because recruitment was not feasible during the COVID-19 pandemic and because of staff changes. Patients were recruited from depression wards only, as a single sample in which comorbid disorders, including BPD and ADHD, were permitted (Section 7).

#### 6. Study setting

Participants were recruited and treated at the Department of Psychiatry and Psychotherapy, University of Oldenburg, located at the Karl-Jaspers-Klinik, Bad Zwischenahn, Germany.

Recruitment for the analyzed sample began in May 2021 and ended in May 2024; the last follow-up assessment took place in August 2024.

#### 7. Eligibility criteria

Eligible participants for the present clinical analysis were inpatients aged 18 to 65 years; able to provide written informed consent; fluent in German; diagnosed with unipolar MDD according to DSM-5 criteria; experiencing a current depressive episode; treatment-resistant in the current depressive episode, defined as failure to respond to at least one antidepressant treatment of adequate dose and duration; and eligible for MRI and rTMS according to safety guidelines. A moderate-to-severe episode was implied by the requirement of inpatient treatment.

The broader DREAMS ethics protocol allowed inclusion of adult patients up to 75 years. The present clinical antidepressant outcome analysis was restricted to non-geriatric adult inpatients aged 18 to 65 years, consistent with the OSF preregistration and the recruitment context of the general adult inpatient wards. No participant older than 65 years was recruited or analyzed in the inpatient clinical trial sample.

Exclusion criteria included MRI contraindications, rTMS contraindications, pregnancy or breastfeeding, acute suicidality, current substance dependence, psychotic disorder or psychotic symptoms, bipolar disorder, neurological illness, history of relevant brain injury or stroke, increased current seizure risk, conditions associated with increased intracranial pressure, inability to follow study procedures, and withdrawal of consent.

Comorbid psychiatric disorders were permitted unless covered by the exclusion criteria above, to retain a clinically representative inpatient sample. This included personality disorders; BPD was therefore not an exclusion criterion, unlike in the registry entry for the group without BPD (Section 5).

#### 8. Recruitment and informed consent

Participants were recruited via clinical referrals, waiting lists for inpatient treatment of affective disorders, direct recruitment during inpatient treatment at the University Clinic for Psychiatry and Psychotherapy / Karl-Jaspers-Klinik, and community or online outreach.

Written informed consent was obtained before study participation. Participation was voluntary, and participants could withdraw consent at any time without negative consequences for clinical care.

#### 9. Randomization and allocation concealment

Treatment allocation was performed by an independent researcher who was otherwise not involved in participant recruitment, treatment delivery, clinical assessments, or patient care.

The allocation sequence was computer-generated by this researcher using a custom MATLAB script generating 10 permuted blocks of size 6. The treatment allocation list contained 30 sham and 30 active assignments in randomized order.

The allocation sequence was not modified during the trial, and all patients were allocated according to it. On January 31, 2024, the target sex ratio for recruiting the sex-matched healthy control group of the broader DREAMS study, which is not part of the present analysis, was adjusted from 50:50 to 60:40 (female:male) to reflect the approximate sex ratio among patients.

Allocation concealment was maintained by restricting access to the allocation sequence to the independent allocation holder and allocation assignments were stored in a password-protected file.

#### 10. Blinding

Participants, research staff involved in recruitment and assessments, technical assistants delivering iTBS, clinical personnel, and clinical raters were blinded to treatment allocation. Staff delivering iTBS were not involved in ratings or clinical care, and follow-up ratings were also performed blind to allocation. The allocation list was disclosed to the study team after registration of the analysis plan (December 2024). Before registration of the analysis plan, outcome data were not analyzed.

To ensure participants and clinical staff remained blinded, active and sham treatments were administered using physically identical coils. For the sham condition, an active sham coil (pCool/aCool; MAG & More) was utilized. According to the manufacturer's specifications, the internal windings of the sham coil are positioned at the top of the coil casing. This creates a greater distance from the scalp, significantly reducing the induced magnetic field. Furthermore, the windings are arranged unidirectionally rather than opposingly, which prevents the typical summation effect at the center of the coil. Consequently, the coil induces only a weak electrical field capable of superficially stimulating scalp muscles and peripheral nerves, thereby mimicking the somatic sensation of active stimulation, but by design cannot achieve therapeutically effective cortical stimulation, even at maximum stimulator output.

Coil identity was concealed by coded labels held by an independent researcher. Operators were informed only of the coded coil-label configuration necessary for treatment delivery and were not informed whether a given label corresponded to active or sham stimulation. In all participants, the resting motor threshold was determined before the treatment phase with an active, uncoded coil; all stimulation sessions were then delivered with the coded coils.

Blinding integrity was assessed using treatment guesses, confidence ratings for treatment guesses, and tolerability ratings. Treatment guesses were collected after the 6-week follow-up assessment and before unblinding; responses were voluntary.

#### 11. Interventions

Target selection and neuronavigation: Stimulation targeted the left DLPFC using MRI-guided neuronavigation. Target selection was implemented using individualized connectivity-guided targeting based on resting-state functional connectivity with the subgenual cingulate cortex, following the method described by Cash et al. (2021). The same individualized target-selection procedure was applied in the active and sham groups.

Active iTBS: Participants allocated to active stimulation received accelerated intermittent theta-burst stimulation with an Apollo TMS system (MAG & More GmbH). Stimulation intensity was set to 90% of the individual resting motor threshold. When 90% was not tolerated, stimulation started at a lower intensity and was titrated upward, following the same procedure in both groups. Each session delivered 1,800 pulses. The schedule comprised 3 sessions per treatment day across 10 treatment weekdays, totaling 30 sessions and 54,000 pulses.

Treatment sessions lasted approximately 10 minutes. Sessions were scheduled in the afternoon, one session per hour, corresponding to 50 minutes between the end of one session and the start of the next. This reconciles the original protocol wording of 60-minute session scheduling with the manuscript wording of 50-minute intersession intervals.

Sham iTBS: Participants allocated to sham stimulation received the same schedule, target-selection procedure, session structure, and clinical workflow, but with the sham coil. The sham coil was designed to mimic acoustic and somatosensory aspects of stimulation while not producing a therapeutically effective cortical field (see Blinding).

#### 12. Concomitant care

iTBS was administered as an add-on to routine multidisciplinary inpatient care. Participants continued to receive routine clinical treatment, including pharmacotherapy, psychotherapy, occupational therapy, art therapy, physical therapy, and other ward-based therapeutic interventions as clinically indicated.

The study was designed not to prolong hospitalization and not to require changes to routine pharmacological or psychotherapeutic treatment, with one exception: during the stimulation phase, benzodiazepines and bupropion were prohibited according to rTMS safety procedures. Psychotropic medication was otherwise managed by the treating clinicians, who were blind to allocation.

#### 13. Outcomes and assessment schedule

Primary clinical outcome: Change in clinician-rated depressive symptom severity during the acute 2-week stimulation phase, measured with the Montgomery-Åsberg Depression Rating Scale (MADRS). MADRS assessments were obtained at treatment baseline, mid-treatment, and post-treatment.

Secondary clinical outcome: Change in self-reported depressive symptom severity during the acute stimulation phase, measured with the Beck Depression Inventory-II (BDI-II). BDI-II was assessed at treatment baseline, daily during the treatment phase before the first iTBS session of the day, and at post-treatment.

Follow-up: MADRS and BDI-II were reassessed at 6-week follow-up via telephone.

Exploratory categorical outcomes: Response was defined as at least 50% symptom reduction from treatment baseline to post-treatment. Remission was defined as MADRS $<10$ or BDI-II $<13$. Loss of response and loss of remission at follow-up were evaluated descriptively among participants who had met the respective criterion at post-treatment and had follow-up data available.

Safety outcomes: Adverse events and common rTMS-related side effects were assessed daily during the stimulation phase. Common side effects included headache, nausea, discomfort, fatigue, and stimulation-related unpleasantness.

Blinding outcomes: Blinding integrity was assessed using guessed treatment assignment, confidence in the treatment guess, mean perceived tolerability/pleasantness ratings across stimulation sessions, and the association between tolerability and guessed assignment.

Broader DREAMS outcomes not included in this manuscript: The broader protocol included sleep monitoring, fMRI tasks, resting-state MRI, diffusion MRI, neurocognitive tasks, interoception measures, saliva and blood biomarkers, and healthy-control comparisons.

#### 14. Participant timeline

The clinical antidepressant analysis used the following relevant assessment schedule: screening and eligibility assessment; pre-treatment/lead-in assessments; treatment baseline immediately before the first iTBS session (primary + secondary outcomes); acute treatment phase across 10 treatment weekdays; mid-treatment MADRS assessment after 1 week; post-treatment MADRS and BDI-II assessment after completion of the acute stimulation phase; and 6-week follow-up assessment.

The treatment-baseline assessment immediately before the first iTBS session served as Time 0 for the longitudinal clinical outcome models.

#### 15. Sample size

The original DREAMS ethics protocol planned a broader mechanistic study with a larger patient sample and healthy controls, powered primarily around sleep-related and mechanistic outcomes.

For the clinical antidepressant outcome analysis, the OSF preregistration specified a target analysis sample of 50 MDD patients, with 25 participants in each treatment group. To account for an expected dropout rate of approximately 15%, recruitment of 30 participants per group was planned. Recruitment ended in May 2024 for feasibility reasons after the planned analysis sample of 50 participants had been reached (51 initiated stimulation); the decision was made while allocation was concealed.

The clinical sample-size calculation was based on MADRS as the primary outcome. G*Power was used to obtain 90% power to detect a small-to-medium effect size of f=0.22 at alpha=.05. The calculation was based on an iTBS effect estimate from prior noninvasive brain stimulation literature, converted to Cohen f for repeated-measures analysis with a within-between interaction. The number of measurements was set to 3, and the correlation among repeated measures was set to 0.5. The resulting sample size estimate was 46. Because both treatment groups continued routine inpatient care, likely reducing the group difference in symptom change between active and sham, the target analysis sample was set to 50 participants.

#### 16. Data collection and management

Clinical and research data were collected by trained research and clinical personnel. Participants were assigned a study code used across study procedures. Personally identifying information and the code key were stored separately from research data. Data were handled pseudonymously and stored according to institutional data-protection procedures and the EU General Data Protection Regulation.

The code list linking participant identities to study codes was stored securely and accessible only to authorized study personnel. Data were reported and published only in anonymized form.

#### 17. Safety monitoring

rTMS was performed according to current safety standards. The ethics protocol described rTMS as a low-risk procedure, with local scalp discomfort, muscle twitching, and headache as common side effects and epileptic seizure as the most serious potential adverse event.

Treatment was conducted in rooms of the University Clinic at the Karl-Jaspers-Klinik. Emergency medication and a standardized emergency procedure were available. Participants could discontinue participation at any time. Individual discontinuation criteria included acute deterioration of mental state, discontinuation of inpatient treatment, inability to follow study procedures, or withdrawal of consent.

The ethics protocol specified no formal study-wide stopping rule. No independent data monitoring committee was established. No interim analyses were performed.

#### 18. Statistical analysis plan

Analysis set: The OSF preregistration specified a per-protocol analysis. A complementary intention-to-treat sensitivity analysis was planned only if at least 5 participants discontinued after receiving at least one iTBS session. No participant discontinued treatment after the first iTBS session; therefore, this conditional ITT sensitivity analysis was not triggered, and the per-protocol set was identical to the modified intention-to-treat population of all participants who initiated stimulation ($n$=51). A post hoc sensitivity analysis included all 57 randomized participants, with MADRS values of the 6 non-initiators multiply imputed (Supplement 2, Table S5). Acute-treatment analyses included participants who initiated iTBS and had evaluable outcome data for the respective model. Outcome-specific baseline severity criteria were applied as preregistered to avoid floor effects.

MADRS primary model: MADRS trajectories during the acute treatment phase were analyzed using latent growth models across three time points. The preregistered model sequence included a linear latent growth model, a non-linear latent growth model, a group comparison model, covariate models including age, sex, and chronic/persistent MDD tested separately, current episode duration as preregistered moderator, and a final combined model selected based on model fit, parsimony, and clinical relevance. Robust maximum likelihood estimation was used. Model fit was evaluated using AIC, BIC, sample-size adjusted BIC, CFI, and RMSEA.

BDI-II secondary model: The OSF preregistration specified latent growth models for BDI-II parallel to the MADRS sequence. Because BDI-II latent growth models showed inadequate global fit for the densely sampled daily data, BDI-II trajectories were analyzed using linear mixed-effects models. All BDI-II mixed-effects models were fitted both with participant-level random intercepts and with additional random slopes for time; both versions are reported (Supplement 2; Supplement 3, Tables S6 and S8). Maximum likelihood was used for fit indices, likelihood-ratio tests, and model comparisons, and restricted maximum likelihood for all parameter estimates.

Multiplicity: Confirmatory P values were adjusted using the Benjamini-Hochberg false discovery rate procedure. The OSF preregistration stated that directional hypotheses would be tested one-tailed and nondirectional hypotheses two-tailed. The manuscript uses more conservative two-tailed tests throughout.

Data transformations: Categorical predictors were dummy-coded. In the episode-duration models (Models 1.5 and 2.5b), treatment group was effect-coded (sham, −0.5; active, +0.5). Continuous predictors showing pronounced skewness were log-transformed. Continuous moderator variables were mean-centered before model estimation.

Missing data: Missing outcome data were handled under a missing-at-random assumption. Latent growth models used robust maximum likelihood estimation. Linear mixed-effects models used all available observations under the model likelihood.

Exploratory analyses: Exploratory analyses included multigroup latent growth models, exploratory moderator analyses, 6-week follow-up trajectory analyses, response and remission analyses, baseline-adjusted MADRS Firth logistic regression sensitivity analyses, blinding integrity analyses, and post hoc target-coordinate quality-control analyses. Further post hoc sensitivity analyses are reported in Supplement 2 (Table S5).

#### 19. Ethics and dissemination

The study was conducted in accordance with the Declaration of Helsinki and approved by the Medical Ethics Committee of the Carl von Ossietzky University Oldenburg. All participants provided written informed consent.

Results are intended for publication in a peer-reviewed journal. Deidentified individual participant data will be available upon reasonable request, subject to a data use agreement and to the restrictions of the ethics approval and applicable data protection law, beginning with publication and ending 10 years after publication (Data Sharing Statement of the manuscript).
