## Supplement 2 for "Adjunctive Accelerated Theta-Burst Stimulation for Inpatient Treatment-Resistant Depression: A Randomized Clinical Trial"

### Supplement 2: Online Supplement

### Supplementary Methods

#### Power Analysis

The sample size calculation was performed for the analysis of the Montgomery-Åsberg Depression Rating Scale (MADRS) scores as our primary outcome measure. We used the software program G*Power to conduct a power analysis. Our goal was to obtain .90 power to detect a small to medium effect size f of .22 at the standard .05 alpha error probability. We based our calculation on a meta-analysis in MDD patients reporting Standardized Mean Difference (SMD) of 0.44 for an intermittent theta burst stimulation (iTBS) effect between an active and sham group (1). We converted SMD to Cohen's f for the sample size estimation in a repeated measures ANOVA, with within-between interaction. The number of measurements was three and the correlation among repeated measures was set to 0.5. The computed total sample size was 46. As the sham group also received pharmacotherapy and psychotherapy during their inpatient stay, it is reasonable to assume that the effect size for the difference between the groups would be smaller. To account for this effect, we aim to include 50 patients in our analysis. To account for an expected drop-out of 15%, we planned to recruit 60 patients in total (i.e., 30 in each group).

#### Imaging Details and individualized neuronavigation target selection

Individualized stimulation targets were defined using a seedmap-based, cluster-guided dorsolateral prefrontal cortex (DLPFC) targeting procedure based on *Cash et al.* (2). Several clinical adaptations, including the restricted DLPFC mask, computation of three candidate clusters, cortical-surface projection of the target coordinates, and post hoc target-quality-control procedure, were further implemented.

Participants underwent structural MRI and 15 minutes of eyes-open resting-state fMRI before treatment. MRI data were acquired on a 3T Siemens Prisma scanner (Siemens AG, Erlangen, Germany) using a 64-channel head coil with a T2*-weighted echoplanar multiband sequence with a multiband acceleration factor of 4 (3). High-resolution anatomical images were acquired with a T1-weighted 3D MP-RAGE sequence. The 15-minute resting-state acquisition was selected as a clinically feasible scan duration. Cash et al. (2) reported high reproducibility of subgenual cingulate cortex (SGC)–DLPFC connectivity maps with sufficient multiband acquisition time in the approximate 15–25-minute range.

Resting-state fMRI data were preprocessed using fMRIPrep 20.2.1 and then processed using the connectivity-guided targeting pipeline. The targeting workflow included global signal regression, temporal band-pass filtering at 0.01–0.1 Hz, and minimal spatial smoothing with a 4-mm full-width-at-half-maximum kernel to preserve spatial specificity of the SGC–DLPFC connectivity maps. The preprocessed BOLD data were used to estimate individual functional connectivity between the subgenual cingulate cortex and candidate stimulation sites within the left DLPFC.

The candidate stimulation space was based on the left DLPFC region-of-interest (ROI) defined by Cash et al. (2) as the combined extent of 20-mm-radius spheres centered on BA9, BA46, the conventional 5-cm TMS site, and the Beam F3 group-average stimulation site. For the present clinical trial, this search space was restricted by excluding regions close to the temples and infero-frontal regions close to the eyes, where stimulation was expected to be less tolerable because of peripheral muscle and nerve activation. The same fixed restricted DLPFC mask was used for all participants and is shown in Figure S6.

The SGC signal was estimated using the seedmap approach rather than a conventional spherical SGC seed. In the seedmap approach, the SGC time series is estimated as a weighted spatial average of gray-matter voxels outside the DLPFC, with voxel weights derived from group-average SGC connectivity. This procedure increases signal-to-noise by using distributed gray-matter information rather than the limited number of voxels contained in a small subgenual seed. The resulting seedmap-derived SGC time series was correlated with each voxel in the restricted left DLPFC candidate mask, yielding an individual SGC–DLPFC functional connectivity map. Candidate DLPFC voxels were ranked according to negative (i.e., anticorrelated) functional connectivity with the SGC.

Target coordinates were computed using the cluster-based procedure. For each participant, a predefined proportion of the most negatively SGC-correlated DLPFC voxels was retained, spatial clustering was performed among these suprathreshold voxels using 26-voxel neighborhoods, and the center of gravity of the largest cluster was defined as the candidate target coordinate. This procedure differs from selecting the single most anticorrelated voxel and was used to improve robustness and reproducibility of individualized target selection. In the present implementation, candidate targets were computed automatically across five retained-voxel thresholds: 0.5%, 1%, 2.5%, 5%, and 10% of the most anticorrelated DLPFC voxels. Spatial convergence of candidate coordinates across thresholds was inspected, and final target selection was anchored to the 2.5% threshold. At each threshold, clusters were ranked by size among the retained anticorrelated voxels. As a clinical adaptation, the three largest candidate clusters were computed rather than only the largest cluster. The largest cluster was used whenever clinically feasible. If stimulation at this location produced excessive scalp discomfort, the target could be moved to the center of gravity of the second- or third-largest candidate cluster. The fallback decision was based solely on stimulation tolerability and was made without reference to treatment allocation or clinical outcome. Across the final sample, the first-ranked cluster was used in 41 participants. The second-ranked cluster was used in 8 participants, balanced across groups (active: n=4; sham: n=4), and the third-ranked cluster was used in 2 participants (active: n=1; sham: n=1). The final Individualized left DLPFC target locations are shown in Figure S7.

A further clinical adaptation concerned coordinate projection for neuronavigation. The cluster algorithm first returned a volumetric cortical coordinate corresponding to the cluster center of gravity. For clinical stimulation, the closest coordinate on the cortical surface was then computed and used as the final stimulation target. This surface coordinate was entered in MNI space into the MAG & More neuronavigation software. The neuronavigation software used each participant's FreeSurfer reconstruction to transform the target into individual anatomical space and guide online coil positioning. The same individualized target-selection procedure was applied for participants allocated to active and sham stimulation.

##### Post hoc quality control of neuronavigation targets

During post hoc quality control of the individualized targeting workflow, we identified a template-orientation mismatch between the fMRIPrep-derived standard-space outputs and the connectivity-guided targeting scripts. Specifically, the fMRIPrep/TemplateFlow standard-space outputs used an RAS+ orientation convention, whereas parts of the original FSL-based targeting workflow assumed the legacy FSL radiological/LAS voxel-ordering convention. Although such orientation differences are normally handled through the NIfTI header, they can affect voxel-index-based operations when downstream scripts rely on orientation assumptions. This mismatch could therefore have affected the fidelity of individualized SGC–DLPFC connectivity mapping by altering the orientation convention used for target computation.

To quantify its impact on the stimulation targets used in the trial, the targeting pipeline was rerun for all participants using the corrected image orientation. The rerun used the same restricted DLPFC mask, seedmap procedure, retained-voxel thresholds, and cluster-based target-selection procedure as the original clinical implementation.

Corrected candidate targets were recomputed across the five retained-voxel thresholds of the original procedure (0.5%, 1%, 2.5%, 5%, and 10% of the most anticorrelated DLPFC voxels). The primary quality-control metric was the Euclidean distance in millimeters between the originally selected clinical coordinate and the corresponding corrected coordinate. To examine whether this deviation was associated with antidepressant outcome, distances were correlated with acute MADRS reduction separately by treatment group (Spearman correlation).

#### Stimulation details

Stimulation was delivered using an Apollo TMS system equipped with the PowerMAG View 3D neuronavigation system (MAG & More GmbH, Munich, Germany). Treatment sessions were scheduled in the afternoon, with treatment time kept consistent within participants across treatment days. Participants received high-dose accelerated intermittent theta-burst stimulation (iTBS), consisting of 3-pulse 50-Hz bursts repeated at 5 Hz. Each 2-second train contained 30 pulses and was followed by an 8-second intertrain interval; 60 trains were delivered per session, yielding 1,800 pulses over approximately 10 minutes. Treatment comprised three daily sessions separated by 50-minute intersession intervals across 10 treatment weekdays, yielding 30 sessions and 54,000 pulses per participant. The coil was positioned using a standardized 45° orientation relative to the midsagittal line. Stimulation intensity was set to 90% of the individual resting motor threshold, which was determined using a semi-automatic electromyography-based parameter estimation by sequential testing (PEST) procedure. When 90% of the resting motor threshold was not tolerated, stimulation started at a lower intensity and was titrated upward, following the same procedure in both groups. Stimulation was added to routine inpatient treatment, which comprised pharmacotherapy, individual and group psychotherapy, art, occupational, and exercise therapy, and physiotherapy when prescribed; it was provided according to clinical need by clinicians blind to allocation.

Consistent with the inpatient implementation of the protocol, accelerated iTBS was delivered as an add-on intervention while participants continued to receive multidisciplinary inpatient care, including pharmacotherapy, individual and group psychotherapy (including psychoeducation), occupational, art, music, and exercise therapy, relaxation training, and physiotherapy when prescribed. Because screening, MRI, and target selection took place during the inpatient stay before stimulation began, the 2-week stimulation course covered only part of the stay, and participants could attend the ward program before, during, and after the stimulation phase. During the stimulation phase, benzodiazepines (4, 5) and bupropion (6) were prohibited according to rTMS safety procedures. A detailed breakdown of concomitant medications is provided in Table S2.

#### Randomization

Participants were allocated 1:1 to active or sham stimulation according to a computer-generated sequence of 10 permuted blocks of 6 (30 active and 30 sham assignments), which was held by an independent researcher. Allocation of patients followed this sequence throughout recruitment and was not modified. On January 31, 2024, the target sex ratio for recruiting the sex-matched healthy control group of the study, which is not analyzed in this report, was adjusted from 50:50 to 60:40 (female:male) to reflect the sex ratio among patients.

##### Blinding procedures

To ensure participants and clinical staff remained blinded, active and sham treatments were administered using physically identical coils. For the sham condition, an active sham coil (pCool/aCool; MAG & More) was utilized. According to the manufacturer’s specifications, the internal windings of the sham coil are positioned at the top of the coil casing. This creates a greater distance from the scalp, significantly reducing the induced magnetic field. Furthermore, the windings are arranged unidirectionally rather than opposingly, which prevents the typical summation effect at the center of the coil. Consequently, the coil induces only a weak electrical field capable of superficially stimulating scalp muscles and peripheral nerves, thereby mimicking the somatic sensation of active stimulation, but by design cannot achieve therapeutically effective cortical stimulation, even at maximum stimulator output.

To maintain double-blinding, the original coil indicators were concealed beneath encoded "A" and "B" labels. An independent researcher, who was not involved in patient care or the study otherwise, held the allocation key. To prevent associative unblinding over the course of the trial, this researcher frequently switched the label assignments and communicated only the necessary configuration to the operators.

#### Blinding Integrity Assessment

**Blinding accuracy.** Blinding integrity was first examined by comparing participants' guessed treatment assignment with actual treatment assignment among participants with available guess data. Overall guessing accuracy was summarized descriptively as the proportion of correct guesses. The association between guessed and actual treatment assignment was evaluated using a Fisher exact test, with effect size summarized as the odds ratio (OR) and 95% CI. Treatment guesses were obtained after the 6-week follow-up assessment, so as not to influence follow-up ratings, and before unblinding; responses were voluntary. In addition, the Bang blinding index (7, 8) was calculated for each group as the proportion of correct minus the proportion of incorrect guesses (range, −1 to 1; 0 indicates guessing at chance level and 1 that all participants guessed correctly), with Wald 95% CIs (7).

**Confidence and guess accuracy.** To further examine blinding integrity, confidence ratings for treatment guesses were compared between participants who correctly guessed their treatment assignment and those who guessed incorrectly. Confidence was rated on a scale from 0 to 100, with higher values indicating greater certainty. Because confidence ratings were not assumed to be normally distributed, groups were compared using the Wilcoxon rank sum test.

**Tolerability of stimulation.** As a final assessment of blinding integrity, tolerability of stimulation was compared between treatment groups using the mean pleasantness ratings across sessions of all participants, with scores ranging from 1 (extremely unpleasant) to 10 (extremely pleasant). Pleasantness ratings were compared between the active and sham groups using the Wilcoxon rank sum test.

**Tolerability and treatment guess.** To assess whether treatment tolerability influenced participants' perception of assignment, logistic regression was used to examine whether mean pleasantness ratings predicted guessed treatment assignment. Guessed treatment assignment was modeled as the dependent variable and mean pleasantness rating as the independent variable.

#### Screening, pre-treatment, and treatment baselines

Given the inpatient setting and variable durations of the ongoing hospitalization before study inclusion, symptom assessments followed a multiphase baseline approach. The initial Screening Assessment via Beck Depression Inventory-II (BDI-II) occurred upon screening. The BDI-II was utilized at screening as a supplementary clinical tool to resolve diagnostic ambiguity to decide upon study enrollment. To capture standardized descriptive clinical characteristics, a Pre-Treatment Assessment (BDI-II) was conducted on the Thursday before the first stimulation session (Monday). To describe symptom change occurring before the start of stimulation, BDI-II scores from screening and pre-treatment baseline were compared using a paired t test. This comparison was used only to characterize pre-stimulation symptom change during the inpatient lead-in period and was not treated as a treatment-effect analysis. Finally, the Treatment Baseline (MADRS and BDI-II) was assessed immediately before the first stimulation session on the first treatment day. This Treatment Baseline served as the reference point (Time 0) for all longitudinal trajectory models.

#### Effect size calculation

##### Standardization via pooled baseline variance (clinical effect sizes)

To provide clinically meaningful metrics, primary effect sizes for overall symptom change and treatment group differences were standardized using the pooled within-group standard deviation of treatment-baseline scores (MADRS, 7.56; BDI-II, 10.72), rather than the variance of change scores or latent slopes (e.g. (9)). This methodological approach evaluates treatment success on the scale of actual clinical severity observed prior to intervention. It prevents the artificial inflation of effect sizes, ensuring the metric directly reflects the clinical impact of accelerated iTBS relative to the natural fluctuation of pre-treatment depressive symptoms.

**Within-Group effect size ($d_{within}$):** This quantifies the overall cohort improvement over time, encompassing the natural course of time, placebo effects, and the active treatment.

$$d_{within} = \frac{\beta_{time} \times T}{SD_{baseline\ pooled}}$$

*Note: $\beta_{time}$ represents the unstandardized fixed-effect coefficient for overall time across the entire sample, and T the length of the acute treatment phase in model time units (2 weeks for the MADRS, 10 days for the BDI-II), so that the numerator is the model-estimated total change over this phase.*

**Between-Group effect size ($d_{between}$):** This isolates the specific therapeutic effect of the intervention by quantifying the clinical advantage of active over sham stimulation.

$$d_{between} = \frac{\Delta\beta_{active} - \Delta\beta_{sham}}{SD_{baseline\ pooled}}$$

*Note: $\beta$ represents the unstandardized model-estimated symptom change from baseline to post-treatment for each respective group.*

**Standardization via slope variance (LGM-specific effect Size)**

For the latent growth models (LGMs), an alternative effect size standardized by the standard deviation of the latent slope could be computed. This metric describes how strongly the rate of change differs between groups relative to the general variance in patient trajectories.

$$d_{slope} = \frac{\beta_{group}}{SD_{slope}}$$

We decided against reporting the effect size $d_{slope}$, as it is not recommended for clinical interpretation. The variance of longitudinal slopes is almost universally smaller than cross-sectional baseline variance, which artificially inflates the resulting effect size.

**Continuous moderation effects**

Effect sizes were intentionally omitted for the main effect of episode duration and the group-by-duration interaction. Cohen's *d* is mathematically formulated to express the standardized mean difference between discrete groups. It is not built for continuous predictors (which, in this study, were mean-centered and $log_{10}$-transformed) or their interaction terms.

Attempting to force a standardized *d* onto a continuous moderation term creates a highly unintuitive metric (i.e., standard deviations of symptom improvement per standard deviation of episode duration). Furthermore, interactions often produce deceptively small *d* values that appear clinically irrelevant, despite having massive clinical impacts when multiplied across the full range of the continuous moderator.

#### Confirmatory longitudinal model specification and model-fit evaluation

Confirmatory longitudinal analyses followed the preregistered sequential model-building strategy separately for the clinician-rated primary outcome (MADRS) and the self-reported secondary outcome (BDI-II). The treatment baseline assessment immediately before the first stimulation session served as Time 0 for all longitudinal treatment-phase models.

All analyses were performed in R (version 4.3.1) (10). Latent growth models were estimated with the lavaan package (version 0.6-21) (11), and linear mixed-effects models with lme4 (version 1.1-35.5) (12), with Satterthwaite degrees of freedom obtained from lmerTest (version 3.1-3) (13).

For MADRS, confirmatory analyses were conducted using latent growth models (LGMs) across the three treatment-phase assessments. The model sequence first compared alternative time-function specifications, including a linear latent growth model and a non-linear latent growth model in which the loading for the third measurement occasion was freely estimated. Treatment group was then added to estimate the latent baseline and slope as contrasted between the two groups. Because the treatment groups differed in baseline MADRS severity, an additional model adjusted the latent slope for baseline severity, by regressing the slope onto the intercept factor. Prespecified covariates were then examined separately by adding age, sex, and chronic MDD status to the latent baseline and slope factors. Current episode duration was tested as the preregistered moderator by adding duration and the duration × treatment-group interaction as predictors of the latent baseline and slope factors. In this model, treatment group was effect-coded (sham, −0.5; active, +0.5), so that the duration effect represents the mean association across both groups. The final model was selected based on model fit, parsimony, preregistered clinical relevance, and information criteria. Time was coded in weeks for the MADRS LGMs.

For BDI-II, preregistered latent growth models were first evaluated using the same sequential logic. However, the high-granular daily BDI-II latent growth models showed inadequate global fit across the confirmatory specifications and were therefore not used for interpretation. Because the BDI-II data included a higher number of repeated assessments during the acute treatment phase, the final confirmatory BDI-II analyses were conducted with linear mixed-effects models (LMMs) instead (deviation from the preregistration), which are less strict in terms of fit and more parsimonious compared to LGMs (14, 15). This is of advantage especially given the relatively small sample size. The LMM sequence followed the same conceptual order as the MADRS models: time-function specification, treatment-group contrast, prespecified covariates, current episode duration as preregistered moderator, and final model selection. As for MADRS, treatment group was effect-coded in the episode-duration model. Time was coded in days for the BDI-II LMMs. Each BDI-II model was fitted in two versions that are both reported: with a random intercept and with a random intercept and a correlated random slope for time, which allows participants to differ in their rate of change. An uncorrelated random-effects structure was specified as fallback for singular or nonconverging fits and was not needed. The slope variance was tested against the random-intercept version with a likelihood-ratio test referred to a 50:50 mixture of $\chi^2$ distributions with 1 and 2 degrees of freedom. Table 2 in the main text shows the random-slope version; fit indices and parameter estimates of both versions are given in Supplement 3 (Tables S6 and S8).

For LGMs, model fit was evaluated using relative and absolute fit indices. Akaike Information Criterion (AIC), Bayesian Information Criterion (BIC), and sample-size adjusted BIC (sBIC) were used as relative fit indices, with lower values indicating better relative model fit among competing models. The Comparative Fit Index (CFI) and Root Mean Square Error of Approximation (RMSEA) were used as global fit indices. CFI values of 0.95 or higher and RMSEA values of 0.06 or lower were interpreted as indicating good global fit, whereas CFI values of 0.9 to 0.95 and RMSEA values up to 0.08 were considered acceptable. Models with CFI values below 0.9 and RMSEA values above 0.08 were considered of inadequate global fit. The final model selection also considered parsimony, and fit to the preregistered analytic sequence.

For LMMs, model fit was summarized using AIC, BIC, and log-likelihood. Lower AIC and BIC values and higher log-likelihood values indicated better relative model fit among competing models. No absolute good-fit or poor-fit thresholds were applied to LMMs, because AIC, BIC, and log-likelihood are comparative rather than absolute fit measures.

Missing data were handled under missing-at-random assumptions using robust maximum likelihood estimation for LGMs. All missing MADRS ratings at weeks 0, 1, and 2 resulted from rating forms that were lost after the interview had been conducted and were thus unrelated to participants' clinical state. For LMMs, fit indices, likelihood-ratio tests, and model comparisons were based on maximum likelihood and all parameter estimates on restricted maximum likelihood; fixed effects were tested with Satterthwaite degrees of freedom, with 95% CIs from the corresponding t distribution.

#### Exploratory model specifications: Follow-up analysis

The 6-week follow-up analysis was preregistered as exploratory. The preregistered approach first attempted to extend the acute-phase latent growth model to include the follow-up assessment using time scores 0, 1, 2, and 8. This model showed poor fit for MADRS trajectories (CFI = 0.850, RMSEA = 0.192), indicating that a single linear growth process across the acute stimulation phase and the longer post-treatment interval was not appropriate. Follow-up attrition further reduced the stability of the extended LGM. The preregistered fallback analysis was a simple group comparison at follow-up. However, because treatment groups differed in baseline MADRS severity, an endpoint-only comparison was considered statistically inappropriate, as it would conflate follow-up group differences with pre-existing baseline differences.

We therefore used an exploratory piecewise linear mixed model with a knot at Week 2, corresponding to the end of stimulation. The model estimated separate slopes for the acute stimulation phase and the post-treatment follow-up phase, included fixed effects for group, phase-specific time terms, and their interactions, and included a random intercept for participant. Covariates retained in the final acute-phase models were not carried forward, so that the follow-up model parallels the confirmatory group model; a version of the MADRS model adjusted for sex and chronic MDD is reported as a sensitivity analysis (Table S5). This approach used all available longitudinal data and modeled the structural break between active treatment and follow-up. Results from this model should be interpreted as exploratory and as addressing whether the acute group difference showed evidence of rapid post-treatment loss, not as a definitive test of long-term durability.

#### Exploratory categorical outcome analyses: response, remission, and follow-up status

##### Exploratory post-treatment response and remission analyses

Categorical clinical outcomes were evaluated at post-treatment for both MADRS and BDI-II. Response was defined as a reduction of at least 50% from treatment baseline to post-treatment. Remission was defined as a post-treatment score below the scale-specific remission threshold, using MADRS < 10 and BDI-II < 13. Analyses included participants with endpoint-specific evaluable baseline and post-treatment data.

Between-group differences were evaluated using Fisher exact tests when a cell count was below 5 and Pearson $\chi^2$ tests otherwise. ORs and 95% confidence intervals (CIs) were reported as effect-size estimates. These categorical analyses were exploratory and were interpreted alongside the prespecified continuous longitudinal trajectory analyses.

##### Exploratory baseline-adjusted MADRS logistic regression sensitivity analyses

Because active and sham groups differed in treatment-baseline MADRS severity, non-preregistered sensitivity analyses were conducted to examine whether categorical MADRS response and remission findings were robust to baseline adjustment. These analyses were restricted to MADRS outcomes because no corresponding baseline group difference was observed for BDI-II.

Baseline-adjusted models included treatment group and treatment-baseline MADRS (linear term) and were specified before model fitting (not preregistered); the functional form of baseline severity was not selected from the data. Because events were few, models were estimated by Firth penalized-likelihood logistic regression (R package logistf), which reduces the small-sample bias of maximum-likelihood estimates; 95% CIs and *P* values were obtained from the penalized profile likelihood. Absolute risk differences (active minus sham) with Newcombe hybrid score 95% CIs were calculated for all four categorical outcomes. These analyses were exploratory and did not replace the prespecified continuous longitudinal trajectory analyses.

##### Exploratory loss-of-status analyses at 6-week follow-up

To descriptively evaluate whether categorical clinical improvements were maintained at the 6-week follow-up, loss of response and loss of remission were assessed among participants who had met the respective criterion at post-treatment. Formal relapse terminology was avoided because a single follow-up assessment at 6 weeks does not support stricter relapse definitions requiring sustained symptom return or diagnostic verification.

Loss of response was defined as no longer showing a reduction of at least 50% relative to the original treatment-baseline score. Loss of remission was defined as crossing back above the scale-specific remission threshold at follow-up, using MADRS ≥ 10 and BDI-II ≥ 13. Because the post-treatment responder and remitter subgroups were small, no formal inferential tests were performed. To avoid misrepresenting missing follow-up data, loss rates were calculated using a restricted denominator comprising only participants with available follow-up data within the respective post-treatment responder or remitter subgroup. Participants with missing follow-up data were not assumed to have either maintained or lost their post-treatment clinical status.

#### Exploratory model specifications: multigroup and moderator analyses

Exploratory multigroup and moderator analyses followed the same outcome-specific modeling frameworks, time coding, estimation settings, missing-data assumptions, and model-fit evaluation criteria described above for the confirmatory longitudinal analyses.

First, preregistered exploratory multigroup analyses were conducted to examine whether selected latent growth parameters differed between treatment groups. For MADRS, multigroup LGMs compared constrained and unconstrained specifications for the intercept variance, the slope variance, and the intercept–slope covariance. For each parameter, the unconstrained model allowed the parameter to vary between the active and sham groups, whereas the constrained model held the parameter equal across groups. Model fit was evaluated descriptively using the LGM fit criteria described above.

For BDI-II, the corresponding latent growth models showed inadequate global fit and were therefore not interpreted. Because the preregistered multigroup constraints referred specifically to latent intercept and slope factors, they were not directly transferrable to the final BDI-II mixed-effects model framework without specifying additional exploratory random-effects variance–covariance structures.

Second, exploratory moderator analyses examined whether predefined baseline variables modified treatment-related symptom trajectories or were associated with baseline depression severity and symptom reduction irrespective of treatment assignment. These moderators were grouped according to the preregistered domains. Psychological moderators comprised comorbid personality disorder status, assessed by trained personnel using the Structured Clinical Interview for DSM-5 Personality Disorders (SCID-5-PD) and subsequently cross-referenced with patient medical records, and childhood trauma severity assessed with the Childhood Trauma Questionnaire (CTQ). Social-integration moderators comprised relationship status, social network size, and loneliness assessed with the UCLA loneliness scale. Employment status was examined as the demographic moderator. Each moderator was tested in a separate model to avoid overparameterization given the sample size.

For MADRS, exploratory moderator analyses were conducted using the same LGM framework as the confirmatory MADRS analyses. In each model, the latent baseline factor and latent slope factor were regressed on the treatment-group contrast, the respective moderator, and the treatment group × moderator interaction. The treatment group × moderator effect on the latent slope was the primary parameter for evaluating whether the active-vs-sham difference in acute MADRS change varied by moderator. The moderator main effect on the latent baseline factor was used to evaluate whether the moderator was associated with baseline depression severity. The moderator main effect on the latent slope was used to evaluate whether the moderator was associated with symptom reduction in the sham group. The treatment group × moderator effect on the latent baseline factor was included to account for possible baseline differences across moderator levels and is reported descriptively. Because group was dummy-coded (sham, 0; active, 1), moderator main effects on the latent baseline and slope factors refer to the sham group.

For BDI-II, exploratory moderation analyses were conducted using the same LMM framework as the confirmatory BDI-II analyses, in both random-effects versions. Each model included fixed effects for time, treatment group, the respective moderator, all two-way interactions among these terms, and the time × treatment group × moderator interaction. The three-way interaction was the primary parameter for evaluating whether the active-vs-sham difference in daily BDI-II change varied by moderator. Random intercepts, and in the second version correlated random slopes for time, were included at the participant level. As for MADRS, moderator main effects and time × moderator effects refer to the sham group. Model fit was evaluated using AIC, BIC, and log-likelihood, as described above.

Continuous moderator variables were mean-centered before model estimation. When continuous variables showed pronounced skewness, log-transformed values were used before mean-centering. Binary moderators were dummy-coded. Because these analyses were exploratory and were not powered for strong subgroup inference, moderation findings were interpreted as exploratory and hypothesis-generating.

#### Post hoc sensitivity analyses

**General.** The analyses in this section were not preregistered. They are reported regardless of their results and were not adjusted for multiplicity.

**Baseline adjustment of the acute effect.** Model 1.3b regressed the latent slope on group and on the latent baseline (Table S7 in Supplement 3). As cross-checks, the week-2 group difference was estimated by analysis of covariance (ANCOVA; week-2 MADRS on baseline MADRS and group, complete cases) and by the mixed model for repeated measures described below. The unadjusted effect of Model 1.3 was decomposed into the baseline-adjusted effect and the part attributable to the latent baseline difference (product of coefficients; 95% CIs by the delta method and by percentile bootstrap with 2000 resamples, also restricted to resamples with admissible solutions).

**All randomized participants.** To include the 6 randomized participants who did not start stimulation, their MADRS values at weeks 0, 1, and 2 were multiply imputed ($m = 50$; predictive mean matching; R package mice) from screening and pre-treatment BDI-II scores, sex, and age. Imputation was performed separately per arm under a missing-at-random assumption and, as the main variant, by reference-based imputation in which non-initiators of the active arm were imputed from the sham arm, because they never received stimulation. Each imputed data set was analyzed with Models 1.3 and 1.3b, and estimates were pooled with Rubin's rules using Barnard–Rubin degrees of freedom. In a tipping-point analysis, the imputed week-2 MADRS values were shifted against the active group ($+\delta$ in active and $-\delta$ in sham non-initiators; $\delta/2$ at week 1) until $P \geq .05$.

**Follow-up.** The piecewise linear mixed model described above remained the primary follow-up analysis. The baseline-adjusted group difference at week 8 was obtained from the same model with the group difference at baseline constrained to zero (constrained longitudinal data analysis). Sensitivity analyses were (1) a mixed model for repeated measures with visit as a categorical factor, an unstructured covariance matrix, and no group term at baseline (Kenward–Roger degrees of freedom; R package mmrm); (2) ANCOVA of week-8 MADRS on baseline MADRS and group in participants with week-8 data; (3) the same ANCOVA after multiple imputation of missing week-8 values within arm ($m = 50$; predictors: MADRS at weeks 0–2, sex, chronic MDD); (4) single imputation of missing week-8 values by the week-2 value (last observation carried forward, LOCF) or by the baseline value (baseline observation carried forward, BOCF); and (5) a two-dimensional tipping-point analysis in which the imputed week-8 values of participants without follow-up were shifted by −10 to +10 MADRS points, separately per arm. Analyses (1) to (5) adjust for baseline MADRS only; the piecewise model was additionally fitted with sex and chronic MDD as covariates.

**Medication changes during stimulation.** Model 1.3 was extended by two indicators of medication changes during stimulation, (a) initiation, discontinuation, or switch and (b) dose adjustment, as predictors of intercept and slope. Because these changes occurred after randomization, this analysis is a sensitivity analysis only.

**MADRS raters.** The distribution of the raters across groups was compared at each visit with Fisher exact tests. Models 1.3 and 1.3b were refitted with the rater at each visit as a time-varying covariate (rater effects constrained to be equal across visits); because only three raters were involved, raters were modelled as fixed effects. As checks, a linear mixed model with rater as fixed effect, Model 1.3 in participants rated by the same rater at weeks 0 and 2, and the piecewise follow-up models with rater were fitted.

**BDI-II: further checks. The confirmatory P values of both versions were adjusted** with the Benjamini–Hochberg procedure as in the main analysis, separately for both versions (random-slope version in Table 2 and random-intercept version in the Results of the main text). As further checks of the time × group effect, the random-intercept model was refitted with a continuous-time first-order autoregressive residual structure (CAR(1)), with cluster-robust bias-reduced (CR2) standard errors, and with Kenward–Roger degrees of freedom; day-10 BDI-II scores were compared between groups by ANCOVA adjusted for day 0; and the model was refitted including the participant excluded for a baseline BDI-II score below 14.

#### Hypotheses

The confirmatory hypotheses were preregistered (doi:10.17605/OSF.IO/6EMSU) before analysis. Hypothesis H1 tested overall acute symptom change during the stimulation phase, H2 whether acute symptom change differed between active and sham iTBS, H3a whether current episode duration was associated with overall symptom change, and H3b whether it moderated the active–sham difference; each hypothesis was tested for MADRS and BDI-II (Table 2 of the main text, footnote b; OSF numbering 1.1/1.2, 2.1/2.2, 3.1a/3.2a, and 3.1b/3.2b for MADRS/BDI-II). Confirmatory P values were adjusted with the Benjamini–Hochberg procedure within each outcome across these four tests (H1, H2, H3a, H3b); for the BDI-II, this was done separately for the random-intercept and random-slope versions. Exploratory analyses were not part of this confirmatory hypothesis set and are reported separately.

### Supplementary Results

#### Comorbid psychiatric diagnoses

Comorbid psychiatric diagnoses were frequent in both treatment groups (Table S1). Overall, 42 of 51 participants (82%) had at least one comorbid psychiatric diagnosis, with comparable proportions in the sham and active groups (19 of 24 [79%] vs 23 of 27 [85%]; OR = 1.50, *P* = .72). Similarly, the total number of comorbid diagnoses was comparable between groups (median [IQR], sham: 2 [2.25]; active: 2 [2.50]; W = 307.5, *P* = .76).

Anxiety disorders were the most common comorbidity category (26 of 51 [51%]), followed by personality disorders (17 of 51 [33%]). There was no evidence of between-group differences in overall comorbidity burden or in any specific comorbid diagnosis, including anxiety disorders, personality disorders, posttraumatic stress disorder, obsessive-compulsive disorder, Attention-Deficit/Hyperactivity Disorder (ADHD)/Attention-Deficit Disorder (ADD), eating disorders, somatoform disorder, dissociative disorder, or autism (all Fisher exact test *P* > .05).

#### Concomitant psychotropic medication

Concomitant psychotropic medications are summarized in Table S2. Medication patterns were comparable between treatment groups: no antidepressant class (all P ≥ .25) and no augmentation or adjunctive medication class (all P ≥ .10) differed between the sham and active groups. As expected in routine inpatient care, medication changes were frequent and similarly distributed. Between hospital admission and the end of the stimulation phase, 19 of 24 sham (79%) and 24 of 27 active participants (89%) had at least one change (P = .45). During the stimulation phase, a psychotropic medication was initiated, discontinued, added, or switched in 9 of 24 (38%) and 11 of 27 participants (41%; P > .99), and doses were adjusted in 15 of 24 (63%) and 21 of 27 participants (78%; P = .36). Because changes during stimulation occurred after randomization and could influence symptom change, the MADRS group comparison model was additionally adjusted for both types of change (Post hoc sensitivity analyses): the treatment effect decreased by about 12% and remained significant (−3.11 points/week; 95% CI, −5.23 to −1.00; P = .004), and neither indicator was significantly associated with the rate of change.

#### Confirmatory longitudinal model results

##### MADRS latent growth models

The confirmatory MADRS latent growth models showed good or in one case acceptable (Model 1.4b) global fit across the preregistered model sequence. The initial linear latent growth model showed good fit according to CFI and RMSEA criteria (CFI = 1.000; RMSEA = 0.000; AIC = 985.9; BIC = 1001.3). The non-linear latent growth model also showed good global fit (CFI = 1.000; RMSEA = 0.000), but did not improve relative fit compared with the more parsimonious linear model, as indicated by slightly higher AIC and BIC values (AIC = 987.4; BIC = 1004.8). The estimated final slope loading in the non-linear model was 2.21 (95% CI, 1.57 to 2.85), close to the linear coding of the final assessment at Week 2. Therefore, the linear time specification was retained for subsequent MADRS models.

In the linear model, MADRS scores decreased significantly across the full cohort, with an estimated slope of −4.40 points per week (95% CI, −5.53 to −3.28; *P* < .001), corresponding to a model-estimated reduction of −8.81 points over the 2-week acute treatment phase. The latent slope variance was significant, indicating interindividual variability in rates of symptom change (estimate, 13.26; 95% CI, 3.10 to 23.42; *P* = .01).

Adding treatment group improved relative model fit compared with the unconditional linear model (AIC = 977.9 vs 985.9; BIC = 997.2 vs 1001.3). In the group comparison model, active treatment allocation was associated with higher baseline MADRS severity (β = 5.10; 95% CI, 1.00 to 9.20; *P* = .01) and a significantly steeper reduction in MADRS scores over time (β = −3.54 points/week; 95% CI, −5.53 to −1.55; *P* < .001). This corresponds to an additional model-estimated improvement of −7.08 MADRS points over the acute 2-week treatment phase in the active group compared with sham (Figure S1).

Because baseline MADRS severity differed between treatment groups, baseline severity was additionally entered as a sensitivity covariate predicting the latent slope. Baseline severity was not significantly associated with subsequent MADRS change (β = −0.12; 95% CI, −0.32 to 0.08; *P* = .24), and the treatment effect on the slope remained significant (β = −2.92; 95% CI, −4.99 to −0.85; *P* = .006; −5.84 points over 2 weeks; Cohen's d = −0.77). Model 1.3b replaces the covariance between latent baseline and slope by a regression and therefore has the same number of parameters and the same fit as Model 1.3; the two models differ only in whether the treatment effect on the slope is conditional on baseline severity.

The prespecified covariate models showed no evidence that age influenced either baseline MADRS severity or acute symptom change. Sex was associated with baseline severity, with male participants showing lower baseline MADRS scores (β = −4.66; 95% CI, −8.10 to −1.21; *P* = .008), but sex was not associated with the slope of symptom change. Chronic MDD was associated with higher baseline MADRS severity (β = 5.44; 95% CI, 0.04 to 10.85; *P* = .048), but was not associated with the slope of symptom change.

The preregistered episode-duration moderation model did not indicate that current episode duration influenced acute MADRS change or moderated the treatment effect. Duration was not significantly associated with the latent slope (β = 1.33; 95% CI, −1.07 to 3.73; *P* = .28), and the duration × treatment group interaction on the slope was not significant (β = 2.94; 95% CI, −1.87 to 7.76; *P* = .23).

The final combined MADRS model showed the best relative fit among the confirmatory MADRS specifications based on AIC, BIC, and sBIC (AIC = 971.8; BIC = 995.0; sBIC = 957.3) and retained good global fit (CFI = 1.000; RMSEA = 0.000). In this final model, active treatment allocation remained the only significant predictor of acute MADRS change (β = −3.60; 95% CI, −5.60 to −1.60; *P* < .001). Chronic MDD and sex remained as predictors of baseline severity, with higher baseline MADRS scores in patients with chronic MDD and lower baseline scores in male participants. The LGM model plot is provided in Figure S2.

**BDI-II model selection and linear mixed-effects models**

Before fitting the BDI-II longitudinal models, one participant was excluded from the BDI-II analysis set because their baseline BDI-II score fell below the predefined minimum-severity threshold for longitudinal outcome analyses (BDI-II < 14) and was excluded from all subsequent analyses of BDI-II scores. The preregistered BDI-II latent growth models showed consistently poor global fit across the confirmatory model sequence. CFI values ranged from 0.818 to 0.850 and RMSEA values ranged from 0.206 to 0.279, exceeding the predefined threshold for adequate fit. Consequently, the BDI-II latent growth model estimates were not interpreted, and BDI-II trajectories were analyzed using linear mixed-effects models with a random intercept and with a random intercept and slope, which accommodate the daily repeated assessments during the acute treatment phase (fit indices and parameter estimates of all models in Tables S6 and S8 in Supplement 3).

Within the LMM framework, the simple linear time model showed a significant overall decrease in BDI-II scores across the acute treatment phase (−0.31 points/day; random intercept, 95% CI, −0.40 to −0.21; P < .001; random slope, 95% CI, −0.47 to −0.14; P < .001). Adding a random slope for time improved the fit of every BDI-II model (likelihood-ratio tests, all P < .001; AIC lower by 56 to 68 points). The SD of individual slopes (0.52 points/day in Model 2.3) was more than twice the estimated group difference, day-to-day fluctuation around each participant's own trajectory was moderate (residual SD, 3.0 points), and individual slopes were estimated with a mean reliability of 0.77. A quadratic model yielded only a minimal reduction in AIC relative to the linear model (random intercept, 3150.8 vs 3151.3; random slope, 3082.9 vs 3084.2), while BIC favored the simpler linear model in both versions (3172.4 vs 3168.5 and 3113.1 vs 3110.0). The quadratic term was not significant (−0.03 points/day$^2$; random intercept, P = .12; random slope, P = .07). Therefore, the linear time specification was retained for the confirmatory BDI-II LMM sequence.

Adding treatment group showed that baseline BDI-II scores did not differ between groups (0.08 points; random intercept, 95% CI, −6.02 to 6.19; random slope, 95% CI, −6.11 to 6.28; P = .98 for both). Active treatment was associated with a steeper daily reduction in BDI-II scores than sham (time × group, −0.23 points/day), which was significant with a random intercept (95% CI, −0.41 to −0.05; P = .01) but not with a random slope (95% CI, −0.57 to 0.10; P = .17). This corresponds to an additional model-estimated improvement of −2.33 BDI-II points over the 10-day acute treatment phase in the active group compared with sham.

The prespecified covariate models showed that age was not associated with baseline BDI-II severity or daily symptom change in either version. Male sex was associated with lower baseline BDI-II severity (−7.70 points; random intercept, 95% CI, −13.50 to −1.90; P = .01; random slope, 95% CI, −13.62 to −1.79; P = .01) but not with the rate of daily symptom change. Chronic MDD was not significantly associated with baseline BDI-II severity (5.75 points; P = .06 in both versions); it was associated with a steeper daily reduction in BDI-II symptoms with a random intercept (time × chronic MDD, −0.24 points/day; 95% CI, −0.42 to −0.06; P = .009) but not with a random slope (95% CI, −0.58 to 0.09; P = .15).

The preregistered episode-duration moderation model did not indicate that current episode duration influenced acute BDI-II change or moderated the treatment effect in either version. Episode duration was not associated with daily symptom change (time × episode duration, 0.20; random intercept, 95% CI, −0.05 to 0.45; P = .12; random slope, 95% CI, −0.27 to 0.67; P = .40), and the time × group × episode duration interaction was not significant (0.42; random intercept, 95% CI, −0.08 to 0.91; P = .10; random slope, 95% CI, −0.52 to 1.35; P = .38).

Among the confirmatory LMMs, the final combined model had the lowest AIC and the highest log-likelihood in both versions (random intercept, AIC = 3139.0, log-likelihood = −1560.5; random slope, AIC = 3080.9, log-likelihood = −1529.4), although BIC favored the simple linear model. In the final combined model, male sex remained associated with lower baseline BDI-II severity (random intercept, −7.23 points; 95% CI, −13.05 to −1.41; P = .02; random slope, −7.13 points; 95% CI, −12.94 to −1.33; P = .02). The time × group term (−0.23 points/day) was significant with a random intercept (95% CI, −0.41 to −0.05; P = .01) but not with a random slope (95% CI, −0.56 to 0.10; P = .17), and the same applied to the time × chronic MDD term (−0.24 points/day; random intercept, 95% CI, −0.42 to −0.06; P = .009; random slope, 95% CI, −0.58 to 0.09; P = .15).

#### Exploratory follow-up analysis

In the piecewise models (Table 2 in the main text; Tables S7 and S8 in Supplement 3), MADRS scores changed little between week 2 and the 6-week follow-up (sham, −0.66 points; 95% CI, −3.60 to 2.27; active, 1.68 points; 95% CI, −1.15 to 4.51), and the rate of change did not differ between groups (0.39 points/week; 95% CI, −0.29 to 1.07; *P* = .26; Cohen’s *d* = 0.31). BDI-II scores continued to decrease in both groups (random-slope model: sham, −3.47 points; 95% CI, −5.21 to −1.73; active, −2.96 points; 95% CI, −4.55 to −1.37), again without a group difference in the rate of change (0.01 points/day; 95% CI, −0.04 to 0.07; *P* = .67; Cohen’s *d* = 0.05; random-intercept model, *P* = .72). Observed mean week-8 MADRS scores were 18.5 in both groups (sham, n = 21; active, n = 23). Because the active group started from a higher baseline, the week-8 difference was also estimated with the group difference at baseline constrained to zero; it favored active stimulation but was not significant (−3.01 points; 95% CI, −6.82 to 0.80; *P* = .12). Follow-up MADRS data were missing for 7 participants (sham, 3; active, 4), 6 of whom had responded at week 2. Further baseline-adjusted analyses and analyses addressing missing follow-up data gave consistent results (Post hoc sensitivity analyses; Table S5).

##### Exploratory loss-of-status analyses at 6-week follow-up

Maintenance of post-treatment categorical status at the 6-week follow-up was evaluated descriptively. For MADRS response, 3 sham participants and 11 active-treatment participants had met response criteria at post-treatment. Among those with available follow-up data, 1 of 1 evaluated sham responder (100.0%) and 1 of 7 evaluated active-treatment responders (14.3%) no longer met response criteria at follow-up. For MADRS remission, 3 sham participants and 7 active-treatment participants had met remission criteria at post-treatment. Among those with available follow-up data, 0 of 1 evaluated sham remitters (0.0%) and 1 of 4 evaluated active-treatment remitters (25.0%) crossed back above the remission threshold.

For BDI-II response, 1 sham participant and 4 active-treatment participants had met response criteria at post-treatment. Among those with available follow-up data, 0 of 1 evaluated sham responders (0.0%) and 0 of 3 evaluated active-treatment responders (0.0%) no longer met response criteria at follow-up. For BDI-II remission, 5 sham participants and 7 active-treatment participants had met remission criteria at post-treatment. Among those with available follow-up data, 0 of 3 evaluated sham remitters (0.0%) and 1 of 6 evaluated active-treatment remitters (16.7%) crossed back above the remission threshold.

These descriptive estimates suggest that most evaluated participants who had achieved post-treatment response or remission maintained their categorical status at the 6-week follow-up. However, interpretation is limited by the small number of post-treatment responders and remitters with available follow-up data. Overall follow-up missingness for MADRS was 12.5% in the sham group (3 of 24) and 14.8% in the active group (4 of 27). For BDI-II, follow-up missingness was 13.0% in the sham group and 11.1% in the active group. Missingness was particularly relevant among participants who had achieved post-treatment clinical milestones; for example, in the active group, 4 MADRS responders and 3 MADRS remitters did not have follow-up MADRS data. Therefore, the calculated loss-of-status percentages among evaluated participants may not fully represent maintenance of clinical improvement in the full post-treatment responder/remitter sample.

#### Exploratory categorical outcome analyses: response, remission, and follow-up status

##### Exploratory post-treatment response and remission analyses

Post-treatment categorical outcomes are summarized in Table S3. MADRS response was observed in 3 sham participants (13.0%) and 11 active-treatment participants (42.3%), corresponding to significantly higher odds of response in the active group (OR = 4.73; 95% CI, 1.01 to 31.09; *P* = .03). MADRS remission occurred in 3 sham participants (12.5%) and 7 active-treatment participants (26.9%), but the between-group difference was not statistically significant in the unadjusted categorical analysis (OR = 2.53; 95% CI, 0.49 to 17.36; *P* = .29).

For BDI-II, response was observed in 1 sham participant (4.5%) and 4 active-treatment participants (14.8%; OR = 3.57; 95% CI, 0.32 to 188.07; *P* = .36). BDI-II remission was observed in 5 sham participants (22.7%) and 7 active-treatment participants (25.9%; OR = 1.19; 95% CI, 0.27 to 5.68; *P* = .80). Thus, unadjusted categorical analyses suggested higher MADRS response in the active group, whereas MADRS remission and both BDI-II categorical outcomes were not significantly different between groups. These categorical results should be interpreted cautiously because dichotomization reduces information and event counts were modest.

##### Exploratory baseline-adjusted MADRS logistic regression sensitivity analyses

Risk differences and baseline-adjusted odds ratios are also given in Table S3. The MADRS response rate was 29.3 percentage points higher with active stimulation (95% CI, 3.9 to 49.8; number needed to treat, 3.4; 95% CI, 2.0 to 25.9); the differences for MADRS remission (14.4; 95% CI, −8.3 to 35.2), BDI-II response (10.3; 95% CI, −9.1 to 28.3), and BDI-II remission (3.2; 95% CI, −21.1 to 25.8) were smaller, with confidence intervals including zero. After adjustment for baseline MADRS severity in Firth logistic regression, active treatment was associated with higher odds of MADRS response (OR = 8.53; 95% CI, 1.88 to 57.40; $\chi^2(1) = 8.22$; P = .004) and MADRS remission (OR = 5.73; 95% CI, 1.11 to 44.73; $\chi^2(1) = 4.41$; P = .04). The corresponding unadjusted Firth estimates were 4.35 (95% CI, 1.19 to 19.53) and 2.36 (95% CI, 0.61 to 10.90).

The categorical MADRS treatment effect was therefore not explained by the higher treatment-baseline MADRS severity in the active group. The larger adjusted odds ratios partly reflect the non-collapsibility of odds ratios when a strong prognostic covariate is added; together with the wide confidence intervals, which reflect the small number of events, this supports interpreting these estimates as exploratory.

#### Exploratory multigroup and moderator model results

##### Exploratory MADRS multigroup models

The exploratory MADRS multigroup latent growth models showed good global fit across the constrained and unconstrained specifications, with CFI values of 1.000 and RMSEA values of 0.000 for Models 1.7 to 1.9 (Table S6 in Supplement 3). However, several of these exploratory multigroup models yielded negative residual variance estimates, consistent with Heywood cases; therefore, variance and covariance estimates from these models were interpreted cautiously.

For the intercept–slope covariance, the constrained model did not fit significantly worse than the unconstrained model, Satorra-Bentler scaled $\chi^2$ difference test: $\chi^2$diff = 0.47, df = 1, *P* = .49. Relative fit indices also favored the more parsimonious constrained model over the unconstrained model (AIC = 977.8 vs 979.3; BIC = 1006.7 vs 1010.2). Thus, there was no statistical evidence that the intercept–slope covariance differed between treatment groups. Descriptively, the unconstrained model suggested different intercept–slope correlations between groups (sham: *r* = −0.48, active: *r* = −0.10); however, this pattern should be interpreted cautiously given the non-significant model comparison and the presence of Heywood cases.

For the intercept variance, constraining the variance to equality across groups did not significantly worsen model fit, $\chi^2_{diff}$ = 0.87, df = 1, *P* = .35. The constrained model also showed slightly lower AIC and BIC values than the unconstrained model (AIC = 977.9 vs 979.3; BIC = 1006.9 vs 1010.2), providing no evidence for group-specific intercept variance differences.

Similarly, constraining the slope variance to equality across groups did not significantly worsen model fit, $\chi^2$diff = 0.37, df = 1, *P* = .54. The constrained model again showed slightly lower AIC and BIC values than the unconstrained model (AIC = 977.6 vs 979.3; BIC = 1006.6 vs 1010.2). Thus, the exploratory multigroup analyses did not provide evidence that variability in acute MADRS change differed between active and sham treatment groups.

**Exploratory MADRS moderator models**

The exploratory MADRS moderator models generally showed good or at least acceptable global fit. Most models had CFI values close to or equal to 1.000 and RMSEA values of 0.000. The childhood-trauma model showed acceptable rather than good RMSEA-based fit (CFI = 0.988; RMSEA = 0.077), and the social-network model showed good global fit but yielded a negative residual variance estimate consistent with a Heywood case.

None of the exploratory moderators significantly modified the active-vs-sham difference in acute MADRS change. The treatment group × moderator effects on the MADRS slope were non-significant for personality disorder status (β = 1.99; 95% CI, −2.04 to 6.01; *P* = .33), childhood trauma (β = 4.62; 95% CI, −12.58 to 21.82; *P* = .60), relationship status (β = −2.57; 95% CI, −6.77 to 1.63; *P* = .23), social network size (β = −0.78; 95% CI, −8.50 to 6.94; *P* = .84), loneliness (β = −0.06; 95% CI, −0.27 to 0.16; *P* = .61), and employment status (β = −1.51; 95% CI, −5.45 to 2.43; *P* = .45).

Several exploratory moderators were associated with baseline MADRS severity or overall symptom change irrespective of treatment-specific moderation (main effects in the sham group, the reference category). Personality disorder status was associated with higher baseline MADRS severity (β = 6.18; 95% CI, 1.44 to 10.92; *P* = .01), as was childhood trauma severity (β = 32.91; 95% CI, 4.73 to 61.10; P = .02) and being in a relationship (β = 6.78; 95% CI, 1.20 to 12.36; *P* = .02). Employment status was associated with lower baseline MADRS severity (β = −9.19; 95% CI, −14.14 to −4.25; *P* < .001) and with a less steep MADRS reduction (β = 2.50; 95% CI, 0.43 to 4.57; *P* = .02). Because these analyses were exploratory and did not indicate treatment-specific moderation effects, they were interpreted as descriptive signals rather than subgroup-defining findings.

**Exploratory BDI-II latent growth and mixed-effects models**

Daily BDI-II scores showed substantial intraindividual variability in the acute change (Figure S3). The exploratory BDI-II latent growth models showed consistently poor global fit across multigroup and moderator specifications, with CFI values below 0.95 and RMSEA values well above 0.06, and even 0.08. Therefore, the BDI-II latent growth parameter estimates were not interpreted. The preregistered multigroup variance and covariance constraints referred to latent intercept and slope factors and were not directly transferred to the final mixed-effects framework.

Within the LMM analysis of the BDI-II data, exploratory treatment-effect moderation by personality disorder status and childhood trauma severity was significant only in the random-intercept version. The time × treatment group × personality disorder interaction (0.46 points/day; random intercept, 95% CI, 0.05 to 0.87; P = .03; random slope, 95% CI, −0.30 to 1.23; P = .23) suggested an attenuated active-vs-sham difference in daily BDI-II improvement among patients with comorbid personality disorder, and the time × treatment group × childhood trauma interaction (1.73 points/day; random intercept, 95% CI, 0.21 to 3.24; P = .03; random slope, 95% CI, −1.11 to 4.57; P = .23) an attenuated difference at higher CTQ levels, without an absence of predicted improvement in the active group at higher CTQ levels.

No significant exploratory BDI-II treatment-effect moderation was observed for relationship status (β = −0.001; 95% CI, −0.37 to 0.37; *P* > .99), social network size (β = 0.08; 95% CI, −0.59 to 0.74; *P* = .82), loneliness (β = 0.01; 95% CI, −0.01 to 0.03; *P* = .49), or employment status (β = −0.33; 95% CI, −0.69 to 0.03; *P* = .07). The employment-status interaction showed a considerable effect size, which did not however reach statistical significance, and was not interpreted as evidence of moderation. In the random-slope versions, none of these interactions was significant either (all P ≥ .32).

Several BDI-II moderator models also showed baseline or time-related associations irrespective of treatment-specific moderation (main effects in the sham group, the reference category). Childhood trauma severity was associated with higher baseline BDI-II severity (50.86; random intercept, 95% CI, 12.32 to 89.41; P = .01; random slope, 95% CI, 11.85 to 89.88; P = .01), and employment with lower baseline BDI-II severity (−10.82 points; random intercept, 95% CI, −19.72 to −1.93; P = .02; random slope, 95% CI, −19.65 to −1.99; P = .02); relationship status was not significantly associated with baseline severity (8.71 points; random intercept, 95% CI, −0.17 to 17.58; P = .054; random slope, P = .059). Employment was associated with a less steep BDI-II decrease in the random-intercept version (time × employment, 0.44 points/day; 95% CI, 0.18 to 0.71; P = .001) but not in the random-slope version (95% CI, −0.05 to 0.93; P = .08). These findings were considered exploratory and were not used to define treatment-selection subgroups.

Overall, the exploratory moderator analyses did not identify robust moderation of the primary clinician-rated MADRS treatment trajectory. The only treatment-specific moderation signals emerged for the self-reported BDI-II outcome, where the active-treatment advantage was attenuated among patients with comorbid personality disorder or higher childhood trauma severity. Because these analyses were exploratory, self-report-specific, not robust to random slopes, and not mirrored in the primary MADRS outcome, they should be interpreted as hypothesis-generating rather than as evidence for patient-selection criteria.

##### Post hoc sensitivity analyses

Results are summarized in Table S5. After adjustment for baseline severity, the acute treatment effect was smaller but remained significant (Model 1.3b: −2.92 points/week; 95% CI, −4.99 to −0.85; *P* = .006); ANCOVA and the mixed model for repeated measures gave consistent week-2 differences (−5.33 and −5.28 points). Of the unadjusted week-2 difference of −7.08 points, −1.23 points (95% CI, −3.40 to 0.92) were attributable to the latent baseline difference of 5.10 points, an upper bound for any bias from regression to the mean; the baseline-adjusted effect also remained significant with percentile bootstrap CIs (2000 resamples), including when the 943 resamples with inadmissible solutions were excluded. When all 57 randomized participants were included, the effect remained significant under the missing-at-random assumption (−3.68 points/week; 95% CI, −5.73 to −1.62; *P* < .001) and when non-initiators were assumed to follow the sham course (−3.18; 95% CI, −5.14 to −1.22; *P* = .002). In the tipping-point analysis, *P* exceeded .05 only when the imputed week-2 MADRS values of all non-initiators were shifted by 15 points against the active group. Adjustment for medication changes during stimulation reduced the effect only slightly (−3.11 points/week; 95% CI, −5.23 to −1.00; *P* = .004); the medication-change indicators themselves were not significantly associated with the slope.

During follow-up, the rate of change did not differ between groups. Observed week-8 means were the same in both groups (18.5), whereas baseline-adjusted differences favored active stimulation in all analyses (−2.1 to −4.7 points) without reaching significance (baseline-constrained piecewise model, −3.01 points; 95% CI, −6.82 to 0.80; *P* = .12; other analyses, *P* = .07 to .40). Six of the 7 participants without follow-up had responded at week 2. In the tipping-point analysis, all 121 combinations of shifts yielded estimates favoring active stimulation (−1.0 to −5.2 points), and none was significant. Adjustment of the piecewise models for sex and chronic MDD did not change these results (follow-up slope difference, 0.39 points/week; *P* = .26; baseline-adjusted week-8 difference, −3.58 points; *P* = .07).

Raters were distributed similarly across groups at weeks 0 to 2 (Fisher *P* ≥ .29), and 48 of 49 participants with rater information were rated by the same rater at weeks 0 and 2 (sham, 23 of 23;

active, 25 of 26). Adjustment for rater did not reduce the treatment effect (Model 1.3: −3.62 points/week; 95% CI, −5.63 to −1.62; $P < .001$; Model 1.3b: −3.14; 95% CI, −5.23 to −1.06; $P = .003$). Results were similar in the mixed model (−3.51; 95% CI, −5.69 to −1.32) and in participants with the same rater at weeks 0 and 2 (n = 48; −3.38; 95% CI, −5.41 to −1.36), and the follow-up results were unchanged (follow-up slope difference, 0.44 points/week; 95% CI, −0.23 to 1.12; baseline-adjusted week-8 difference, −3.00 points; 95% CI, −6.76 to 0.77).

BDI-II. FDR-adjusted P values of the confirmatory BDI-II tests are given in Table 2 of the main text for the random-slope version (overall decrease, P = .003; time × group, P = .34) and in the Results of the main text for the random-intercept version (overall decrease, P < .001; time × group, P = .02; episode duration, all P ≥ .12). For the time × group effect, results similar to the random-slope version were obtained with cluster-robust standard errors (P = .17) and in the ANCOVA of day-10 scores (−2.05 points; 95% CI, −5.07 to 0.98; P = .18); with CAR(1) residuals, the confidence interval was narrower but also included zero (−0.23 points/day; 95% CI, −0.48 to 0.02; P = .08). Kenward–Roger degrees of freedom and inclusion of the participant with a baseline BDI-II score below 14 did not change the random-intercept result (P = .01 for both; all post hoc analyses shown in Table S5).

#### Side effects

Side-effect profiles were comparable between treatment groups. Headache was reported by 10 of 24 participants in the sham group and 15 of 27 participants in the active group, corresponding to occurrence rates of 41.7% and 55.6%, respectively; this difference was not statistically significant ($\chi^2(1) = 0.98$, $P = .32$; Table S4). Discomfort occurred in 6 of 24 sham participants and 7 of 27 active participants, corresponding to rates of 25.0% and 25.9%, respectively ($\chi^2(1) = 0.01$, $P = .94$). Nausea was reported by 5 of 24 sham participants and 4 of 27 active participants, corresponding to rates of 20.8% and 14.8%, respectively (OR = 0.67, $P = .72$). Fatigue was reported on at least one treatment day by 10 of 24 sham participants and 8 of 27 active participants (41.7% and 29.6%; $\chi^2(1) = 0.81$, P = .37). No serious adverse events, seizures, syncope, (hypo)manic symptoms, treatment-emergent suicidal ideation, or cognitive complaints occurred in either group. The number of symptom-positive treatment days for headache, discomfort, and nausea, as well as fatigue severity ratings during the treatment phase, also did not differ significantly between groups (all $P \geq .18$; Table S4).

#### Blinding Integrity

Across complementary analyses, treatment guesses did not differ from chance and were not related to guess confidence or tolerability.

**Blinding accuracy.** Treatment-guess data were available for 38 participants. Overall, 24 of 38 participants (63.2%) correctly guessed their treatment assignment (sham, 12 of 18; active, 12 of 20). Guessed treatment assignment was not significantly associated with actual treatment assignment (Fisher exact test: OR = 2.91; 95% CI, 0.67 to 13.98; $P = .12$). Because participants may partly infer assignment from perceived benefit or treatment experience, guess accuracy alone should not be interpreted as direct evidence of unblinding (8). Bang blinding indices were 0.20 (95% CI, −0.23 to 0.63) in the active group and 0.33 (95% CI, −0.10 to 0.77) in the sham group; both confidence intervals included 0, so guessing beyond chance level could not be demonstrated in either group. By a commonly used rule of thumb, indices between −0.2 and 0.2 indicate random guessing (16); the point estimate lay at this threshold in the active group and above it in the sham group. Correct guesses in the sham group may partly reflect the absence of perceived improvement, particularly because guesses were obtained at follow-up.

**Confidence and guess accuracy.** Confidence ratings did not differ meaningfully between participants who correctly guessed their treatment assignment and those who guessed incorrectly (median [IQR], 83.0 [47.8] vs 59.5 [30.8]; Wilcoxon rank sum test: $W = 141$; estimated location shift (incorrect minus correct), −5.0; 95% CI, −28.0 to 12.0; $P = .42$; Figure S4). The effect size was small ($r = 0.13$).

**Tolerability of stimulation.** Mean tolerability ratings of all 51 participants (sham, mean [SD], 6.43 [1.56]; active, 6.65 [1.33]) did not differ significantly between the sham and active groups (Wilcoxon rank sum test: $W = 296$; estimated location shift (sham minus active), −0.23; 95% CI, −1.13 to 0.63; $P = .60$; Figure S5). The effect size was small ($r = 0.08$).

**Tolerability and treatment guess.** Mean tolerability ratings were not significantly associated with guessed treatment assignment in logistic regression (participants with a treatment guess, n = 38; OR = 0.78; 95% CI, 0.48 to 1.28; *P* = .32), indicating that stimulation tolerability did not meaningfully predict whether participants guessed Active vs Sham assignment.

#### BDI-II Screening and pre-treatment baseline scores

BDI-II scores decreased before the start of stimulation. Among participants with both screening and pre-treatment baseline assessments available (n = 50), mean BDI-II scores decreased from 32.98 (SD, 8.20) at screening to 28.00 (SD, 10.87) at pre-treatment baseline. The mean paired difference was −4.98 points (95% CI, −7.07 to −2.89; $t(49) = -4.79$; $P < .001$; Cohen's dz = −0.68), calculated as pre-treatment baseline minus screening. This early decrease is consistent with the inpatient study context, in which participants had already spent time in a structured clinical environment and received routine clinical care before the start of stimulation. Therefore, the treatment baseline assessment immediately before the first stimulation session was used as Time 0 for all longitudinal treatment-phase models.

#### Post hoc quality-control analysis of neuronavigation targets

Deviations between the originally selected and the corrected coordinates were in the millimeter to low-centimeter range. In the active-treatment group, the mean distance between the originally selected and corrected surface coordinate was 8.61 mm. In the sham group, the mean distance was 10.37 mm.

Target-coordinate deviation was not significantly associated with acute MADRS reduction in either group (active: $\rho = 0.03$, $P = .88$; sham: $\rho = 0.12$, $P = .62$). These findings did not provide evidence that the observed target-coordinate deviations explained variability in acute MADRS improvement.

These quality-control analyses should not be interpreted as a direct test of the broader personalized-target proximity hypothesis. Prior retrospective work relating proximity to personalized targets and clinical response involved substantially larger distances between clinically applied and personalized targets, with a median distance of 30 mm and values exceeding 60 mm (17). In contrast, the present analysis examined small-to-moderate deviations (0.1–1.5 cm) between two implementations of the same seedmap-based, cluster-guided targeting procedure. The difference is important because the present target was defined as the center of gravity of an anticorrelated DLPFC cluster, not as a single maximally anticorrelated voxel. This cluster-level definition may provide more tolerance to small-to-moderate coordinate deviations, particularly when the original and corrected coordinates remain within or near the same functionally relevant anticorrelated DLPFC region (17, 2).

Several technical considerations further support this cautious interpretation. TMS with a figure-8 coil does not stimulate a mathematical point, but induces a spatially extended electric field; reviews and modeling work describe figure-8 stimulation as relatively focal but spatially extended, with estimates of focal cortical influence in the range of approximately 1.5–2 $cm^2$ and spatial resolution around 1.5–2 cm under standard stimulation conditions (18–21). Given that the maximum coordinate deviation was below 15 mm, the corrected coordinates likely remained within or near the broader spatial extent of the induced field, although this inference is approximate and cannot replace individualized electric-field modeling. Focality and depth also depend on coil geometry and stimulation parameters (22, 18). Target engagement additionally depends on coil orientation, local anatomy, scalp-to-cortex distance, stimulation intensity, and individual electric-field distribution; coil orientation in particular can affect the magnitude and site of cortical activation (23). Therefore, the present analysis does not establish a general tolerance threshold for targeting error, but indicates that the observed deviations were not associated with reduced acute MADRS improvement in this dataset.

### Supplementary Tables

##### Table S1. Comorbid psychiatric diagnoses

| | Total (*n* = 51) | Sham (*n* = 24) | Active (*n* = 27) | |
|---|---|---|---|---|
| **Comorbid psychiatric diagnosis** | **No. (%)** | **No. (%)** | **No. (%)** | ***P*** |
| No. participants with comorbidities | 42 (82%) | 19 (79%) | 23 (85%) | .72 |
| Anxiety | 26 (51%) | 12 (50%) | 14 (52%) | >.99 |
| Personality disorder (PD) | 17 (33%) | 10 (42%) | 7 (26%) | .25 |
| Emotionally unstable PD of type Borderline | 5 (10%) | 2 (8%) | 3 (11%) | >.99 |
| Combined PD | 3 (6%) | 1 (4%) | 2 (7%) | >.99 |
| Obsessive-compulsive PD | 1 (2%) | 1 (4%) | 0 (0%) | .47 |
| Schizotypal PD | 2 (4%) | 2 (8%) | 0 (0%) | .22 |
| Narcissistic PD | 2 (4%) | 1 (4%) | 1 (4%) | >.99 |
| Avoidant PD | 4 (8%) | 3 (13%) | 1 (4%) | .33 |
| Posttraumatic stress disorder | 8 (16%) | 3 (13%) | 5 (19%) | .71 |
| Obsessive-compulsive disorder | 5 (10%) | 2 (8%) | 3 (11%) | >.99 |
| Attention-Deficit/Hyperactivity Disorder (ADHD)/Attention-Deficit Disorder (ADD) | 4 (8%) | 2 (8%) | 2 (7%) | >.99 |
| Eating disorder | 3 (6%) | 1 (4%) | 2 (7%) | >.99 |
| Somatoform disorder | 2 (4%) | 0 (0%) | 2 (7%) | .49 |
| Dissociative disorder | 2 (4%) | 0 (0%) | 2 (7%) | .49 |
| Autism | 1 (2%) | 1 (4%) | 0 (0%) | .47 |

Note: Comorbid diagnoses are shown separately for the sham and active groups. P values were calculated using two-sided Fisher exact tests.

**Table S2. Concomitant psychotropic medication by treatment group**

| Medication | Sham, n/N (%) | Active, n/N (%) | P |
|---|---|---|---|
| *Antidepressant classes* | | | |
| SNRI | 17/24 (70.8) | 14/27 (51.9) | .25 |
| NaSSA | 7/24 (29.2) | 12/27 (44.4) | .39 |
| TCA | 7/24 (29.2) | 9/27 (33.3) | .77 |
| Atypical antidepressant | 7/24 (29.2) | 8/27 (29.6) | >.99 |
| SSRI | 2/24 (8.3) | 6/27 (22.2) | .26 |
| NDRI | 1/24 (4.2) | 0/27 (0.0) | .47 |
| NRI | 0/24 (0.0) | 1/27 (3.7) | >.99 |
| *Augmentation and adjunctive medication classes* | | | |
| Atypical antipsychotic | 11/24 (45.8) | 10/27 (37.0) | .58 |
| Gabapentinoid | 1/24 (4.2) | 6/27 (22.2) | .10 |
| Typical antipsychotic | 2/24 (8.3) | 4/27 (14.8) | .67 |
| Thyroid hormone | 3/24 (12.5) | 2/27 (7.4) | .66 |
| Benzodiazepine | 2/24 (8.3) | 1/27 (3.7) | .60 |
| Mood stabilizer | 1/24 (4.2) | 2/27 (7.4) | >.99 |
| Z-drug | 1/24 (4.2) | 0/27 (0.0) | .47 |
| *Medication changes* | | | |
| Any change from admission through the stimulation phase[b] | 19/24 (79.2) | 24/27 (88.9) | .45 |
| Initiation, discontinuation, addition, or switch during stimulation[c] | 9/24 (37.5) | 11/27 (40.7) | >.99 |
| Dose adjustment during stimulation[d] | 15/24 (62.5) | 21/27 (77.8) | .36 |

Abbreviations: NaSSA, noradrenergic and specific serotonergic antidepressant; NDRI, norepinephrine-dopamine reuptake inhibitor; NRI, norepinephrine reuptake inhibitor; SNRI, serotonin-norepinephrine reuptake inhibitor; SSRI, selective serotonin reuptake inhibitor; TCA, tricyclic antidepressant.

Note: Participants are counted once per class, irrespective of the number of agents received within the class; P values are from two-sided Fisher exact tests. Agents: SNRI, desvenlafaxine, duloxetine, milnacipran, venlafaxine; NaSSA, mirtazapine; TCA, amitriptyline, clomipramine, doxepin, opipramol, tianeptine, trimipramine; atypical antidepressant, agomelatine; SSRI, citalopram, escitalopram, fluoxetine, sertraline; NDRI, bupropion[a]; NRI, atomoxetine; atypical antipsychotics, amisulpride, aripiprazole, olanzapine, quetiapine, risperidone; typical antipsychotics, pipamperone, promethazine; gabapentinoid, pregabalin; thyroid hormone, L-thyroxine; benzodiazepine, lorazepam[a]; mood stabilizer, lithium; Z-drug, zopiclone.

[a] Discontinued before the first stimulation session. [b] Initiation, discontinuation, addition, switch, or dose adjustment of a psychotropic medication from hospital admission through the stimulation phase. [c] From the first to the last stimulation session. [d] Any increase or decrease in the dose of a psychotropic medication during the stimulation phase.

#### Table S3. Response and remission at post-treatment

| End point | Sham, n/N (%) | Active, n/N (%) | Risk difference, % points (95% CI) | OR (95% CI)[a] | P[a] | Adjusted OR (95% CI)[b] | P[b] |
|---|---|---|---|---|---|---|---|
| MADRS response | 3/23 (13.0) | 11/26 (42.3) | 29.3 (3.9 to 49.8) | 4.73 (1.01 to 31.09) | .03 | 8.53 (1.88 to 57.40) | .004 |
| MADRS remission | 3/24 (12.5) | 7/26 (26.9) | 14.4 (−8.3 to 35.2) | 2.53 (0.49 to 17.36) | .29 | 5.73 (1.11 to 44.73) | .04 |
| BDI-II response | 1/22 (4.5) | 4/27 (14.8) | 10.3 (−9.1 to 28.3) | 3.57 (0.32 to 188.07) | .36 | — | — |
| BDI-II remission | 5/22 (22.7) | 7/27 (25.9) | 3.2 (−21.1 to 25.8) | 1.19 (0.27 to 5.68) | .80 | — | — |

Abbreviations: BDI-II, Beck Depression Inventory-II; CI, confidence interval; MADRS, Montgomery-Åsberg Depression Rating Scale; OR, odds ratio.

Note: Denominators comprise participants with the ratings required for each end point (response: treatment baseline and post-treatment; remission: post-treatment). Risk differences are active minus sham, with Newcombe hybrid score 95% CIs.

[a] Preregistered unadjusted analysis: Fisher exact test (conditional maximum likelihood OR with exact 95% CI) for MADRS response and remission and BDI-II response; $\chi^2$ test for BDI-II remission ($\chi^2(1) = 0.07$). [b] Firth penalized-likelihood logistic regression adjusted for treatment-baseline MADRS (linear term; n = 49 for both outcomes), with profile penalized-likelihood 95% CIs and P values (penalized likelihood-ratio $\chi^2(1) = 8.22$ for response and 4.41 for remission). The corresponding unadjusted Firth ORs were 4.35 (95% CI, 1.19 to 19.53) and 2.36 (95% CI, 0.61 to 10.90). BDI-II outcomes were not adjusted because baseline BDI-II scores did not differ between groups.

#### Table S4. Adverse events and side effects during the stimulation phase

| | Sham (n = 24) | Active (n = 27) | Effect (95% CI) | Test statistic | P |
|---|---|---|---|---|---|
| *Occurrence on at least 1 treatment day, n/N (%); effect, odds ratio* | | | | | |
| Headache | 10/24 (41.7) | 15/27 (55.6) | 1.73 (0.50 to 6.15) | $\chi^2(1) = 0.98$ | .32 |
| Discomfort | 6/24 (25.0) | 7/27 (25.9) | 1.05 (0.25 to 4.56) | $\chi^2(1) = 0.01$ | .94 |
| Nausea | 5/24 (20.8) | 4/27 (14.8) | 0.67 (0.11 to 3.59) | Fisher exact | .72 |
| Fatigue | 10/24 (41.7) | 8/27 (29.6) | 0.60 (0.16 to 2.18) | $\chi^2(1) = 0.81$ | .37 |
| Serious adverse events | 0/24 (0.0) | 0/27 (0.0) | — | — | — |
| Seizures | 0/24 (0.0) | 0/27 (0.0) | — | — | — |
| Syncope | 0/24 (0.0) | 0/27 (0.0) | — | — | — |
| Mania or hypomania | 0/24 (0.0) | 0/27 (0.0) | — | — | — |
| Treatment-emergent suicidal ideation | 0/24 (0.0) | 0/27 (0.0) | — | — | — |
| Cognitive complaints | 0/24 (0.0) | 0/27 (0.0) | — | — | — |
| *Symptom-positive days and fatigue rating, mean (SD); effect, location shift* | | | | | |
| Headache-positive days | 1.29 (2.27) | 1.93 (2.32) | 0.00 (0.00 to 1.00) | W = 269.5 | .27 |
| Discomfort-positive days | 1.25 (2.91) | 1.04 (2.39) | 0.00 (0.00 to 0.00) | W = 325.0 | .99 |
| Nausea-positive days | 0.67 (1.69) | 0.52 (1.65) | 0.00 (0.00 to 0.00) | W = 343.0 | .59 |
| Fatigue severity rating | 4.67 (1.47) | 5.15 (1.36)[a] | 0.56 (−0.22 to 1.29) | W = 242.0 | .18 |

Abbreviations: CI, confidence interval; OR, odds ratio; SD, standard deviation; W, Wilcoxon rank-sum statistic.

Note: Side effects were assessed daily before stimulation with a structured questionnaire that included a free-text option; because day-1 ratings preceded the first session, analyses cover treatment days 2 to 10. Side effects were not assessed during follow-up. Suicidal ideation was monitored with MADRS item 10 and by clinical assessment, (hypo)manic symptoms by clinical assessment and the free-text option, and cognitive complaints with the questionnaire and its free-text option. Treatment-emergent suicidal ideation was defined as an increase of at least 2 points from baseline on MADRS item 10 or as emergent suicidal ideation in the clinical assessment. Increases of 1 point on MADRS item 10 occurred in 4 of 26 active and 3 of 23 sham participants with evaluable ratings (only transient in 2 per group); no participant had an increase of 2 or more points. Occurrence was defined as reporting the respective symptom at least once during treatment and was compared using Pearson $\chi^2$ tests, or Fisher exact tests when a cell count was below 5; odds ratios compare the odds of occurrence in the active group with those in the sham group (values greater than 1 indicate higher odds in the active group). Symptom-positive days are the number of treatment days on which participants reported the respective symptom; days and the fatigue rating were compared using Wilcoxon rank-sum tests, with the Hodges-Lehmann estimate of the location shift (active minus sham; positive values indicate more symptom-positive days or higher ratings in the active group). Events were recorded on each treatment day irrespective of attributed cause. Fatigue was assessed in two ways: as a side effect reported on at least one treatment day (occurrence) and as a rating of overall fatigue severity during the treatment phase (scale 0–10; higher values indicate more fatigue).

[a] n = 26.

**Table S5. Post hoc sensitivity analyses**

| Analysis | *Estimate (95% CI)* | *P value* | *N* |
|---|---|---|---|
| **Acute treatment effect: group difference in MADRS slope (points/week)** | | | |
| Model 1.3 (preregistered) | −3.54 (−5.53 to −1.55) | <.001 | 51 |
| Model 1.3b, adjusted for latent baseline | −2.92 (−4.99 to −0.85) | .006 | 51 |
| All randomized, multiple imputation (MAR) | −3.68 (−5.73 to −1.62) | <.001 | 57 |
| All randomized, reference-based imputation | −3.18 (−5.14 to −1.22) | .002 | 57 |
| All randomized, reference-based imputation, Model 1.3b | −2.54 (−4.58 to −0.49) | .02 | 57 |
| All randomized, tipping point δ = 12.5 | −2.39 (−4.64 to −0.15) | .04 | 57 |
| All randomized, tipping point δ = 15 | −2.15 (−4.48 to 0.18) | .07 | 57 |
| Adjusted for medication changes (start/stop/switch and dose) | −3.11 (−5.23 to −1.00) | .004 | 51 |
| Adjusted for start/stop/switch only | −3.45 (−5.43 to −1.47) | .001 | 51 |
| Adjusted for rater (Model 1.3) | −3.62 (−5.63 to −1.62) | <.001 | 51 |
| Adjusted for rater (Model 1.3b) | −3.14 (−5.23 to −1.06) | .003 | 51 |
| Linear mixed model, adjusted for rater | −3.51 (−5.69 to −1.32) | .002 | 51 |
| Same rater at weeks 0 and 2 (Model 1.3) | −3.38 (−5.41 to −1.36) | .001 | 48 |
| **Acute treatment effect: group difference at week 2 (MADRS points)** | | | |
| Model 1.3b (2 × slope difference) | −5.84 (−9.99 to −1.69) | .006 | 51 |
| Part attributable to the latent baseline difference (Model 1.3 minus Model 1.3b) | −1.23 (−3.40 to 0.92) | .26 | 51 |
| Model 1.3, adjusted for rater (2 × slope difference) | −7.25 (−11.25 to −3.24) | <.001 | 51 |
| ANCOVA, complete cases | −5.33 (−9.82 to −0.83) | .02 | 49 |
| MMRM | −5.28 (−9.40 to −1.16) | .01 | 51 |
| **Follow-up: group difference in MADRS slope from week 2 to week 8 (points/week)** | | | |
| Piecewise LMM (Table S7 in Supplement 3) | 0.39 (−0.29 to 1.07) | .26 | 51 |
| Piecewise LMM, adjusted for sex and chronic MDD | 0.39 (−0.29 to 1.06) | .26 | 51 |
| Piecewise LMM, adjusted for rater | 0.44 (−0.23 to 1.12) | .19 | 51 |
| **Follow-up: group difference at week 8 (MADRS points)** | | | |
| Piecewise LMM, not adjusted for baseline | 0.39 (−4.59 to 5.37) | .88 | 51 |
| Piecewise LMM, baseline-constrained | −3.01 (−6.82 to 0.80) | .12 | 51 |
| Piecewise LMM, baseline-constrained, adjusted for sex and chronic MDD | −3.58 (−7.39 to 0.23) | .07 | 51 |
| Piecewise LMM, baseline-constrained, adjusted for rater | −3.00 (−6.76 to 0.77) | .12 | 51 |
| MMRM | −3.66 (−8.63 to 1.32) | .15 | 51 |
| ANCOVA, complete cases | −3.03 (−8.33 to 2.26) | .25 | 43 |

| Analysis | Estimate (95% CI) | P value | N |
|---|---|---|---|
| ANCOVA after multiple imputation (MAR) | −3.13 (−8.28 to 2.02) | .23 | 51 |
| ANCOVA, LOCF | −4.68 (−9.86 to 0.50) | .08 | 50 |
| ANCOVA, BOCF | −2.11 (−7.07 to 2.85) | .40 | 50 |
| Tipping point, active +10 and sham −10 points | −0.99 (−6.26 to 4.27) | .71 | 51 |
| Tipping point, active −10 and sham +10 points | −5.16 (−10.78 to 0.46) | .07 | 51 |
| **Secondary outcome: group difference in BDI-II slope (points/day)** | | | |
| Random intercept (Satterthwaite df) | −0.23 (−0.41 to −0.05) | .01 | 50 |
| Random intercept (Kenward–Roger df) | −0.23 (−0.41 to −0.05) | .01 | 50 |
| Random intercept and random slope for time | −0.23 (−0.57 to 0.10) | .17 | 50 |
| Random intercept, CR2 cluster-robust standard errors | −0.23 (−0.57 to 0.11) | .17 | 50 |
| Random intercept, CAR(1) residuals | −0.23 (−0.48 to 0.02) | .08 | 50 |
| Random intercept, including participant with baseline BDI-II < 14 | −0.23 (−0.40 to −0.05) | .01 | 51 |
| ANCOVA of day-10 scores (BDI-II points) | −2.05 (−5.07 to 0.98) | .18 | 50 |
| **Follow-up: group difference in BDI-II slope from day 10 to week 8 (points/day)** | | | |
| Piecewise LMM, random intercept (Table S8 in Supplement 3) | 0.01 (−0.05 to 0.07) | .72 | 50 |
| Piecewise LMM, random intercept and random slope for time | 0.01 (−0.04 to 0.07) | .67 | 50 |

Abbreviations: ANCOVA, analysis of covariance; BDI-II, Beck Depression Inventory-II; BOCF, baseline observation carried forward; CAR(1), continuous-time first-order autoregressive residual structure; CI, confidence interval; CR2, cluster-robust bias-reduced standard errors; LMM, linear mixed model; LOCF, last observation carried forward; MADRS, Montgomery-Åsberg Depression Rating Scale; MAR, missing at random; MDD, major depressive disorder; MMRM, mixed model for repeated measures.

Note: Negative values indicate lower (better) MADRS or BDI-II scores with active stimulation. Multiple-imputation estimates are pooled over 50 imputations. Tipping point (all randomized): shift δ of the imputed week-2 MADRS values of non-initiators against the active group (+δ active, −δ sham). Tipping point (follow-up): shift of the imputed week-8 values of participants without follow-up (positive = worse than predicted); none of the 121 combinations from −10 to +10 points per arm was significant. Piecewise models are fitted without covariates unless stated. Rater: MADRS rater at each visit as fixed-effect covariate. BDI-II analyses include 50 participants after exclusion of 1 participant with a baseline BDI-II score below 14 unless stated; the ANCOVA compares day-10 scores adjusted for day 0. P values are not adjusted for multiplicity.

### Supplementary Figures

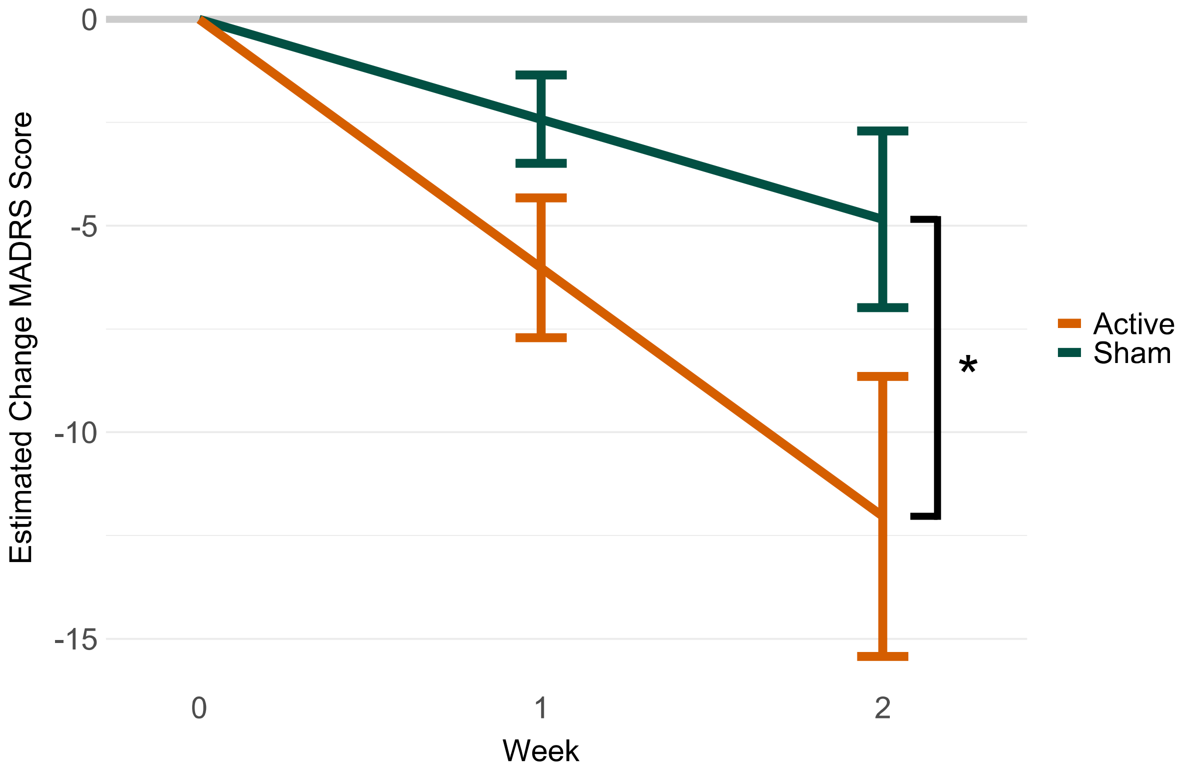

**Figure S1. Model-estimated MADRS change during acute treatment.**

Model-estimated change in Montgomery-Åsberg Depression Rating Scale (MADRS) scores is shown from baseline to week 2 by treatment group. Error bars indicate 95% CI. The asterisk denotes the significant group-by-time effect.

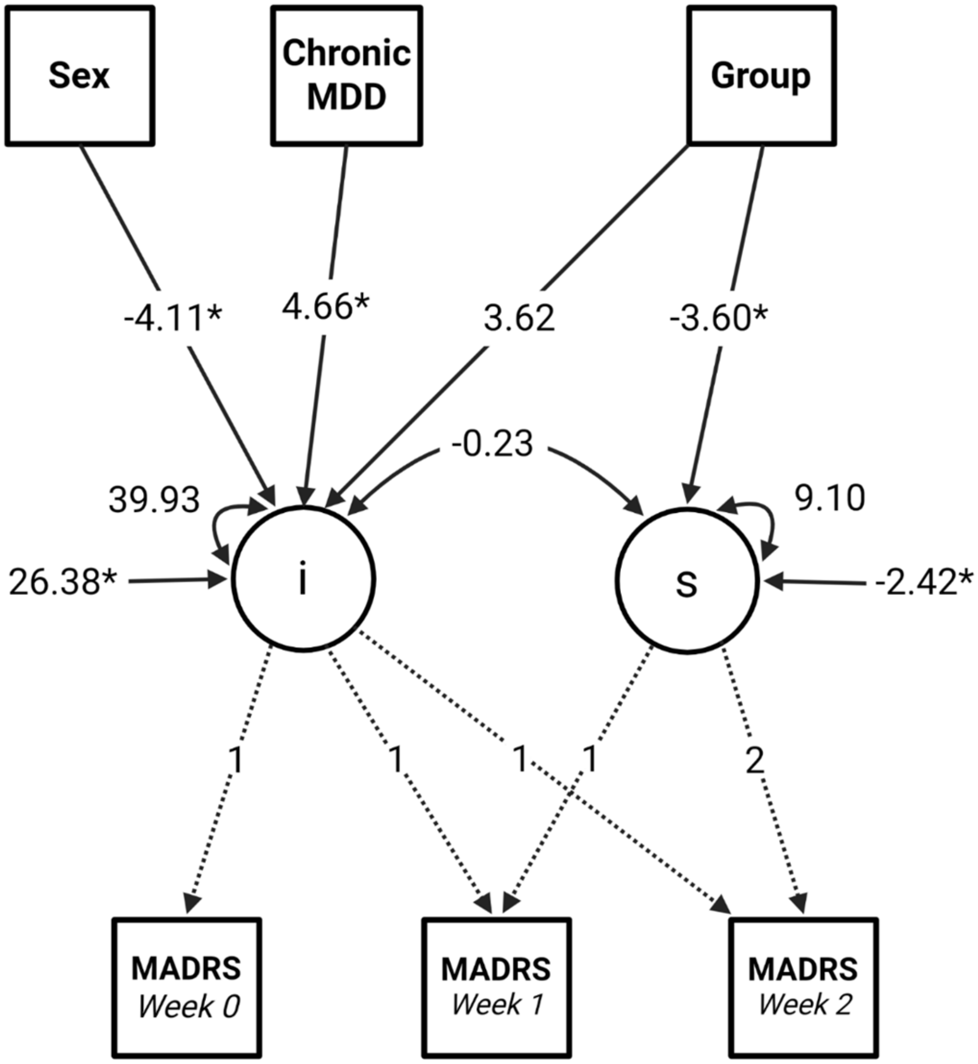

**Figure S2. Final latent growth curve model for MADRS scores.**

Structural equation model of Montgomery-Åsberg Depression Rating Scale (MADRS) scores from baseline to week 2. The intercept factor (i) represents baseline MADRS severity, and the slope factor (s) represents linear weekly change. The time variable for the slope factor was coded in weeks (0, 1, 2); therefore, slope estimates represent the expected change in MADRS score per week. Values are unstandardized estimates, except for the intercept–slope association, which is shown as a correlation; asterisks indicate $P < .05$ for regression paths and intercepts.

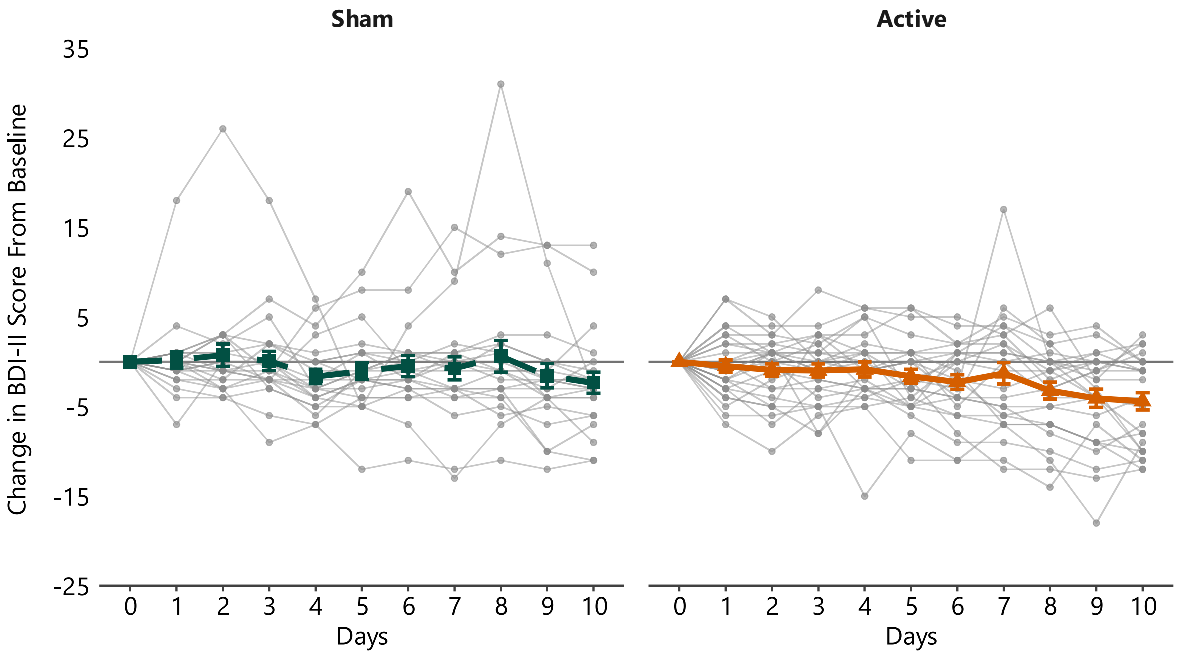

**Figure S3. Individual BDI-II change during acute treatment.**

Individual change in Beck Depression Inventory-II (BDI-II) scores from treatment baseline is shown across the 10-day treatment phase by group. Gray lines represent individual participants; colored lines show observed group mean change, with error bars indicating SE. Negative values indicate symptom improvement.

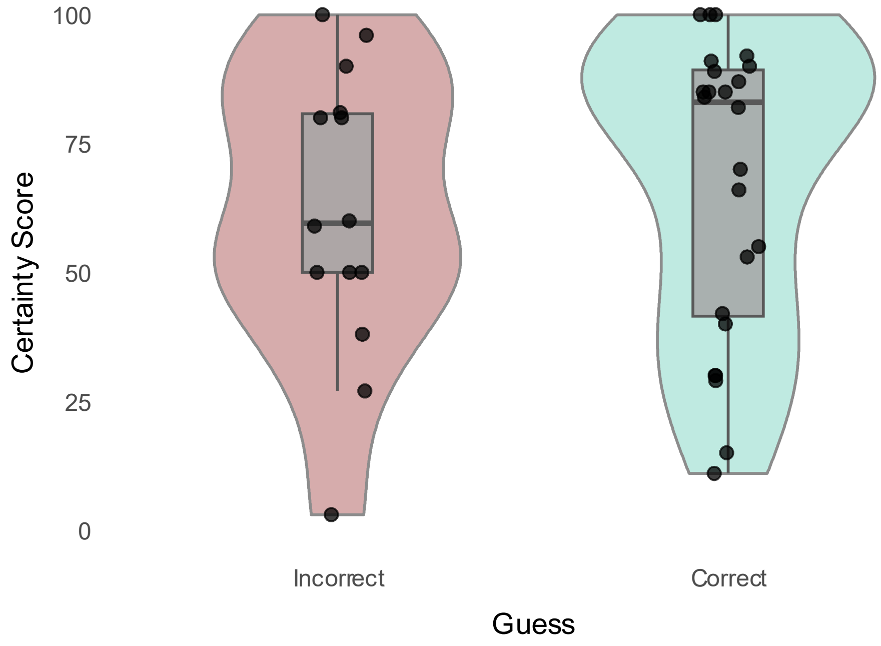

**Figure S4. Treatment-guess confidence by guess accuracy.**

Individual confidence ratings are shown separately for participants with incorrect and correct treatment guesses. Higher scores indicate higher confidence in the treatment guess.

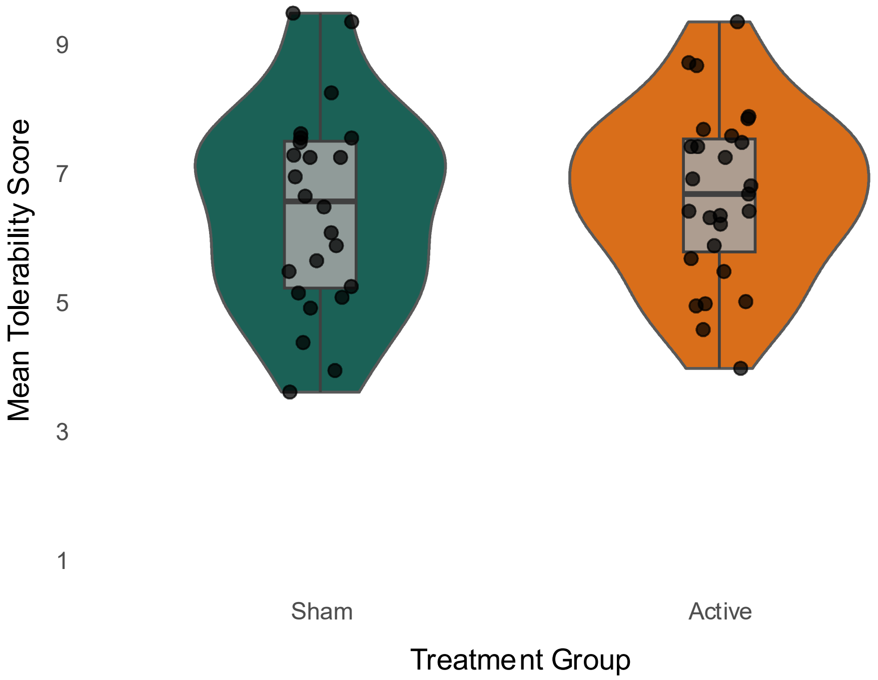

**Figure S5. Perceived stimulation tolerability by treatment group.**

Individual participant mean tolerability ratings across the 30 stimulation sessions are shown separately for the sham and active treatment groups. Higher scores indicate greater perceived tolerability.

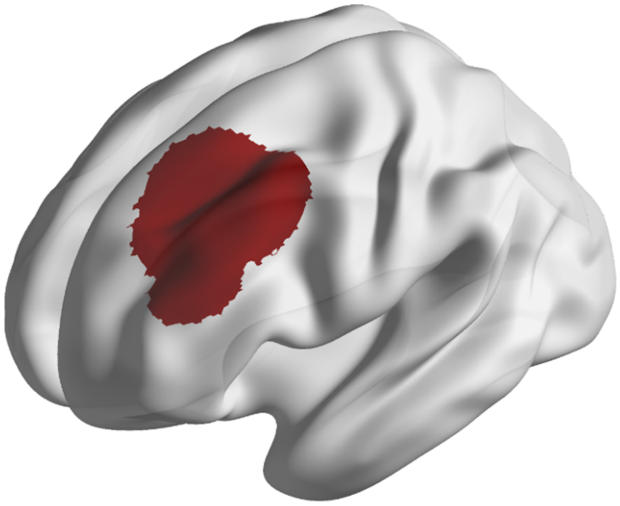

**Figure S6. Restricted DLPFC Mask for individualized, connectivity-guided target selection.**

The dorsolateral prefrontal cortex mask was restricted to exclude regions close to the temples and infero-frontal regions close to the eyes, where stimulation is less tolerable because of peripheral muscle and nerve activation.

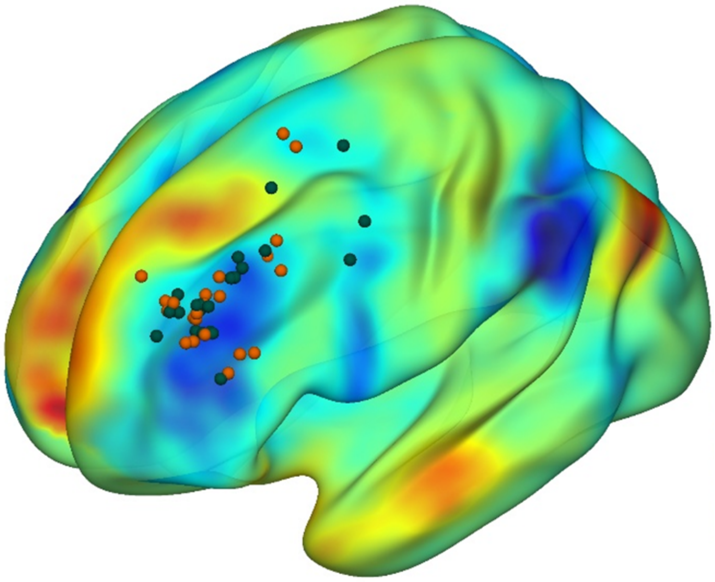

**Figure S7. Spatial Distribution of individualized functional connectivity MRI–guided target locations**

Individualized stimulation targets are shown for the active stimulation group (orange spheres; *n* = 27) and the sham control group (green spheres; *n* = 24). Targets were localized within the left dorsolateral prefrontal cortex (dlPFC) using individual resting-state functional MRI. Specifically, targets were defined by the maximal negative functional connectivity (anticorrelation) with the subgenual anterior cingulate cortex. To optimize spatial reliability and signal-to-noise ratio, target identification was based on anticorrelated clusters rather than single peak voxels, adhering to the methodology described by Cash et al. (2021)(2).
