## Supplement 3 for "Adjunctive Accelerated Theta-Burst Stimulation for Inpatient Treatment-Resistant Depression: A Randomized Clinical Trial"

This supplement reports the fit indices and parameter estimates of the preregistered confirmatory and exploratory longitudinal models of the acute treatment phase for the clinician-rated Montgomery-Åsberg Depression Rating Scale (MADRS) and the self-reported Beck Depression Inventory-II (BDI-II), and the parameter estimates of the exploratory piecewise follow-up models. Model specifications are described in the Supplementary Methods and the results are summarized in the Supplementary Results of Supplement 2; the confirmatory estimates are shown in Table 2 of the main text.

MADRS trajectories were analyzed with latent growth models (robust maximum likelihood). The preregistered BDI-II latent growth models showed inadequate fit and were not interpreted; BDI-II trajectories were therefore analyzed with linear mixed models, each fitted with a random intercept and with a random intercept and a correlated random slope for time (both deviations from the preregistration). For the linear mixed models, fit indices and likelihood-ratio tests are based on maximum likelihood and parameter estimates on restricted maximum likelihood.

#### Contents

Table S6. Fit indices of all longitudinal models
Table S7. Parameter estimates of the MADRS models
Table S8. Parameter estimates of the linear mixed models of daily BDI-II scores

### Table S6. Fit indices of all longitudinal models

#### A. Latent growth models (MADRS and BDI-II; robust maximum likelihood)

| Model | AIC | BIC | sBIC | CFI | RMSEA (95% CI) |
|---|---|---|---|---|---|
| **MADRS, confirmatory models** | | | | | |
| Model 1.1: Linear latent growth model | 985.9 | 1001.3 | 976.2 | 1.000 | 0.000 (0.000 to 0.329) |
| Model 1.2: Non-linear latent growth model | 987.4 | 1004.8 | 976.5 | 1.000 | 0.000 (0.000 to 0.000) |
| Model 1.3: Group comparison model | 977.9 | 997.2 | 965.8 | 1.000 | 0.000 (0.000 to 0.275) |
| Model 1.3b: Group model adjusting for baseline differences (sensitivity, not preregistered) | 977.9 | 997.2 | 965.8 | 1.000 | 0.000 (0.000 to 0.275) |
| Model 1.4a: Covariate of age | 980.5 | 1003.7 | 966.0 | 1.000 | 0.000 (0.000 to 0.213) |
| Model 1.4b: Covariate of sex | 975.4 | 998.6 | 960.9 | 0.994 | 0.063 (0.000 to 0.252) |
| Model 1.4c: Covariate of chronic MDD | 977.1 | 1000.3 | 962.6 | 1.000 | 0.000 (0.000 to 0.201) |
| Model 1.5: Moderation by episode duration | 980.8 | 1007.8 | 963.8 | 1.000 | 0.000 (0.000 to 0.148) |
| Model 1.6: Final combined model | 971.8 | 995.0 | 957.3 | 1.000 | 0.000 (0.000 to 0.133) |
| **MADRS, exploratory models** | | | | | |
| Model 1.7a: Unconstrained covariance | 979.3 | 1010.2 | 960.0 | 1.000 | 0.000 (0.000 to 0.379) |
| Model 1.7b: Constrained covariance | 977.8 | 1006.7 | 959.6 | 1.000 | 0.000 (0.000 to 0.300) |
| Model 1.8a: Unconstrained intercept variance | 979.3 | 1010.2 | 960.0 | 1.000 | 0.000 (0.000 to 0.379) |
| Model 1.8b: Constrained intercept variance | 977.9 | 1006.9 | 959.8 | 1.000 | 0.000 (0.000 to 0.309) |
| Model 1.9a: Unconstrained slope variance | 979.3 | 1010.2 | 960.0 | 1.000 | 0.000 (0.000 to 0.379) |
| Model 1.9b: Constrained slope variance | 977.6 | 1006.6 | 959.5 | 1.000 | 0.000 (0.000 to 0.293) |
| Model 1.10: Moderation by personality disorder | 976.8 | 1003.8 | 959.9 | 1.000 | 0.000 (0.000 to 0.200) |
| Model 1.11: Moderation by childhood trauma | 978.4 | 1005.5 | 961.5 | 0.988 | 0.077 (0.000 to 0.236) |
| Model 1.12: Moderation by relationship status | 976.8 | 1003.8 | 959.8 | 1.000 | 0.000 (0.000 to 0.178) |
| Model 1.13: Moderation by social network size | 960.8 | 987.6 | 943.6 | 0.994 | 0.053 (0.000 to 0.226) |
| Model 1.14: Moderation by loneliness | 956.6 | 983.4 | 939.4 | 1.000 | 0.000 (0.000 to 0.158) |
| Model 1.15: Moderation by employment status | 974.3 | 1001.4 | 957.4 | 1.000 | 0.000 (0.000 to 0.190) |
| **BDI-II, confirmatory models (inadequate fit; not interpreted)** | | | | | |
| Model 2.1: Linear latent growth model | 3425.3 | 3457.8 | 3404.4 | 0.850 | 0.236 (0.206 to 0.265) |
| Model 2.2: Non-linear latent growth model | 3478.7 | 3530.3 | 3445.6 | 0.818 | 0.279 (0.249 to 0.311) |
| Model 2.3: Group comparison model | 3427.4 | 3463.8 | 3404.1 | 0.843 | 0.226 (0.199 to 0.255) |
| Model 2.4a: Covariate of age | 3430.6 | 3470.8 | 3404.9 | 0.845 | 0.213 (0.187 to 0.240) |
| Model 2.4b: Covariate of sex | 3423.6 | 3463.8 | 3397.9 | 0.843 | 0.215 (0.189 to 0.242) |
| Model 2.4c: Covariate of chronic MDD | 3427.3 | 3467.4 | 3401.5 | 0.839 | 0.218 (0.192 to 0.245) |
| Model 2.5: Moderation by episode duration | 3430.1 | 3474.1 | 3401.9 | 0.839 | 0.206 (0.181 to 0.232) |
| **BDI-II, exploratory models (inadequate fit; not interpreted)** | | | | | |

| Model | AIC | BIC | sBIC | CFI | RMSEA (95% CI) |
|---|---|---|---|---|---|
| Model 2.7a: Unconstrained covariance | 3408.7 | 3473.7 | 3367.0 | 0.797 | 0.288 (0.258 to 0.318) |
| Model 2.7b: Constrained covariance | 3409.6 | 3472.7 | 3369.1 | 0.796 | 0.288 (0.258 to 0.318) |
| Model 2.8a: Unconstrained intercept variance | 3408.7 | 3473.7 | 3367.0 | 0.797 | 0.288 (0.258 to 0.318) |
| Model 2.8b: Constrained intercept variance | 3408.4 | 3471.5 | 3367.9 | 0.797 | 0.287 (0.257 to 0.318) |
| Model 2.9a: Unconstrained slope variance | 3408.7 | 3473.7 | 3367.0 | 0.797 | 0.288 (0.258 to 0.318) |
| Model 2.9b: Constrained slope variance | 3407.3 | 3470.4 | 3366.8 | 0.798 | 0.287 (0.257 to 0.317) |
| Model 2.10: Moderation by personality disorder | 3427.0 | 3470.9 | 3398.7 | 0.827 | 0.216 (0.191 to 0.242) |
| Model 2.11: Moderation by childhood trauma | 3425.0 | 3469.0 | 3396.8 | 0.840 | 0.207 (0.182 to 0.233) |
| Model 2.12: Moderation by relationship status | 3430.9 | 3474.9 | 3402.7 | 0.838 | 0.207 (0.182 to 0.233) |
| Model 2.13: Moderation by social network size | 3360.9 | 3404.4 | 3332.2 | 0.850 | 0.197 (0.171 to 0.223) |
| Model 2.14: Moderation by loneliness | 3357.8 | 3401.3 | 3329.1 | 0.846 | 0.201 (0.175 to 0.227) |
| Model 2.15: Moderation by employment status | 3426.8 | 3470.8 | 3398.6 | 0.836 | 0.210 (0.185 to 0.235) |

### B. Linear mixed models of daily BDI-II scores (maximum likelihood)

| | Random intercept | | | Random intercept and slope | | | Slope variance | |
|---|---|---|---|---|---|---|---|---|
| **Model** | **AIC** | **BIC** | **Log-likelihood** | **AIC** | **BIC** | **Log-likelihood** | **LR** | **P** |
| **Confirmatory models** | | | | | | | | |
| Model 2.1: Simple linear model | 3151.3 | 3168.5 | −1571.6 | 3084.2 | 3110.0 | −1536.1 | 71.1 | <.001 |
| Model 2.2: Simple quadratic model | 3150.8 | 3172.4 | −1570.4 | 3082.9 | 3113.1 | −1534.5 | 71.9 | <.001 |
| Model 2.3: Group comparison model | 3148.8 | 3174.6 | −1568.4 | 3086.1 | 3120.6 | −1535.0 | 66.7 | <.001 |
| Model 2.4a: Covariate of age | 3151.6 | 3186.0 | −1567.8 | 3089.2 | 3132.3 | −1534.6 | 66.4 | <.001 |
| Model 2.4b: Covariate of sex | 3145.3 | 3179.7 | −1564.6 | 3082.7 | 3125.8 | −1531.3 | 66.6 | <.001 |
| Model 2.4c: Covariate of chronic MDD | 3143.4 | 3177.9 | −1563.7 | 3085.1 | 3128.2 | −1532.6 | 62.3 | <.001 |
| Model 2.5b: Moderation by episode duration | 3148.8 | 3191.9 | −1564.4 | 3088.9 | 3140.6 | −1532.4 | 63.9 | <.001 |
| Model 2.6: Final combined model | 3139.0 | 3177.8 | −1560.5 | 3080.9 | 3128.3 | −1529.4 | 62.2 | <.001 |
| **Exploratory moderator models** | | | | | | | | |
| Model 2.10: Moderation by personality disorder | 3143.0 | 3186.1 | −1561.5 | 3085.4 | 3137.2 | −1530.7 | 61.5 | <.001 |
| Model 2.11: Moderation by childhood trauma | 3142.8 | 3185.9 | −1561.4 | 3083.5 | 3135.2 | −1529.8 | 63.3 | <.001 |
| Model 2.12: Moderation by relationship status | 3152.2 | 3195.3 | −1566.1 | 3089.7 | 3141.4 | −1532.9 | 66.5 | <.001 |
| Model 2.13: Moderation by social network size | 3087.5 | 3130.4 | −1533.7 | 3028.8 | 3080.3 | −1502.4 | 62.7 | <.001 |
| Model 2.14: Moderation by loneliness | 3086.8 | 3129.7 | −1533.4 | 3025.3 | 3076.8 | −1500.6 | 65.5 | <.001 |
| Model 2.15: Moderation by employment status | 3141.3 | 3184.4 | −1560.6 | 3085.6 | 3137.3 | −1530.8 | 59.7 | <.001 |
| **Exploratory follow-up model** | | | | | | | | |
| Piecewise follow-up model | 3547.8 | 3582.9 | −1765.9 | 3481.5 | 3525.3 | −1730.7 | 70.3 | <.001 |

Abbreviations: AIC, Akaike information criterion; BDI-II, Beck Depression Inventory-II; BIC, Bayesian information criterion; CFI, comparative fit index; LR, likelihood ratio; MADRS, Montgomery-Åsberg Depression Rating Scale; MDD, major depressive disorder; RMSEA, root mean square error of approximation; sBIC, sample-size adjusted BIC.

Note: Lower AIC, BIC, and sBIC values indicate better relative fit among competing models. For the latent growth models, CFI ≥ 0.95 and RMSEA ≤ 0.06 indicate good fit, CFI 0.90 to 0.95 and RMSEA ≤ 0.08 acceptable fit (Hu & Bentler, 1999). The BDI-II latent growth models showed inadequate fit throughout and were replaced by linear mixed models. Panel B: all models fitted with maximum likelihood, so that fit indices are comparable across models; the slope variance was tested against the random-intercept version by a likelihood-ratio test referred to a 50:50 mixture of $\chi^2$ distributions with 1 and 2 degrees of freedom. Exploratory multigroup models 2.7–2.9 were specified as latent growth models only (see Supplementary Methods).

### Table S7. Parameter estimates of the MADRS models

| Parameter | Estimate (95% CI) | SE | z | P |
|---|---|---|---|---|
| **Model 1.1: Linear latent growth model (confirmatory)** | | | | |
| *Latent means* | | | | |
| Baseline mean | 26.10 (23.98 to 28.22) | 1.08 | 24.14 | <.001 |
| Slope mean | −4.40 (−5.53 to −3.28) | 0.57 | −7.68 | <.001 |
| *Variances and covariances* | | | | |
| Baseline variance | 56.04 (30.25 to 81.84) | 13.16 | 4.26 | <.001 |
| Slope variance | 13.26 (3.10 to 23.42) | 5.18 | 2.56 | .01 |
| Intercept–slope covariance | −8.86 (−21.30 to 3.58) | 6.35 | −1.40 | .16 |
| Residual variance (week 0) | 5.53 (−14.60 to 25.67) | 10.27 | 0.54 | .59 |
| Residual variance (week 1) | 14.01 (3.80 to 24.22) | 5.21 | 2.69 | .007 |
| Residual variance (week 2) | 6.99 (−11.50 to 25.48) | 9.44 | 0.74 | .46 |
| **Model 1.2: Non-linear latent growth model (confirmatory)** | | | | |
| *Latent means* | | | | |
| Baseline mean | 26.04 (23.88 to 28.19) | 1.10 | 23.63 | <.001 |
| Slope mean | −3.99 (−5.58 to −2.40) | 0.81 | −4.92 | <.001 |
| *Variances and covariances* | | | | |
| Baseline variance | 54.65 (30.36 to 78.94) | 12.39 | 4.41 | <.001 |
| Slope variance | 11.38 (1.52 to 21.24) | 5.03 | 2.26 | .02 |
| Intercept–slope covariance | −7.39 (−17.94 to 3.16) | 5.38 | −1.37 | .17 |
| Residual variance (week 0) | 6.92 (−11.88 to 25.73) | 9.59 | 0.72 | .47 |
| Residual variance (week 1) | 14.34 (4.28 to 24.40) | 5.13 | 2.79 | .005 |
| Residual variance (week 2) | 3.05 (−19.57 to 25.66) | 11.54 | 0.26 | .79 |
| *Estimated factor loadings* | | | | |
| Non-linear slope loading (week 2) | 2.21 (1.57 to 2.85) | 0.33 | 6.78 | <.001 |
| **Model 1.3: Group comparison model (confirmatory)** | | | | |
| *Regressions on baseline* | | | | |
| Group | 5.10 (1.00 to 9.20) | 2.09 | 2.44 | .01 |
| *Regressions on slope* | | | | |
| Group | −3.54 (−5.53 to −1.55) | 1.02 | −3.49 | <.001 |
| *Latent means* | | | | |
| Baseline mean | 23.30 (20.22 to 26.38) | 1.57 | 14.83 | <.001 |
| Slope mean | −2.49 (−3.53 to −1.45) | 0.53 | −4.70 | <.001 |
| *Variances and covariances* | | | | |
| Baseline variance | 52.68 (27.59 to 77.77) | 12.80 | 4.12 | <.001 |

| Parameter | Estimate (95% CI) | SE | z | P |
|---|---|---|---|---|
| Slope variance | 11.06 (1.17 to 20.94) | 5.04 | 2.19 | .03 |
| Intercept–slope covariance | −6.38 (−18.86 to 6.11) | 6.37 | −1.00 | .32 |
| Residual variance (week 0) | 1.91 (−17.11 to 20.93) | 9.70 | 0.20 | .84 |
| Residual variance (week 1) | 14.58 (4.25 to 24.91) | 5.27 | 2.77 | .006 |
| Residual variance (week 2) | 8.34 (−10.21 to 26.89) | 9.46 | 0.88 | .38 |
| **Model 1.3b: Group model adjusting for baseline differences (sensitivity, not preregistered)** | | | | |
| *Regressions on baseline* | | | | |
| Group | 5.10 (1.00 to 9.20) | 2.09 | 2.44 | .01 |
| *Regressions on slope* | | | | |
| Group | −2.92 (−4.99 to −0.85) | 1.06 | −2.76 | .006 |
| Baseline | −0.12 (−0.32 to 0.08) | 0.10 | −1.18 | .24 |
| *Latent means* | | | | |
| Baseline mean | 23.30 (20.22 to 26.38) | 1.57 | 14.83 | <.001 |
| Slope mean | 0.33 (−4.46 to 5.12) | 2.44 | 0.13 | .89 |
| *Variances and covariances* | | | | |
| Baseline variance | 52.68 (27.59 to 77.77) | 12.80 | 4.12 | <.001 |
| Slope variance | 10.29 (2.14 to 18.43) | 4.16 | 2.48 | .01 |
| Residual variance (week 0) | 1.91 (−17.11 to 20.93) | 9.70 | 0.20 | .84 |
| Residual variance (week 1) | 14.58 (4.25 to 24.91) | 5.27 | 2.77 | .006 |
| Residual variance (week 2) | 8.34 (−10.21 to 26.89) | 9.46 | 0.88 | .38 |
| **Model 1.4a: Covariate of age (confirmatory)** | | | | |
| *Regressions on baseline* | | | | |
| Group | 5.15 (1.08 to 9.23) | 2.08 | 2.48 | .01 |
| Age | 0.08 (−0.08 to 0.23) | 0.08 | 0.97 | .33 |
| *Regressions on slope* | | | | |
| Group | −3.51 (−5.47 to −1.55) | 1.00 | −3.50 | <.001 |
| Age | 0.02 (−0.06 to 0.09) | 0.04 | 0.47 | .63 |
| *Latent means* | | | | |
| Baseline mean | 20.15 (13.02 to 27.28) | 3.64 | 5.54 | <.001 |
| Slope mean | −3.22 (−6.49 to 0.05) | 1.67 | −1.93 | .054 |
| *Variances and covariances* | | | | |
| Baseline variance | 52.35 (26.35 to 78.35) | 13.27 | 3.95 | <.001 |
| Slope variance | 11.40 (1.55 to 21.26) | 5.03 | 2.27 | .02 |
| Intercept–slope covariance | −6.95 (−19.76 to 5.86) | 6.54 | −1.06 | .29 |
| Residual variance (week 0) | 1.25 (−17.84 to 20.33) | 9.74 | 0.13 | .90 |
| Residual variance (week 1) | 14.99 (4.37 to 25.61) | 5.42 | 2.77 | .006 |

| Parameter | Estimate (95% CI) | SE | z | P |
|---|---|---|---|---|
| Residual variance (week 2) | 7.36 (−11.24 to 25.96) | 9.49 | 0.78 | .44 |
| **Model 1.4b: Covariate of sex (confirmatory)** | | | | |
| *Regressions on baseline* | | | | |
| Group | 4.95 (1.11 to 8.79) | 1.96 | 2.53 | .01 |
| Sex | −4.66 (−8.10 to −1.21) | 1.76 | −2.65 | .008 |
| *Regressions on slope* | | | | |
| Group | −3.56 (−5.55 to −1.57) | 1.02 | −3.50 | <.001 |
| Sex | −0.06 (−2.12 to 2.00) | 1.05 | −0.06 | .95 |
| *Latent means* | | | | |
| Baseline mean | 30.15 (25.47 to 34.83) | 2.39 | 12.62 | <.001 |
| Slope mean | −2.37 (−5.40 to 0.66) | 1.55 | −1.53 | .13 |
| *Variances and covariances* | | | | |
| Baseline variance | 43.12 (23.99 to 62.25) | 9.76 | 4.42 | <.001 |
| Slope variance | 9.12 (−0.09 to 18.33) | 4.70 | 1.94 | .052 |
| Intercept–slope covariance | −4.33 (−14.98 to 6.31) | 5.43 | −0.80 | .42 |
| Residual variance (week 0) | 5.77 (−11.50 to 23.03) | 8.81 | 0.65 | .51 |
| Residual variance (week 1) | 12.59 (3.50 to 21.69) | 4.64 | 2.71 | .007 |
| Residual variance (week 2) | 12.40 (−5.32 to 30.13) | 9.05 | 1.37 | .17 |
| **Model 1.4c: Covariate of chronic MDD (confirmatory)** | | | | |
| *Regressions on baseline* | | | | |
| Group | 3.51 (−0.35 to 7.37) | 1.97 | 1.78 | .07 |
| Chronic MDD | 5.44 (0.04 to 10.85) | 2.76 | 1.97 | .048 |
| *Regressions on slope* | | | | |
| Group | −3.67 (−5.74 to −1.60) | 1.06 | −3.47 | <.001 |
| Chronic MDD | 0.29 (−1.67 to 2.26) | 1.00 | 0.29 | .77 |
| *Latent means* | | | | |
| Baseline mean | 19.84 (14.54 to 25.14) | 2.70 | 7.34 | <.001 |
| Slope mean | −2.64 (−4.15 to −1.12) | 0.77 | −3.42 | <.001 |
| *Variances and covariances* | | | | |
| Baseline variance | 47.57 (25.69 to 69.45) | 11.16 | 4.26 | <.001 |
| Slope variance | 10.70 (1.44 to 19.96) | 4.72 | 2.27 | .02 |
| Intercept–slope covariance | −6.25 (−17.51 to 5.01) | 5.75 | −1.09 | .28 |
| Residual variance (week 0) | 2.60 (−14.78 to 19.97) | 8.87 | 0.29 | .77 |
| Residual variance (week 1) | 14.18 (4.70 to 23.66) | 4.84 | 2.93 | .003 |
| Residual variance (week 2) | 9.26 (−8.58 to 27.09) | 9.10 | 1.02 | .31 |
| **Model 1.5: Moderation by episode duration (confirmatory)** | | | | |

| Parameter | Estimate (95% CI) | SE | z | P |
|---|---|---|---|---|
| *Regressions on baseline* | | | | |
| Group | 4.00 (−0.24 to 8.24) | 2.16 | 1.85 | .06 |
| Duration (centered) | 3.96 (−0.92 to 8.85) | 2.49 | 1.59 | .11 |
| Duration × Group (centered) | −2.72 (−12.54 to 7.09) | 5.01 | −0.54 | .59 |
| *Regressions on slope* | | | | |
| Group | −3.93 (−5.95 to −1.90) | 1.03 | −3.80 | <.001 |
| Duration (centered) | 1.33 (−1.07 to 3.73) | 1.23 | 1.09 | .28 |
| Duration × Group (centered) | 2.94 (−1.87 to 7.76) | 2.46 | 1.20 | .23 |
| *Latent means* | | | | |
| Baseline mean | 26.05 (23.99 to 28.11) | 1.05 | 24.79 | <.001 |
| Slope mean | −4.45 (−5.47 to −3.44) | 0.52 | −8.60 | <.001 |
| *Variances and covariances* | | | | |
| Baseline variance | 49.35 (26.97 to 71.74) | 11.42 | 4.32 | <.001 |
| Slope variance | 10.60 (1.14 to 20.07) | 4.83 | 2.20 | .03 |
| Intercept–slope covariance | −6.60 (−18.65 to 5.44) | 6.14 | −1.07 | .28 |
| Residual variance (week 0) | 2.01 (−16.71 to 20.73) | 9.55 | 0.21 | .83 |
| Residual variance (week 1) | 14.47 (4.45 to 24.50) | 5.11 | 2.83 | .005 |
| Residual variance (week 2) | 8.66 (−8.46 to 25.77) | 8.73 | 0.99 | .32 |
| **Model 1.6: Final combined model (confirmatory)** | | | | |
| *Regressions on baseline* | | | | |
| Group | 3.62 (−0.21 to 7.46) | 1.96 | 1.85 | .06 |
| Chronic MDD | 4.66 (0.23 to 9.08) | 2.26 | 2.06 | .04 |
| Sex | −4.11 (−7.18 to −1.05) | 1.56 | −2.63 | .009 |
| *Regressions on slope* | | | | |
| Group | −3.60 (−5.60 to −1.60) | 1.02 | −3.52 | <.001 |
| *Latent means* | | | | |
| Baseline mean | 26.38 (20.37 to 32.38) | 3.06 | 8.61 | <.001 |
| Slope mean | −2.42 (−3.49 to −1.35) | 0.55 | −4.43 | <.001 |
| *Variances and covariances* | | | | |
| Baseline variance | 39.93 (22.51 to 57.35) | 8.89 | 4.49 | <.001 |
| Slope variance | 9.10 (0.40 to 17.80) | 4.44 | 2.05 | .04 |
| Intercept–slope covariance | −4.44 (−14.23 to 5.35) | 5.00 | −0.89 | .37 |
| Residual variance (week 0) | 5.92 (−10.14 to 21.97) | 8.19 | 0.72 | .47 |
| Residual variance (week 1) | 12.51 (4.01 to 21.00) | 4.33 | 2.89 | .004 |
| Residual variance (week 2) | 12.63 (−4.45 to 29.71) | 8.71 | 1.45 | .15 |
| **Model 1.7a: Unconstrained covariance (exploratory)** | | | | |

| Parameter | Estimate (95% CI) | SE | z | P |
|---|---|---|---|---|
| *Latent means* | | | | |
| Baseline mean [Sham] | 23.21 (20.23 to 26.18) | 1.52 | 15.29 | <.001 |
| Baseline mean [Active] | 28.36 (25.71 to 31.01) | 1.35 | 20.98 | <.001 |
| Slope mean [Sham] | −2.60 (−3.60 to −1.60) | 0.51 | −5.11 | <.001 |
| Slope mean [Active] | −6.05 (−7.74 to −4.35) | 0.86 | −7.00 | <.001 |
| *Variances and covariances* | | | | |
| Baseline variance [Sham] | 64.72 (18.41 to 111.03) | 23.63 | 2.74 | .006 |
| Baseline variance [Active] | 42.28 (17.75 to 66.80) | 12.51 | 3.38 | <.001 |
| Slope variance [Sham] | 7.93 (−8.28 to 24.15) | 8.27 | 0.96 | .34 |
| Slope variance [Active] | 14.12 (2.02 to 26.21) | 6.17 | 2.29 | .02 |
| Intercept–slope covariance [Sham] | −10.87 (−30.18 to 8.45) | 9.86 | −1.10 | .27 |
| Intercept–slope covariance [Active] | −2.38 (−18.66 to 13.90) | 8.30 | −0.29 | .77 |
| Residual variance (week 0) [Sham] | −3.49 (−35.81 to 28.84) | 16.49 | −0.21 | .83 |
| Residual variance (week 0) [Active] | 6.48 (−15.48 to 28.45) | 11.21 | 0.58 | .56 |
| Residual variance (week 1) [Sham] | 20.88 (4.60 to 37.17) | 8.31 | 2.51 | .01 |
| Residual variance (week 1) [Active] | 9.17 (−3.79 to 22.13) | 6.61 | 1.39 | .17 |
| Residual variance (week 2) [Sham] | −1.35 (−30.89 to 28.18) | 15.07 | −0.09 | .93 |
| Residual variance (week 2) [Active] | 16.43 (−6.44 to 39.30) | 11.67 | 1.41 | .16 |
| **Model 1.7b: Constrained covariance (exploratory)** | | | | |
| *Latent means* | | | | |
| Baseline mean [Sham] | 23.38 (20.34 to 26.42) | 1.55 | 15.09 | <.001 |
| Baseline mean [Active] | 28.39 (25.73 to 31.04) | 1.36 | 20.92 | <.001 |
| Slope mean [Sham] | −2.62 (−3.63 to −1.62) | 0.51 | −5.14 | <.001 |
| Slope mean [Active] | −6.05 (−7.74 to −4.36) | 0.86 | −7.01 | <.001 |
| *Variances and covariances* | | | | |
| Baseline variance [Sham] | 56.72 (22.57 to 90.87) | 17.43 | 3.25 | .001 |
| Baseline variance [Active] | 47.33 (21.37 to 73.29) | 13.25 | 3.57 | <.001 |
| Slope variance [Sham] | 5.05 (−8.45 to 18.54) | 6.88 | 0.73 | .46 |
| Slope variance [Active] | 16.50 (5.60 to 27.41) | 5.56 | 2.97 | .003 |
| Intercept–slope covariance [Sham] | −6.76 (−19.56 to 6.03) | 6.53 | −1.04 | .30 |
| Intercept–slope covariance [Active] | −6.76 (−19.56 to 6.03) | 6.53 | −1.04 | .30 |
| Residual variance (week 0) [Sham] | 2.56 (−22.41 to 27.54) | 12.74 | 0.20 | .84 |
| Residual variance (week 0) [Active] | 2.84 (−17.08 to 22.76) | 10.16 | 0.28 | .78 |
| Residual variance (week 1) [Sham] | 18.39 (5.94 to 30.83) | 6.35 | 2.90 | .004 |
| Residual variance (week 1) [Active] | 10.65 (−1.67 to 22.98) | 6.29 | 1.69 | .09 |
| Residual variance (week 2) [Sham] | 3.09 (−24.55 to 30.72) | 14.10 | 0.22 | .83 |

| Parameter | Estimate (95% CI) | SE | z | P |
|---|---|---|---|---|
| Residual variance (week 2) [Active] | 13.72 (−6.11 to 33.56) | 10.12 | 1.36 | .17 |
| **Model 1.8a: Unconstrained intercept variance (exploratory)** | | | | |
| *Latent means* | | | | |
| Baseline mean [Sham] | 23.21 (20.23 to 26.18) | 1.52 | 15.29 | <.001 |
| Baseline mean [Active] | 28.36 (25.71 to 31.01) | 1.35 | 20.98 | <.001 |
| Slope mean [Sham] | −2.60 (−3.60 to −1.60) | 0.51 | −5.11 | <.001 |
| Slope mean [Active] | −6.05 (−7.74 to −4.35) | 0.86 | −7.00 | <.001 |
| *Variances and covariances* | | | | |
| Baseline variance [Sham] | 64.72 (18.41 to 111.03) | 23.63 | 2.74 | .006 |
| Baseline variance [Active] | 42.28 (17.75 to 66.80) | 12.51 | 3.38 | <.001 |
| Slope variance [Sham] | 7.93 (−8.28 to 24.15) | 8.27 | 0.96 | .34 |
| Slope variance [Active] | 14.12 (2.02 to 26.21) | 6.17 | 2.29 | .02 |
| Intercept–slope covariance [Sham] | −10.87 (−30.18 to 8.45) | 9.86 | −1.10 | .27 |
| Intercept–slope covariance [Active] | −2.38 (−18.66 to 13.90) | 8.30 | −0.29 | .77 |
| Residual variance (week 0) [Sham] | −3.49 (−35.81 to 28.84) | 16.49 | −0.21 | .83 |
| Residual variance (week 0) [Active] | 6.48 (−15.48 to 28.45) | 11.21 | 0.58 | .56 |
| Residual variance (week 1) [Sham] | 20.88 (4.60 to 37.17) | 8.31 | 2.51 | .01 |
| Residual variance (week 1) [Active] | 9.17 (−3.79 to 22.13) | 6.61 | 1.39 | .17 |
| Residual variance (week 2) [Sham] | −1.35 (−30.89 to 28.18) | 15.07 | −0.09 | .93 |
| Residual variance (week 2) [Active] | 16.43 (−6.44 to 39.30) | 11.67 | 1.41 | .16 |
| **Model 1.8b: Constrained intercept variance (exploratory)** | | | | |
| *Latent means* | | | | |
| Baseline mean [Sham] | 23.36 (20.43 to 26.29) | 1.49 | 15.63 | <.001 |
| Baseline mean [Active] | 28.38 (25.71 to 31.05) | 1.36 | 20.85 | <.001 |
| Slope mean [Sham] | −2.62 (−3.63 to −1.62) | 0.51 | −5.13 | <.001 |
| Slope mean [Active] | −6.05 (−7.75 to −4.35) | 0.86 | −6.99 | <.001 |
| *Variances and covariances* | | | | |
| Baseline variance [Sham] | 51.95 (26.88 to 77.02) | 12.79 | 4.06 | <.001 |
| Baseline variance [Active] | 51.95 (26.88 to 77.02) | 12.79 | 4.06 | <.001 |
| Slope variance [Sham] | 5.38 (−8.40 to 19.15) | 7.03 | 0.76 | .44 |
| Slope variance [Active] | 15.75 (3.40 to 28.09) | 6.30 | 2.50 | .01 |
| Intercept–slope covariance [Sham] | −6.94 (−21.04 to 7.15) | 7.19 | −0.97 | .33 |
| Intercept–slope covariance [Active] | −5.16 (−22.39 to 12.07) | 8.79 | −0.59 | .56 |
| Residual variance (week 0) [Sham] | 2.11 (−23.62 to 27.84) | 13.13 | 0.16 | .87 |
| Residual variance (week 0) [Active] | 3.20 (−20.13 to 26.54) | 11.90 | 0.27 | .79 |
| Residual variance (week 1) [Sham] | 18.61 (6.27 to 30.95) | 6.30 | 2.96 | .003 |

| Parameter | Estimate (95% CI) | SE | z | P |
|---|---|---|---|---|
| Residual variance (week 1) [Active] | 10.52 (−3.48 to 24.52) | 7.14 | 1.47 | .14 |
| Residual variance (week 2) [Sham] | 2.56 (−24.30 to 29.42) | 13.70 | 0.19 | .85 |
| Residual variance (week 2) [Active] | 13.91 (−9.09 to 36.92) | 11.74 | 1.19 | .24 |
| **Model 1.9a: Unconstrained slope variance (exploratory)** | | | | |
| *Latent means* | | | | |
| Baseline mean [Sham] | 23.21 (20.23 to 26.18) | 1.52 | 15.29 | <.001 |
| Baseline mean [Active] | 28.36 (25.71 to 31.01) | 1.35 | 20.98 | <.001 |
| Slope mean [Sham] | −2.60 (−3.60 to −1.60) | 0.51 | −5.11 | <.001 |
| Slope mean [Active] | −6.05 (−7.74 to −4.35) | 0.86 | −7.00 | <.001 |
| *Variances and covariances* | | | | |
| Baseline variance [Sham] | 64.72 (18.41 to 111.03) | 23.63 | 2.74 | .006 |
| Baseline variance [Active] | 42.28 (17.75 to 66.80) | 12.51 | 3.38 | <.001 |
| Slope variance [Sham] | 7.93 (−8.28 to 24.15) | 8.27 | 0.96 | .34 |
| Slope variance [Active] | 14.12 (2.02 to 26.21) | 6.17 | 2.29 | .02 |
| Intercept–slope covariance [Sham] | −10.87 (−30.18 to 8.45) | 9.86 | −1.10 | .27 |
| Intercept–slope covariance [Active] | −2.38 (−18.66 to 13.90) | 8.30 | −0.29 | .77 |
| Residual variance (week 0) [Sham] | −3.49 (−35.81 to 28.84) | 16.49 | −0.21 | .83 |
| Residual variance (week 0) [Active] | 6.48 (−15.48 to 28.45) | 11.21 | 0.58 | .56 |
| Residual variance (week 1) [Sham] | 20.88 (4.60 to 37.17) | 8.31 | 2.51 | .01 |
| Residual variance (week 1) [Active] | 9.17 (−3.79 to 22.13) | 6.61 | 1.39 | .17 |
| Residual variance (week 2) [Sham] | −1.35 (−30.89 to 28.18) | 15.07 | −0.09 | .93 |
| Residual variance (week 2) [Active] | 16.43 (−6.44 to 39.30) | 11.67 | 1.41 | .16 |
| **Model 1.9b: Constrained slope variance (exploratory)** | | | | |
| *Latent means* | | | | |
| Baseline mean [Active] | 28.34 (25.69 to 30.99) | 1.35 | 20.96 | <.001 |
| Baseline mean [Sham] | 23.04 (20.03 to 26.05) | 1.54 | 14.99 | <.001 |
| Slope mean [Active] | −6.05 (−7.74 to −4.36) | 0.86 | −7.00 | <.001 |
| Slope mean [Sham] | −2.59 (−3.60 to −1.58) | 0.51 | −5.03 | <.001 |
| *Variances and covariances* | | | | |
| Baseline variance [Active] | 39.90 (17.10 to 62.70) | 11.63 | 3.43 | <.001 |
| Baseline variance [Sham] | 72.20 (27.39 to 117.01) | 22.86 | 3.16 | .002 |
| Slope variance [Active] | 10.89 (0.22 to 21.57) | 5.44 | 2.00 | .045 |
| Slope variance [Sham] | 10.89 (0.22 to 21.57) | 5.44 | 2.00 | .045 |
| Intercept–slope covariance [Active] | 0.00 (−15.19 to 15.20) | 7.75 | 0.00 | >.99 |
| Intercept–slope covariance [Sham] | −14.22 (−30.50 to 2.06) | 8.31 | −1.71 | .09 |
| Residual variance (week 0) [Active] | 9.68 (−10.84 to 30.21) | 10.47 | 0.92 | .36 |

| Parameter | Estimate (95% CI) | SE | z | P |
|---|---|---|---|---|
| Residual variance (week 0) [Sham] | −9.27 (−33.10 to 14.56) | 12.16 | −0.76 | .45 |
| Residual variance (week 1) [Active] | 7.75 (−4.22 to 19.72) | 6.11 | 1.27 | .20 |
| Residual variance (week 1) [Sham] | 23.98 (10.43 to 37.54) | 6.91 | 3.47 | <.001 |
| Residual variance (week 2) [Active] | 20.78 (−1.75 to 43.30) | 11.49 | 1.81 | .07 |
| Residual variance (week 2) [Sham] | −6.42 (−27.89 to 15.05) | 10.95 | −0.59 | .56 |
| **Model 1.10: Moderation by personality disorder (exploratory)** | | | | |
| *Regressions on baseline* | | | | |
| Group | 5.39 (0.43 to 10.34) | 2.53 | 2.13 | .03 |
| Personality disorder | 6.18 (1.44 to 10.92) | 2.42 | 2.55 | .01 |
| Personality disorder × Group | 0.46 (−5.53 to 6.45) | 3.06 | 0.15 | .88 |
| *Regressions on slope* | | | | |
| Group | −4.12 (−6.56 to −1.67) | 1.25 | −3.30 | <.001 |
| Personality disorder | −0.94 (−2.98 to 1.10) | 1.04 | −0.91 | .36 |
| Personality disorder × Group | 1.99 (−2.04 to 6.01) | 2.05 | 0.97 | .33 |
| *Latent means* | | | | |
| Baseline mean | 21.52 (17.58 to 25.45) | 2.01 | 10.72 | <.001 |
| Slope mean | −2.17 (−3.51 to −0.82) | 0.69 | −3.16 | .002 |
| *Variances and covariances* | | | | |
| Baseline variance | 42.55 (18.01 to 67.09) | 12.52 | 3.40 | <.001 |
| Slope variance | 9.39 (−0.12 to 18.90) | 4.85 | 1.94 | .053 |
| Intercept–slope covariance | −4.99 (−17.38 to 7.40) | 6.32 | −0.79 | .43 |
| Residual variance (week 0) | 4.49 (−13.61 to 22.59) | 9.24 | 0.49 | .63 |
| Residual variance (week 1) | 13.00 (3.08 to 22.91) | 5.06 | 2.57 | .01 |
| Residual variance (week 2) | 12.07 (−5.77 to 29.90) | 9.10 | 1.33 | .18 |
| **Model 1.11: Moderation by childhood trauma (exploratory)** | | | | |
| *Regressions on baseline* | | | | |
| Group | 3.42 (−0.13 to 6.98) | 1.81 | 1.89 | .06 |
| Childhood trauma (centered) | 32.91 (4.73 to 61.10) | 14.38 | 2.29 | .02 |
| Childhood trauma × Group (centered) | −20.97 (−55.65 to 13.71) | 17.69 | −1.18 | .24 |
| *Regressions on slope* | | | | |
| Group | −3.21 (−5.11 to −1.32) | 0.97 | −3.32 | <.001 |
| Childhood trauma (centered) | −7.41 (−15.73 to 0.92) | 4.25 | −1.74 | .08 |
| Childhood trauma × Group (centered) | 4.62 (−12.58 to 21.82) | 8.77 | 0.53 | .60 |
| *Latent means* | | | | |
| Baseline mean | 24.52 (22.13 to 26.92) | 1.22 | 20.08 | <.001 |
| Slope mean | −2.71 (−3.70 to −1.72) | 0.50 | −5.37 | <.001 |

| Parameter | Estimate (95% CI) | SE | z | P |
|---|---|---|---|---|
| *Variances and covariances* | | | | |
| Baseline variance | 41.76 (23.25 to 60.28) | 9.45 | 4.42 | <.001 |
| Slope variance | 9.47 (0.77 to 18.17) | 4.44 | 2.13 | .03 |
| Intercept–slope covariance | −3.01 (−12.77 to 6.75) | 4.98 | −0.60 | .55 |
| Residual variance (week 0) | 5.28 (−10.12 to 20.68) | 7.86 | 0.67 | .50 |
| Residual variance (week 1) | 13.24 (4.17 to 22.30) | 4.62 | 2.86 | .004 |
| Residual variance (week 2) | 10.33 (−9.25 to 29.92) | 9.99 | 1.03 | .30 |
| **Model 1.12: Moderation by relationship status (exploratory)** | | | | |
| *Regressions on baseline* | | | | |
| Group | 7.17 (0.96 to 13.39) | 3.17 | 2.26 | .02 |
| Relationship | 6.78 (1.20 to 12.36) | 2.85 | 2.38 | .02 |
| Relationship × Group | −5.70 (−13.58 to 2.18) | 4.02 | −1.42 | .16 |
| *Regressions on slope* | | | | |
| Group | −1.92 (−4.97 to 1.14) | 1.56 | −1.23 | .22 |
| Relationship | −0.27 (−2.62 to 2.08) | 1.20 | −0.22 | .82 |
| Relationship × Group | −2.57 (−6.77 to 1.63) | 2.14 | −1.20 | .23 |
| *Latent means* | | | | |
| Baseline mean | 20.58 (16.38 to 24.77) | 2.14 | 9.62 | <.001 |
| Slope mean | −2.46 (−3.60 to −1.33) | 0.58 | −4.24 | <.001 |
| *Variances and covariances* | | | | |
| Baseline variance | 47.23 (26.96 to 67.50) | 10.34 | 4.57 | <.001 |
| Slope variance | 10.33 (2.05 to 18.62) | 4.23 | 2.44 | .01 |
| Intercept–slope covariance | −5.71 (−16.24 to 4.83) | 5.38 | −1.06 | .29 |
| Residual variance (week 0) | 2.21 (−15.24 to 19.65) | 8.90 | 0.25 | .80 |
| Residual variance (week 1) | 14.99 (5.28 to 24.70) | 4.95 | 3.03 | .002 |
| Residual variance (week 2) | 6.39 (−9.76 to 22.53) | 8.24 | 0.78 | .44 |
| **Model 1.13: Moderation by social network size (exploratory)** | | | | |
| *Regressions on baseline* | | | | |
| Group | 4.86 (0.98 to 8.74) | 1.98 | 2.46 | .01 |
| Social network (centered) | −9.59 (−20.76 to 1.58) | 5.70 | −1.68 | .09 |
| Social network × Group (centered) | 1.66 (−12.13 to 15.45) | 7.04 | 0.24 | .81 |
| *Regressions on slope* | | | | |
| Group | −3.59 (−5.61 to −1.56) | 1.03 | −3.47 | <.001 |
| Social network (centered) | 1.04 (−2.98 to 5.06) | 2.05 | 0.51 | .61 |
| Social network × Group (centered) | −0.78 (−8.50 to 6.94) | 3.94 | −0.20 | .84 |
| *Latent means* | | | | |

| Parameter | Estimate (95% CI) | SE | z | P |
|---|---|---|---|---|
| Baseline mean | 23.43 (20.52 to 26.34) | 1.49 | 15.77 | <.001 |
| Slope mean | −2.45 (−3.54 to −1.36) | 0.56 | −4.40 | <.001 |
| *Variances and covariances* | | | | |
| Baseline variance | 52.43 (26.03 to 78.83) | 13.47 | 3.89 | <.001 |
| Slope variance | 13.70 (3.45 to 23.96) | 5.23 | 2.62 | .009 |
| Intercept–slope covariance | −8.89 (−21.76 to 3.98) | 6.57 | −1.35 | .18 |
| Residual variance (week 0) | −3.01 (−22.29 to 16.27) | 9.84 | −0.31 | .76 |
| Residual variance (week 1) | 16.20 (5.55 to 26.84) | 5.43 | 2.98 | .003 |
| Residual variance (week 2) | 4.14 (−15.40 to 23.67) | 9.97 | 0.41 | .68 |
| **Model 1.14: Moderation by loneliness (exploratory)** | | | | |
| *Regressions on baseline* | | | | |
| Group | 5.17 (1.36 to 8.99) | 1.95 | 2.66 | .008 |
| Loneliness (centered) | −0.00 (−0.32 to 0.32) | 0.16 | −0.02 | .98 |
| Loneliness × Group (centered) | 0.39 (0.01 to 0.78) | 0.19 | 2.03 | .04 |
| *Regressions on slope* | | | | |
| Group | −3.46 (−5.53 to −1.40) | 1.05 | −3.29 | .001 |
| Loneliness (centered) | 0.06 (−0.07 to 0.18) | 0.06 | 0.89 | .37 |
| Loneliness × Group (centered) | −0.06 (−0.27 to 0.16) | 0.11 | −0.51 | .61 |
| *Latent means* | | | | |
| Baseline mean | 23.44 (20.25 to 26.62) | 1.62 | 14.43 | <.001 |
| Slope mean | −2.54 (−3.61 to −1.47) | 0.55 | −4.65 | <.001 |
| *Variances and covariances* | | | | |
| Baseline variance | 45.39 (21.50 to 69.28) | 12.19 | 3.72 | <.001 |
| Slope variance | 11.82 (2.49 to 21.16) | 4.76 | 2.48 | .01 |
| Intercept–slope covariance | −6.94 (−18.23 to 4.35) | 5.76 | −1.20 | .23 |
| Residual variance (week 0) | 1.09 (−16.54 to 18.73) | 9.00 | 0.12 | .90 |
| Residual variance (week 1) | 14.57 (4.87 to 24.26) | 4.95 | 2.94 | .003 |
| Residual variance (week 2) | 6.54 (−10.38 to 23.47) | 8.64 | 0.76 | .45 |
| **Model 1.15: Moderation by employment status (exploratory)** | | | | |
| *Regressions on baseline* | | | | |
| Group | 2.03 (−2.82 to 6.89) | 2.48 | 0.82 | .41 |
| Employment | −9.19 (−14.14 to −4.25) | 2.52 | −3.64 | <.001 |
| Employment × Group | 6.93 (−0.14 to 14.01) | 3.61 | 1.92 | .055 |
| *Regressions on slope* | | | | |
| Group | −2.90 (−5.27 to −0.53) | 1.21 | −2.40 | .02 |
| Employment | 2.50 (0.43 to 4.57) | 1.06 | 2.37 | .02 |

| Parameter | Estimate (95% CI) | SE | z | P |
|---|---|---|---|---|
| Employment × Group | −1.51 (−5.45 to 2.43) | 2.01 | −0.75 | .45 |
| *Latent means* | | | | |
| Baseline mean | 27.36 (24.14 to 30.58) | 1.64 | 16.64 | <.001 |
| Slope mean | −3.55 (−4.31 to −2.79) | 0.39 | −9.16 | <.001 |
| *Variances and covariances* | | | | |
| Baseline variance | 41.12 (22.89 to 59.35) | 9.30 | 4.42 | <.001 |
| Slope variance | 9.90 (−0.17 to 19.97) | 5.14 | 1.93 | .054 |
| Intercept–slope covariance | −2.99 (−14.30 to 8.31) | 5.77 | −0.52 | .60 |
| Residual variance (week 0) | 2.84 (−15.43 to 21.11) | 9.32 | 0.30 | .76 |
| Residual variance (week 1) | 14.13 (4.04 to 24.22) | 5.15 | 2.75 | .006 |
| Residual variance (week 2) | 9.20 (−9.45 to 27.85) | 9.51 | 0.97 | .33 |
| **Piecewise follow-up model (linear mixed model with random intercept; time in weeks) (exploratory)** | | | | |
| **Parameter** | **Estimate (95% CI)** | **SE** | **t (df)** | **P** |
| *Random effects* | | | | |
| Intercept (SD) | 6.96 (5.49 to 8.60) | — | — | — |
| Residual (SD) | 5.11 (4.50 to 5.69) | — | — | — |
| *Fixed effects* | | | | |
| Intercept | 23.43 (19.98 to 26.87) | 1.73 | 13.55 (79.1) | <.001 |
| Time (Acute phase) | −2.53 (−4.01 to −1.04) | 0.75 | −3.37 (138.6) | <.001 |
| Group (Active) | 4.92 (0.21 to 9.63) | 2.37 | 2.08 (78.0) | .04 |
| Time (Follow-up phase) | −0.11 (−0.60 to 0.38) | 0.25 | −0.45 (139.1) | .66 |
| Time (Acute phase) × Group | −3.43 (−5.47 to −1.40) | 1.03 | −3.34 (138.6) | .001 |
| Time (Follow-up phase) × Group | 0.39 (−0.29 to 1.07) | 0.34 | 1.14 (139.6) | .26 |

Abbreviations: CI, confidence interval; MADRS, Montgomery-Åsberg Depression Rating Scale; MDD, major depressive disorder; SE, standard error.

Note: Latent growth models (Models 1.1–1.15) were estimated with robust maximum likelihood; time was coded in weeks (0, 1, 2), so slope estimates are changes in MADRS score per week; the test statistic is the Wald z. Group was coded 0 (sham) and 1 (active); sex 0 (female) and 1 (male); continuous moderators were mean-centered and, where skewed, log-transformed. In Model 1.5, group was effect-coded (sham, −0.5; active, +0.5), so that the duration effects and latent means refer to the average of both groups. Bracketed labels in the multigroup models (1.7–1.9) denote group-specific parameters. Models 1.7a, 1.8a, 1.9a, 1.9b, and 1.13 yielded negative residual variance estimates (Heywood cases); their variance and covariance estimates should be interpreted with caution. Piecewise follow-up model: fixed effects with t tests and Satterthwaite degrees of freedom (in parentheses); slopes are changes in MADRS score per week in the acute phase (weeks 0–2) and in the follow-up phase (weeks 2–8).

**Table S8. Parameter estimates of the linear mixed models of daily BDI-II scores**

| | Random intercept | | | Random intercept and slope | | |
|---|---|---|---|---|---|---|
| Parameter | Estimate (95% CI) | t (df) | P | Estimate (95% CI) | t (df) | P |
| **Model 2.1: Simple linear model (confirmatory)** | | | | | | |
| *Random effects* | | | | | | |
| Intercept (SD) | 10.45 (8.59 to 12.78) | — | — | 10.62 (8.70 to 13.02) | — | — |
| Slope, time (SD) | — | — | — | 0.52 (0.40 to 0.67) | — | — |
| Intercept–slope correlation | — | — | — | −0.18 (−0.46 to 0.13) | — | — |
| Residual (SD) | 3.42 (3.21 to 3.64) | — | — | 2.96 (2.77 to 3.16) | — | — |
| *Fixed effects* | | | | | | |
| Intercept | 26.12 (23.10 to 29.14) | 17.38 (51.4) | <.001 | 26.12 (23.06 to 29.18) | 17.17 (49.0) | <.001 |
| Time | −0.31 (−0.40 to −0.21) | −6.62 (499.0) | <.001 | −0.31 (−0.47 to −0.14) | −3.63 (49.0) | <.001 |
| **Model 2.2: Simple quadratic model (confirmatory)** | | | | | | |
| *Random effects* | | | | | | |
| Intercept (SD) | 10.45 (8.59 to 12.78) | — | — | 10.62 (8.70 to 13.02) | — | — |
| Slope, time (SD) | — | — | — | 0.52 (0.40 to 0.67) | — | — |
| Intercept–slope correlation | — | — | — | −0.18 (−0.46 to 0.13) | — | — |
| Residual (SD) | 3.41 (3.21 to 3.63) | — | — | 2.95 (2.76 to 3.15) | — | — |
| *Fixed effects* | | | | | | |
| Intercept | 25.73 (22.68 to 28.79) | 16.89 (54.2) | <.001 | 25.73 (22.65 to 28.82) | 16.76 (50.9) | <.001 |
| Time | −0.05 (−0.38 to 0.29) | −0.28 (498.0) | .78 | −0.05 (−0.37 to 0.28) | −0.29 (386.7) | .77 |
| Time$^2$ | −0.03 (−0.06 to 0.01) | −1.56 (498.0) | .12 | −0.03 (−0.05 to 0.00) | −1.81 (449.0) | .07 |
| **Model 2.3: Group comparison model (confirmatory)** | | | | | | |
| *Random effects* | | | | | | |
| Intercept (SD) | 10.55 (8.58 to 12.77) | — | — | 10.74 (8.70 to 13.02) | — | — |
| Slope, time (SD) | — | — | — | 0.52 (0.39 to 0.65) | — | — |
| Intercept–slope correlation | — | — | — | −0.19 (−0.47 to 0.13) | — | — |
| Residual (SD) | 3.40 (3.19 to 3.62) | — | — | 2.96 (2.77 to 3.16) | — | — |
| *Fixed effects* | | | | | | |
| Intercept | 26.08 (21.59 to 30.56) | 11.66 (50.3) | <.001 | 26.08 (21.52 to 30.63) | 11.51 (48.0) | <.001 |
| Time | −0.18 (−0.31 to −0.05) | −2.65 (498.0) | .008 | −0.18 (−0.43 to 0.07) | −1.46 (48.0) | .15 |
| Group (Active) | 0.08 (−6.02 to 6.19) | 0.03 (50.3) | .98 | 0.08 (−6.11 to 6.28) | 0.03 (48.0) | .98 |
| Time × Group | −0.23 (−0.41 to −0.05) | −2.53 (498.0) | .01 | −0.23 (−0.57 to 0.10) | −1.40 (48.0) | .17 |
| **Model 2.4a: Covariate of age (confirmatory)** | | | | | | |
| *Random effects* | | | | | | |
| Intercept (SD) | 10.58 (8.51 to 12.67) | — | — | 10.75 (8.62 to 12.90) | — | — |

| Parameter | Random intercept | | | Random intercept and slope | | |
|---|---|---|---|---|---|---|
| | Estimate (95% CI) | t (df) | P | Estimate (95% CI) | t (df) | P |
| Slope, time (SD) | — | — | — | 0.52 (0.39 to 0.65) | — | — |
| Intercept–slope correlation | — | — | — | −0.18 (−0.46 to 0.14) | — | — |
| Residual (SD) | 3.40 (3.19 to 3.61) | — | — | 2.96 (2.77 to 3.16) | — | — |
| *Fixed effects* | | | | | | |
| Intercept | 26.09 (21.59 to 30.60) | 11.64 (49.2) | <.001 | 26.09 (21.53 to 30.66) | 11.50 (47.0) | <.001 |
| Time | −0.18 (−0.31 to −0.05) | −2.66 (497.0) | .008 | −0.18 (−0.43 to 0.07) | −1.45 (47.0) | .15 |
| Group (Active) | 0.05 (−6.08 to 6.18) | 0.02 (49.2) | .99 | 0.05 (−6.16 to 6.26) | 0.02 (47.0) | .99 |
| Age | 0.11 (−0.13 to 0.36) | 0.94 (49.2) | .35 | 0.11 (−0.13 to 0.36) | 0.93 (47.0) | .36 |
| Time × Group | −0.23 (−0.41 to −0.05) | −2.52 (497.0) | .01 | −0.23 (−0.57 to 0.11) | −1.38 (47.0) | .17 |
| Time × Age | 0.00 (−0.01 to 0.00) | −0.65 (497.0) | .52 | 0.00 (−0.02 to 0.01) | −0.36 (47.0) | .72 |
| **Model 2.4b: Covariate of sex (confirmatory)** | | | | | | |
| *Random effects* | | | | | | |
| Intercept (SD) | 9.89 (7.95 to 11.84) | — | — | 10.12 (8.10 to 12.15) | — | — |
| Slope, time (SD) | — | — | — | 0.52 (0.39 to 0.65) | — | — |
| Intercept–slope correlation | — | — | — | −0.21 (−0.49 to 0.11) | — | — |
| Residual (SD) | 3.40 (3.19 to 3.62) | — | — | 2.96 (2.77 to 3.16) | — | — |
| *Fixed effects* | | | | | | |
| Intercept | 29.42 (24.51 to 34.34) | 12.02 (49.5) | <.001 | 29.42 (24.41 to 34.44) | 11.81 (47.0) | <.001 |
| Time | −0.17 (−0.32 to −0.01) | −2.14 (497.0) | .03 | −0.17 (−0.46 to 0.12) | −1.17 (47.0) | .25 |
| Group (Active) | −0.13 (−5.87 to 5.62) | −0.04 (49.5) | .97 | −0.13 (−5.98 to 5.73) | −0.04 (47.0) | .97 |
| Sex (Male) | −7.70 (−13.50 to −1.90) | −2.67 (49.5) | .01 | −7.70 (−13.62 to −1.79) | −2.62 (47.0) | .01 |
| Time × Group | −0.23 (−0.41 to −0.05) | −2.54 (497.0) | .01 | −0.23 (−0.57 to 0.11) | −1.39 (47.0) | .17 |
| Time × Sex | −0.02 (−0.21 to 0.16) | −0.26 (497.0) | .79 | −0.02 (−0.37 to 0.32) | −0.14 (47.0) | .89 |
| **Model 2.4c: Covariate of chronic MDD (confirmatory)** | | | | | | |
| *Random effects* | | | | | | |
| Intercept (SD) | 10.41 (8.37 to 12.46) | — | — | 10.45 (8.37 to 12.54) | — | — |
| Slope, time (SD) | — | — | — | 0.51 (0.37 to 0.64) | — | — |
| Intercept–slope correlation | — | — | — | −0.13 (−0.42 to 0.19) | — | — |
| Residual (SD) | 3.38 (3.17 to 3.59) | — | — | 2.96 (2.77 to 3.16) | — | — |
| *Fixed effects* | | | | | | |
| Intercept | 23.58 (18.42 to 28.73) | 9.19 (49.3) | <.001 | 23.58 (18.42 to 28.74) | 9.19 (47.0) | <.001 |
| Time | −0.07 (−0.23 to 0.08) | −0.95 (497.0) | .34 | −0.07 (−0.36 to 0.21) | −0.53 (47.0) | .60 |
| Group (Active) | 0.03 (−6.00 to 6.06) | 0.01 (49.3) | >.99 | 0.03 (−6.01 to 6.07) | 0.01 (47.0) | >.99 |
| Chronic MDD | 5.75 (−0.31 to 11.80) | 1.91 (49.3) | .06 | 5.75 (−0.32 to 11.81) | 1.91 (47.0) | .06 |
| Time × Group | −0.23 (−0.41 to −0.05) | −2.52 (497.0) | .01 | −0.23 (−0.56 to 0.10) | −1.40 (47.0) | .17 |

| Parameter | Random intercept | | | Random intercept and slope | | |
|---|---|---|---|---|---|---|
| | Estimate (95% CI) | t (df) | P | Estimate (95% CI) | t (df) | P |
| Time × Chronic MDD | −0.24 (−0.42 to −0.06) | −2.64 (497.0) | .009 | −0.24 (−0.58 to 0.09) | −1.46 (47.0) | .15 |
| **Model 2.5b: Moderation by episode duration (confirmatory)** | | | | | | |
| *Random effects* | | | | | | |
| Intercept (SD) | 10.34 (8.23 to 12.25) | — | — | 10.63 (8.42 to 12.62) | — | — |
| Slope, time (SD) | — | — | — | 0.52 (0.38 to 0.64) | — | — |
| Intercept–slope correlation | — | — | — | −0.23 (−0.50 to 0.09) | — | — |
| Residual (SD) | 3.39 (3.18 to 3.60) | — | — | 2.96 (2.77 to 3.16) | — | — |
| *Fixed effects* | | | | | | |
| Intercept | 26.11 (22.92 to 29.29) | 16.49 (48.3) | <.001 | 26.11 (22.84 to 29.37) | 16.11 (46.0) | <.001 |
| Time | −0.32 (−0.42 to −0.23) | −6.59 (496.0) | <.001 | −0.32 (−0.50 to −0.14) | −3.59 (46.0) | <.001 |
| Group (Active) | −1.76 (−8.12 to 4.61) | −0.55 (48.3) | .58 | −1.76 (−8.28 to 4.77) | −0.54 (46.0) | .59 |
| Episode duration | 7.04 (−1.26 to 15.33) | 1.71 (48.3) | .09 | 7.04 (−1.46 to 15.53) | 1.67 (46.0) | .10 |
| Time × Group | −0.28 (−0.47 to −0.09) | −2.88 (496.0) | .004 | −0.28 (−0.64 to 0.08) | −1.57 (46.0) | .12 |
| Time × Episode duration | 0.20 (−0.05 to 0.45) | 1.55 (496.0) | .12 | 0.20 (−0.27 to 0.67) | 0.85 (46.0) | .40 |
| Group × Episode duration | 1.30 (−15.28 to 17.89) | 0.16 (48.3) | .88 | 1.30 (−15.69 to 18.29) | 0.15 (46.0) | .88 |
| Time × Group × Episode duration | 0.42 (−0.08 to 0.91) | 1.64 (496.0) | .10 | 0.42 (−0.52 to 1.35) | 0.89 (46.0) | .38 |
| **Model 2.6: Final combined model (confirmatory)** | | | | | | |
| *Random effects* | | | | | | |
| Intercept (SD) | 9.86 (7.85 to 11.69) | — | — | 9.97 (7.90 to 11.85) | — | — |
| Slope, time (SD) | — | — | — | 0.51 (0.37 to 0.64) | — | — |
| Intercept–slope correlation | — | — | — | −0.16 (−0.45 to 0.16) | — | — |
| Residual (SD) | 3.38 (3.17 to 3.59) | — | — | 2.96 (2.77 to 3.16) | — | — |
| *Fixed effects* | | | | | | |
| Intercept | 27.29 (21.55 to 33.03) | 9.56 (47.8) | <.001 | 27.24 (21.48 to 33.00) | 9.51 (46.8) | <.001 |
| Time | −0.07 (−0.23 to 0.08) | −0.95 (497.0) | .34 | −0.07 (−0.36 to 0.21) | −0.53 (47.0) | .60 |
| Group (Active) | −0.16 (−5.89 to 5.58) | −0.05 (48.5) | .96 | −0.15 (−5.93 to 5.62) | −0.05 (46.1) | .96 |
| Sex (Male) | −7.23 (−13.05 to −1.41) | −2.50 (46.0) | .02 | −7.13 (−12.94 to −1.33) | −2.48 (46.0) | .02 |
| Chronic MDD | 4.43 (−1.42 to 10.28) | 1.52 (48.4) | .13 | 4.45 (−1.44 to 10.34) | 1.52 (46.2) | .14 |
| Time × Group | −0.23 (−0.41 to −0.05) | −2.52 (497.0) | .01 | −0.23 (−0.56 to 0.10) | −1.40 (47.0) | .17 |
| Time × Chronic MDD | −0.24 (−0.42 to −0.06) | −2.64 (497.0) | .009 | −0.24 (−0.58 to 0.09) | −1.46 (47.0) | .15 |
| **Model 2.10: Moderation by personality disorder (exploratory)** | | | | | | |
| *Random effects* | | | | | | |
| Intercept (SD) | 10.11 (8.04 to 11.98) | — | — | 10.52 (8.33 to 12.49) | — | — |
| Slope, time (SD) | — | — | — | 0.51 (0.37 to 0.63) | — | — |
| Intercept–slope correlation | — | — | — | −0.27 (−0.54 to 0.05) | — | — |

| Parameter | Random intercept | | | Random intercept and slope | | |
|---|---|---|---|---|---|---|
| | Estimate (95% CI) | t (df) | P | Estimate (95% CI) | t (df) | P |
| Residual (SD) | 3.38 (3.17 to 3.59) | — | — | 2.96 (2.77 to 3.16) | — | — |
| *Fixed effects* | | | | | | |
| Intercept | 25.37 (20.20 to 30.54) | 9.86 (48.3) | <.001 | 25.37 (20.01 to 30.73) | 9.53 (46.0) | <.001 |
| Time | −0.16 (−0.32 to 0.00) | −2.01 (496.0) | .045 | −0.16 (−0.46 to 0.13) | −1.10 (46.0) | .28 |
| Group (Active) | −1.33 (−8.19 to 5.53) | −0.39 (48.3) | .70 | −1.33 (−8.44 to 5.79) | −0.38 (46.0) | .71 |
| Personality disorder (PD) | 2.31 (−7.06 to 11.68) | 0.50 (48.3) | .62 | 2.31 (−7.41 to 12.02) | 0.48 (46.0) | .63 |
| Time × Group | −0.34 (−0.55 to −0.13) | −3.18 (496.0) | .002 | −0.34 (−0.73 to 0.05) | −1.75 (46.0) | .09 |
| Time × PD | −0.06 (−0.34 to 0.23) | −0.39 (496.0) | .69 | −0.06 (−0.59 to 0.48) | −0.22 (46.0) | .83 |
| Group × PD | 7.22 (−6.18 to 20.61) | 1.08 (48.3) | .28 | 7.22 (−6.67 to 21.10) | 1.05 (46.0) | .30 |
| Time × Group × PD | 0.46 (0.05 to 0.87) | 2.21 (496.0) | .03 | 0.46 (−0.30 to 1.23) | 1.21 (46.0) | .23 |
| **Model 2.11: Moderation by childhood trauma (exploratory)** | | | | | | |
| *Random effects* | | | | | | |
| Intercept (SD) | 9.85 (7.84 to 11.68) | — | — | 10.01 (7.92 to 11.89) | — | — |
| Slope, time (SD) | — | — | — | 0.52 (0.38 to 0.64) | — | — |
| Intercept–slope correlation | — | — | — | −0.18 (−0.47 to 0.14) | — | — |
| Residual (SD) | 3.39 (3.18 to 3.60) | — | — | 2.96 (2.77 to 3.16) | — | — |
| *Fixed effects* | | | | | | |
| Intercept | 27.94 (23.50 to 32.38) | 12.65 (48.5) | <.001 | 27.94 (23.45 to 32.43) | 12.52 (46.0) | <.001 |
| Time | −0.22 (−0.36 to −0.08) | −3.05 (496.0) | .002 | −0.22 (−0.48 to 0.05) | −1.67 (46.0) | .10 |
| Group (Active) | −2.44 (−8.41 to 3.52) | −0.82 (48.5) | .41 | −2.44 (−8.48 to 3.60) | −0.81 (46.0) | .42 |
| Childhood trauma (CTQ) | 50.86 (12.32 to 89.41) | 2.65 (48.5) | .01 | 50.86 (11.85 to 89.88) | 2.62 (46.0) | .01 |
| Time × Group | −0.22 (−0.41 to −0.03) | −2.28 (496.0) | .02 | −0.22 (−0.57 to 0.13) | −1.25 (46.0) | .22 |
| Time × CTQ | −1.01 (−2.23 to 0.20) | −1.64 (496.0) | .10 | −1.01 (−3.29 to 1.26) | −0.90 (46.0) | .37 |
| Group × CTQ | −29.62 (−77.77 to 18.54) | −1.24 (48.5) | .22 | −29.62 (−78.36 to 19.13) | −1.22 (46.0) | .23 |
| Time × Group × CTQ | 1.73 (0.21 to 3.24) | 2.24 (496.0) | .03 | 1.73 (−1.11 to 4.57) | 1.22 (46.0) | .23 |
| **Model 2.12: Moderation by relationship status (exploratory)** | | | | | | |
| *Random effects* | | | | | | |
| Intercept (SD) | 10.31 (8.21 to 12.22) | — | — | 10.54 (8.35 to 12.51) | — | — |
| Slope, time (SD) | — | — | — | 0.53 (0.39 to 0.65) | — | — |
| Intercept–slope correlation | — | — | — | −0.20 (−0.48 to 0.12) | — | — |
| Residual (SD) | 3.41 (3.19 to 3.62) | — | — | 2.96 (2.77 to 3.16) | — | — |
| *Fixed effects* | | | | | | |
| Intercept | 22.29 (16.44 to 28.14) | 7.66 (48.3) | <.001 | 22.29 (16.33 to 28.25) | 7.53 (46.0) | <.001 |
| Time | −0.20 (−0.37 to −0.02) | −2.18 (496.0) | .03 | −0.20 (−0.53 to 0.14) | −1.18 (46.0) | .24 |
| Group (Active) | 4.17 (−4.47 to 12.81) | 0.97 (48.3) | .34 | 4.17 (−4.63 to 12.97) | 0.95 (46.0) | .35 |

| Parameter | Random intercept | | | Random intercept and slope | | |
|---|---|---|---|---|---|---|
| | Estimate (95% CI) | t (df) | P | Estimate (95% CI) | t (df) | P |
| Relationship | 8.71 (−0.17 to 17.58) | 1.97 (48.3) | .054 | 8.71 (−0.33 to 17.74) | 1.94 (46.0) | .06 |
| Time × Group | −0.24 (−0.50 to 0.02) | −1.80 (496.0) | .07 | −0.24 (−0.73 to 0.26) | −0.97 (46.0) | .34 |
| Time × Relationship | 0.04 (−0.23 to 0.31) | 0.29 (496.0) | .77 | 0.04 (−0.47 to 0.55) | 0.16 (46.0) | .88 |
| Group × Relationship | −9.21 (−21.33 to 2.91) | −1.53 (48.3) | .13 | −9.21 (−21.55 to 3.13) | −1.50 (46.0) | .14 |
| Time × Group × Relationship | 0.00 (−0.37 to 0.37) | −0.00 (496.0) | >.99 | 0.00 (−0.70 to 0.69) | −0.00 (46.0) | >.99 |
| **Model 2.13: Moderation by social network size (exploratory)** | | | | | | |
| *Random effects* | | | | | | |
| Intercept (SD) | 10.13 (8.04 to 12.02) | — | — | 10.09 (7.96 to 12.00) | — | — |
| Slope, time (SD) | — | — | — | 0.52 (0.38 to 0.64) | — | — |
| Intercept–slope correlation | — | — | — | −0.11 (−0.41 to 0.22) | — | — |
| Residual (SD) | 3.41 (3.19 to 3.62) | — | — | 2.97 (2.78 to 3.18) | — | — |
| *Fixed effects* | | | | | | |
| Intercept | 27.02 (22.60 to 31.45) | 12.28 (47.3) | <.001 | 27.02 (22.63 to 31.42) | 12.38 (45.0) | <.001 |
| Time | −0.20 (−0.34 to −0.06) | −2.88 (486.0) | .004 | −0.20 (−0.45 to 0.06) | −1.57 (45.0) | .12 |
| Group (Active) | −0.97 (−6.94 to 4.99) | −0.33 (47.3) | .74 | −0.97 (−6.90 to 4.95) | −0.33 (45.0) | .74 |
| Social network size | −15.00 (−32.49 to 2.49) | −1.73 (47.3) | .09 | −15.00 (−32.37 to 2.37) | −1.74 (45.0) | .09 |
| Time × Group | −0.21 (−0.39 to −0.03) | −2.24 (486.0) | .03 | −0.21 (−0.55 to 0.13) | −1.22 (45.0) | .23 |
| Time × Social network size | 0.33 (−0.21 to 0.87) | 1.20 (486.0) | .23 | 0.33 (−0.68 to 1.34) | 0.65 (45.0) | .52 |
| Group × Social network size | 3.47 (−18.25 to 25.19) | 0.32 (47.3) | .75 | 3.47 (−18.10 to 25.04) | 0.32 (45.0) | .75 |
| Time × Group × Social network size | 0.08 (−0.59 to 0.74) | 0.22 (486.0) | .82 | 0.08 (−1.18 to 1.33) | 0.12 (45.0) | .90 |
| **Model 2.14: Moderation by loneliness (exploratory)** | | | | | | |
| *Random effects* | | | | | | |
| Intercept (SD) | 9.56 (7.58 to 11.34) | — | — | 9.70 (7.65 to 11.53) | — | — |
| Slope, time (SD) | — | — | — | 0.53 (0.39 to 0.66) | — | — |
| Intercept–slope correlation | — | — | — | −0.18 (−0.47 to 0.15) | — | — |
| Residual (SD) | 3.42 (3.21 to 3.63) | — | — | 2.97 (2.78 to 3.18) | — | — |
| *Fixed effects* | | | | | | |
| Intercept | 26.54 (22.34 to 30.74) | 12.71 (47.6) | <.001 | 26.54 (22.29 to 30.78) | 12.59 (45.0) | <.001 |
| Time | −0.19 (−0.33 to −0.05) | −2.73 (486.0) | .007 | −0.19 (−0.45 to 0.07) | −1.47 (45.0) | .15 |
| Group (Active) | −0.02 (−5.67 to 5.63) | −0.01 (47.6) | >.99 | −0.02 (−5.73 to 5.69) | −0.01 (45.0) | >.99 |
| Loneliness | 0.36 (−0.11 to 0.83) | 1.55 (47.6) | .13 | 0.36 (−0.11 to 0.84) | 1.53 (45.0) | .13 |
| Time × Group | −0.22 (−0.41 to −0.04) | −2.35 (486.0) | .02 | −0.22 (−0.57 to 0.13) | −1.27 (45.0) | .21 |
| Time × Loneliness | −0.01 (−0.02 to 0.01) | −0.75 (486.0) | .46 | −0.01 (−0.04 to 0.02) | −0.40 (45.0) | .69 |
| Group × Loneliness | 0.16 (−0.44 to 0.76) | 0.54 (47.6) | .59 | 0.16 (−0.44 to 0.76) | 0.53 (45.0) | .60 |
| Time × Group × Loneliness | 0.01 (−0.01 to 0.03) | 0.68 (486.0) | .49 | 0.01 (−0.03 to 0.04) | 0.37 (45.0) | .71 |

| Parameter | Random intercept | | | Random intercept and slope | | |
|---|---|---|---|---|---|---|
| | Estimate (95% CI) | t (df) | P | Estimate (95% CI) | t (df) | P |
| **Model 2.15: Moderation by employment status (exploratory)** | | | | | | |
| *Random effects* | | | | | | |
| Intercept (SD) | 10.35 (8.23 to 12.26) | — | — | 10.29 (8.15 to 12.22) | — | — |
| Slope, time (SD) | — | — | — | 0.51 (0.37 to 0.62) | — | — |
| Intercept–slope correlation | — | — | — | −0.10 (−0.39 to 0.23) | — | — |
| Residual (SD) | 3.37 (3.16 to 3.57) | — | — | 2.96 (2.77 to 3.16) | — | — |
| *Fixed effects* | | | | | | |
| Intercept | 30.78 (24.91 to 36.64) | 10.55 (48.2) | <.001 | 30.78 (24.96 to 36.60) | 10.65 (46.0) | <.001 |
| Time | −0.37 (−0.55 to −0.20) | −4.17 (496.0) | <.001 | −0.37 (−0.69 to −0.05) | −2.31 (46.0) | .03 |
| Group (Active) | −4.22 (−12.23 to 3.80) | −1.06 (48.2) | .30 | −4.22 (−12.17 to 3.74) | −1.07 (46.0) | .29 |
| Employment (Employed) | −10.82 (−19.72 to −1.93) | −2.45 (48.2) | .02 | −10.82 (−19.65 to −1.99) | −2.47 (46.0) | .02 |
| Time × Group | −0.09 (−0.33 to 0.15) | −0.74 (496.0) | .46 | −0.09 (−0.53 to 0.35) | −0.41 (46.0) | .68 |
| Time × Employment | 0.44 (0.18 to 0.71) | 3.27 (496.0) | .001 | 0.44 (−0.05 to 0.93) | 1.81 (46.0) | .08 |
| Group × Employment | 9.91 (−2.18 to 22.00) | 1.65 (48.2) | .11 | 9.91 (−2.09 to 21.91) | 1.66 (46.0) | .10 |
| Time × Group × Employment | −0.33 (−0.69 to 0.03) | −1.80 (496.0) | .07 | −0.33 (−1.00 to 0.34) | −1.00 (46.0) | .32 |
| **Piecewise follow-up model (time in days; random slope for the acute phase) (exploratory)** | | | | | | |
| *Random effects* | | | | | | |
| Intercept (SD) | 10.56 (8.58 to 12.78) | — | — | 10.74 (8.68 to 13.04) | — | — |
| Slope, time in acute phase (SD) | — | — | — | 0.55 (0.42 to 0.70) | — | — |
| Intercept–slope correlation | — | — | — | −0.20 (−0.48 to 0.13) | — | — |
| Residual (SD) | 3.94 (3.70 to 4.17) | — | — | 3.45 (3.24 to 3.67) | — | — |
| *Fixed effects* | | | | | | |
| Intercept | 26.08 (21.56 to 30.59) | 11.59 (51.1) | <.001 | 26.08 (21.50 to 30.65) | 11.46 (48.1) | <.001 |
| Time (Acute phase) | −0.18 (−0.33 to −0.03) | −2.29 (540.0) | .02 | −0.18 (−0.45 to 0.09) | −1.33 (52.5) | .19 |
| Group (Active) | 0.08 (−6.06 to 6.23) | 0.03 (51.1) | .98 | 0.08 (−6.14 to 6.31) | 0.03 (48.1) | .98 |
| Time (Follow-up phase) | −0.09 (−0.13 to −0.04) | −3.57 (540.1) | <.001 | −0.08 (−0.12 to −0.04) | −3.92 (497.9) | <.001 |
| Time (Acute) × Group | −0.23 (−0.44 to −0.02) | −2.19 (540.0) | .03 | −0.23 (−0.60 to 0.13) | −1.27 (52.5) | .21 |
| Time (Follow-up) × Group | 0.01 (−0.05 to 0.07) | 0.36 (540.1) | .72 | 0.01 (−0.04 to 0.07) | 0.43 (497.5) | .67 |

Abbreviations: BDI-II, Beck Depression Inventory-II; CI, confidence interval; MDD, major depressive disorder; SD, standard deviation.

Note: Parameter estimates from restricted maximum likelihood. Fixed effects with t tests and Satterthwaite degrees of freedom (in parentheses) and 95% CIs from the t distribution with these degrees of freedom; random-effect SDs and the intercept–slope correlation with profile-likelihood 95% CIs. Time was coded in days (0–10; follow-up model: acute phase days 0–10, follow-up phase days 10–52), so estimates for time and its interactions are changes in BDI-II score per day. Group was coded 0 (sham) and 1 (active); continuous moderators were mean-centered and, where skewed, log-transformed. In Model 2.5b, group was effect-coded (sham, −0.5; active, +0.5), so that the intercept, time, and episode-duration effects refer to the average of both groups. One participant with a baseline BDI-II score below 14 was excluded (n = 50). Model fit is given in Table S6, panel B.
